# Safety and feasibility of fecal microbiota transplantation for Parkinson’s disease patients: a protocol for a self-controlled interventional donor-FMT pilot study

**DOI:** 10.1101/2022.05.23.22274942

**Authors:** Karuna E.W. Vendrik, Vlada O. Bekker-Chernova, Ed J. Kuijper, Elisabeth M. Terveer, Jacobus J. van Hilten, Maria Fiorella Contarino, the FMT4PD study group

**Author notes:** Corresponding author: E: mail (MFC).

## Abstract

Several experimental studies suggest a role of the gut microbiota in the pathophysiology of Parkinson’s disease (PD) via the gut-brain axis. In addition, the gut microbiota can influence the metabolism of levodopa, which is the mainstay of treatment of PD. Therefore, modifying the gut microbiota by fecal microbiota transplantation (FMT) could be a supportive treatment strategy. We developed a study protocol for a single center, prospective, self-controlled, interventional, safety and feasibility donor-FMT pilot study with randomization and double blinded allocation of donor feces. The primary objectives are to assess feasibility and safety of FMT in PD patients. Secondary objectives are to explore whether FMT leads to alterations of motor complications and PD symptoms in the short term, determine alterations in gut microbiota composition and donor-recipient microbiota similarities and their association with PD symptoms and motor complications, assess the ease of the study protocol and examine FMT-related adverse events in PD patients. The study population will consist of 16 idiopathic PD patients that use levodopa and experience motor complications. They will receive FMT with feces from one of two selected healthy human donors. Patients will be pretreated with vancomycin and bowel lavage to increase donor feces engraftment, and domperidone to prevent nausea and regurgitation. The FMT will be performed via direct injection of the fecal suspension into the duodenum via a gastroscope. There will be six follow-up moments during three months. Blood will be drawn on three occasions. Physical examination, filling in of questionnaires and a diary, and collection of stool samples are performed at baseline and each in-person follow-up visit (except for the FMT visit). This study was approved by the Medical Ethical Committee Leiden Den Haag Delft (ref. P20.087) and is registered in the Netherlands Trial Register (NL9438). Study results will be disseminated through publication in peer-reviewed journals and international conferences.

## Introduction

Parkinson’s disease (PD) is characterized by the degeneration of neurons and the presence of Lewy bodies and Lewy neuritis in the central nervous system (CNS), enteric nervous system (ENS) and peripheral autonomic nervous system[1]. The etiology and pathogenesis of PD is still largely unknown, although a role for the aggregation of alpha-synuclein (αSyn) is generally acknowledged[2].

GI symptoms (including obstipation and delayed transit) are frequently observed in PD patients and often precede the onset of motor symptoms[3, 4]. Alpha-synucleinopathy is present in the ENS and vagal nerves in an early phase of disease[5-10]. This led to the hypothesis that the disease may start in the gut, with misfolded αSyn fragments being transported to the CNS through the vagal nerve[5, 11-14]. The hypothesis is supported by studies suggesting that αSyn forms could be transported from the gut to the brain[12-14]. Furthermore, it is suggested that aggregation of αSyn in the brain and gut is a consequence of inflammation Dinduced oxidative stress[15-17].

The gut microbiota is the community of micro-organisms that resides in the gut. Several recent studies indicate that the gut microbiota and their metabolic products in PD patients differ from healthy individuals[16, 18-24], with a more pro-inflammatory and less anti-inflammatory composition in PD[16]. Specific taxa appear to be associated with symptom severity[20] and gut bacterial tyrosine decarboxylases can metabolize levodopa to dopamine without being susceptible for carbidopa, which may alter the bioavailability of levodopa[25, 26]. Based on the available data, it is hypothesized that interventions aimed at modifying the gut microbiota could influence PD symptoms severity and disease progression and/or improve levodopa absorption and efficacy, resulting in a decrease of levodopa-mediated motor complications. Fecal microbiota transplantation (FMT) could potentially restore the disturbed gut microbiota composition and metabolic activity of the microbiota [27-29]. FMT is an effective and safe treatment for multiple recurrent[30-32] and severe[33] *Clostridioides difficile* infections (CDI). Serious adverse events in this patient category have been described, but occur in only 0-5% of patients[34-36]. Currently, CDI is the only registered indication for FMT[37-39], but preliminary data on FMT in several neurological disorders are becoming available [40].

Since there are no available treatments to cure or slow down the progression of PD, and most patients with advanced disease experience decreased efficacy and/or adverse effects of medication, the development of a new treatment strategy is crucial. A potential beneficial effect of FMT in PD is shown in several mouse studies [41-43]. Recently, one case report[44] and three case series (15, 11, and 6 patients)[45-47] have been published reporting the results of FMT in PD patients. In general, some improvement of motor and non-motor symptoms, including constipation, was reported in all series. Gut microbiota analysis was reported in only one case report and one case series (11 patients)[44, 47], which showed significant changes in the gut microbiota. However, large variability of methods concerning pre-treatment, FMT administration route, follow-up and clinical evaluation exists. No results of randomized clinical trials on FMT in PD patients have been reported yet. We report the protocol of a single center, prospective, self-controlled, interventional safety and feasibility donor-FMT pilot study with randomization and double blinded allocation of donor feces. The primary objective of the study is to demonstrate that FMT is feasible and safe in this patient group. In addition, we hypothesize that FMT will lead to a decrease of motor complications, and improvement of PD symptoms, also including constipation, in the short term. Considering the scanty availability of data on FMT in this patient population, we have decided to focus on treatment safety without a control group. To correct for the known variability in occurrence and severity of both motor and gastrointestinal symptoms, we have introduced a ‘standard-of-care evaluation’ as comparator for these outcomes. In order to control for possible donor-related effects, feces of two donors will be randomly assigned.

## Materials and Methods

### Objectives and study design

The study was designed in close collaboration with the Dutch Parkinson patients association, with participation of “patient researchers”. The primary objectives are to assess feasibility and safety of FMT in PD patients. Secondary objectives are to explore whether FMT leads to alterations of motor complications (fluctuations or dyskinesias) and PD motor and non-motor symptoms (including constipation) in the short term, determine alterations in gut microbiota composition and donor-recipient microbiota similarities and their association with PD symptoms and motor complications, assess the ease of the study protocol and examine FMT-related adverse events (AEs) in PD patients.

The study is a single center prospective self-controlled interventional safety and feasibility donor-FMT pilot study with randomization and double blinded allocation of donor feces. Sixteen patients will be included. The follow-up period will be three months. The study site is Leiden University Medical Center. The 2013 SPIRIT checklist and the more detailed approved study protocol (version 3, October 2020) are shown in the Supplementary S1 and S2 Files. Fig 1 provides an overview of all study procedures. In Fig 2, an overview of the study design is shown.

**Fig 1.**
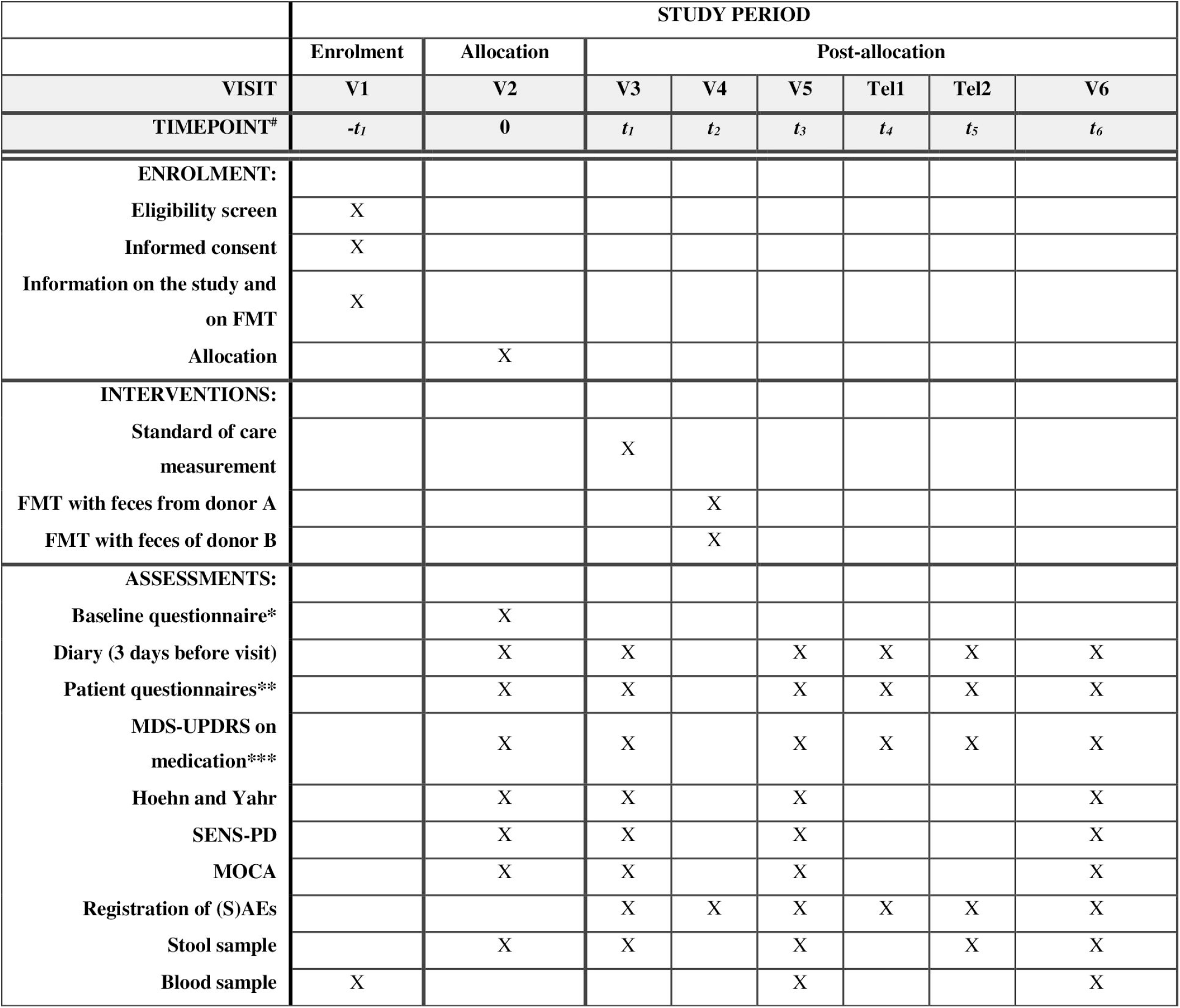
Schedule of study procedures. ^#^ -t_1_: screening visit: unspecified timepoint, t_1_: 1 week post-allocation (standard-of-care), t_2_: FMT: unspecified timepoint, t_3_: 1 week post-FMT, t_4_: 2 weeks post-FMT, t_5_: 6 weeks post-FMT, t_6_: 3 months post-FMT. *The baseline questionnaire includes questions on health status, disease-related variables and medication use (PD and non-PD). **Patient questionnaires are filled in by the participant prior to a visit/telephone appointment and include questions on health status, diet, medication use, constipation (Cleveland clinic constipation score[48] and ROME IV constipation criteria[49]), SENS-PD[50], Q10 (wearing off)[51], and MDS-UPDRS IB and II (and a study load questionnaire at V6)[52]. *** MDS-UPDRS IA, III and IV (III not during telephone appointments)[52]. Abbreviations: FMT: fecal microbiota transplantation, MDS-UPDRS: Movement Disorder Society-Sponsored Revision of the Unified Parkinson’s Disease Rating Scale[52], MOCA: Montreal Cognitive Assessment[53], (S)AEs: (serious) adverse events, SENS-PD: SEverity of Non-dopaminergic Symptoms in Parkinson’s Disease[50], Tel: telephone appointment, V: visit.

**Fig 2.**
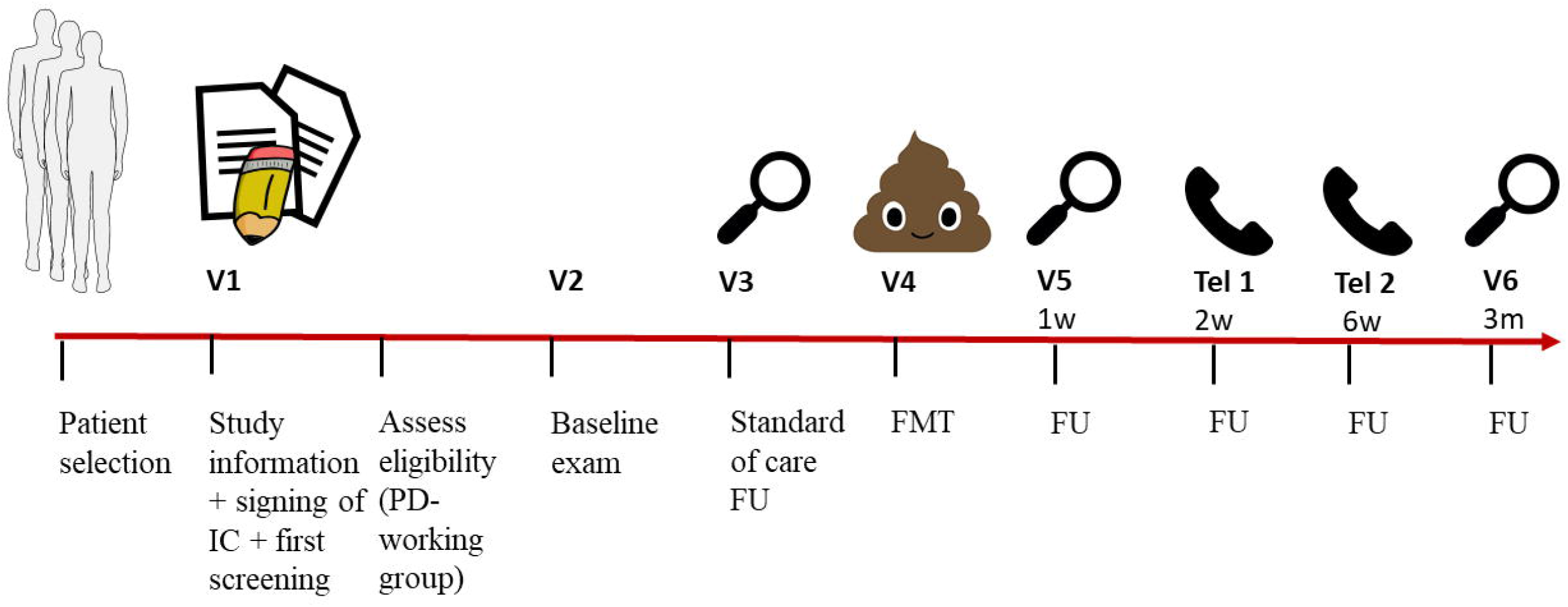
Graphical abstract of study design. Abbreviations: FMT: fecal microbiota transplantation, FU: follow-up, IC: informed consent, m: months, PD: Parkinson’s disease, Tel: telephone appointment, V: visit, w: week(s).

### Patient selection and characteristics of study population

PD patients will be primarily recruited in the LUMC, and, if needed, from other hospitals by using advertisements. The study population will consist of 16 idiopathic stable PD patients with motor complications despite adequate medication. In- and exclusion criteria are reported in Table 1. During the study, PD patients are allowed to increase or decrease the dosage of medication or change the type of medication if needed. This will be taken into account for the analysis and interpretation of the study results.

**Table 1.**
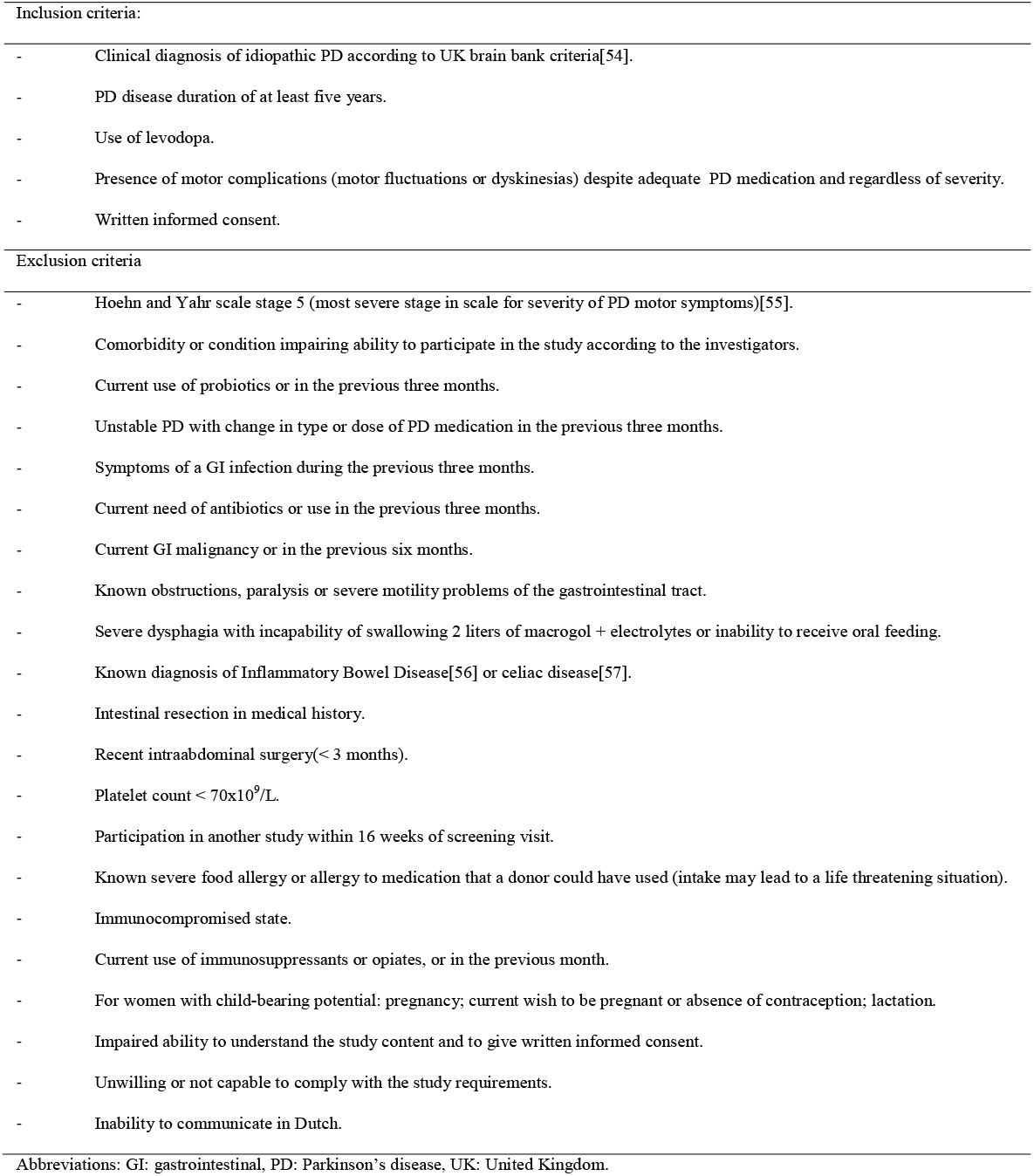
In- and exclusion criteria for patients with PD to participate in the FMT4PD study.

All subjects will receive one FMT. To assess the variability of the study endpoints and to provide self-control data, two standard-of-care measurements (baseline and V3) will be performed before FMT.

The Netherlands Donor Feces Bank (NDFB - http://www.ndfb.nl/), located in the LUMC, provides ready-to-use quality assured fecal suspensions from healthy donors for FMT in the Netherlands. General protocols for screening of donors and preparation of fecal suspensions in use at the NDFB have been described before.[36, 58] Importantly, persons with constipation cannot become a donor and donors are asked whether there are any genetic diseases in the family. Two donors will be selected from the donor pool of the NDFB to minimize the risk of no or a negative response due to donor specific characteristics and to explore which donor gut microbiota characteristics are beneficial for PD patients. The donor selection will be randomized and double-blind. An employee of the NDFB will use the cloud-based Castor clinical data management platform to produce a randomization list for the two donors using a variable block randomization method, that will not be disclosed to the investigators (that enroll participants)/physicians/patients involved in this trial. A technician will be informed of the outcome of randomisation for each patient, will prepare the selected material and will ensure that the syringes with the fecal suspension do not contain donor identifying information. The randomization code could be broken in case of suspected FMT-related infections or adverse reactions, or when the Data safety monitoring Board (DSMB) deems it necessary.

### Sample size

Since this is a pilot study, only 16 patients will be included. This is the number that is needed to have >80% chance that any FMT-related SAEs, that occur in >10% of the cases, might occur in the current study population. The occurrence of FMT-related SAEs in >10% of the PD patients is deemed useful information that might change the design of a future clinical trial or might result in the choice not to perform such a trial.

### Study procedures

#### Screening

Selected patients will receive a patient information letter. If interested, the patient will be further informed on the study during the first visit. The research physician will determine whether the patient meets the in- and exclusion criteria and is able to participate. If the patient agrees to participate, the informed consent form will be signed in presence of the investigator. Thereafter, blood and clinical questionnaires concerning sociodemographic variables, present and past medical history and medication use will be collected. If participants give permission for the LUMC Biobank Parkinson, their blood samples (and some DNA from the blood) will be stored for indefinite duration for future (yet unknown) analyses. The final eligibility of the patient will be discussed in the “Parkinson working group”, including at least one infectious disease specialist, gastroenterologist, medical microbiologists (FMT experts) and neurologist (PD expert).

#### Clinical evaluations

In the three days before the baseline visit, the patient will fill in a diary to describe the motor complications during the day. The day before the baseline examination, the patients will fill in a questionnaire, including questions on health status, diet, constipation, disease-related variables, medication use and motor and non-motor symptoms during the week previous to the visit. Furthermore, on the day of the visit, the investigators will complete an additional baseline questionnaire with more detailed questions about health status, disease-related variables (motor and non-motor symptoms) and medication use and will perform a physical examination. Patients will be instructed to report all SAEs immediately to the investigators during the study period. (S)AEs will also be assessed at each visit using a standardized form. During the standard-of-care visit and post-FMT follow-up the same evaluations will be repeated (except for the baseline questionnaire). Additional blood samples will be collected at one week and three months post-FMT. During the last visit, the study load will be assessed.

#### Stool sampling

During this study, stool samples are collected for analysis and evaluation of the FMT treatment effect and (S)AEs. The baseline stool sample, including all feces from one defecation collected in a fecotainer within four hours after defecation before the baseline visit, will also be used for the preparation of an autologous fecal suspension to be stored for a potential rescue FMT. Additional stool (using feces collection tubs) will be collected at one week after baseline (standard-of-care visit), and one week, six weeks and three months post-FMT. Patients will be requested to collect stool samples of each defecation from three days before a study visit, or earlier if the patient has severe constipation, until the visit, and to store it in the refrigerator. The most fresh stool sample will be delivered to the laboratory at the regular study visit. At six weeks post-FMT, patients will be requested to send a stool sample by mail, as soon as possible after defecation, with storage in the refrigerator until transport. The stool samples will be stored and can be retrieved for microbiota analysis, culturing purposes, safety reasons (SAEs) or future research purposes.

Fecal suspensions and stool samples are stored in a -80°C freezer. The suspension for FMT contains 198 ml and is derived from 60 gram feces with added glycerol up to a percentage of 10% as cryoprotectant. Autologous suspensions are allowed to be 99 ml, which means that if the baseline stool sample does not contain at least 33 gram, the patient will be asked to collect another stool sample. For the other stool samples, at least two times 1 gram feces is stored with 10% glycerol for culturing purposes and at least two times 1 gram feces is stored for microbiota analysis. In addition, when there is more feces left and if participants give permission for LUMC Biobank Parkinson storage, two aliquots of 1 gram with 10% glycerol and two aliquots of 1 gram without glycerol will be stored in the LUMC Biobank Parkinson for future research purposes. Stool samples for this study will be destroyed 20 years after end of the study or for indefinite duration when stored in the LUMC Biobank Parkinson. The feces consistency for every stool sample will be registered by the patient and the investigator using the Bristol stool scale.

#### FMT procedure

The patients will receive a healthy donor FMT in the hospital via direct injection into the horizontal duodenum through a gastroscope. Defrosted ready-to-use fecal suspensions will be provided by the NDFB. The pre-treatment includes 2 liters of laxatives (macrogol + electrolytes) on the day prior to FMT, and vancomycin 250 milligram (mg) four times per day for five days until 24 hours before FMT.[58] In case of obstipation, additional laxatives (Bisacodyl, maximum 2 times 5 mg per day) will be administered in the two days before FMT to improve the efficacy of the bowel lavage. When this is not contraindicated, one pill of domperidone 10 mg will be administered orally on the day of FMT prior to FMT, to prevent nausea and to improve gastric motility. Domperidone could also be used after FMT, in case of nausea or vomiting. When preferred, mild sedation by intravenous administration of 0.5–7,5 mg midazolam before or during gastroscopy can be provided. The post-FMT observation period with regular vital parameter checks in the hospital will be at least two hours.

### Outcomes

Study parameters/endpoints are shown in Table 2.

**Table 2.**
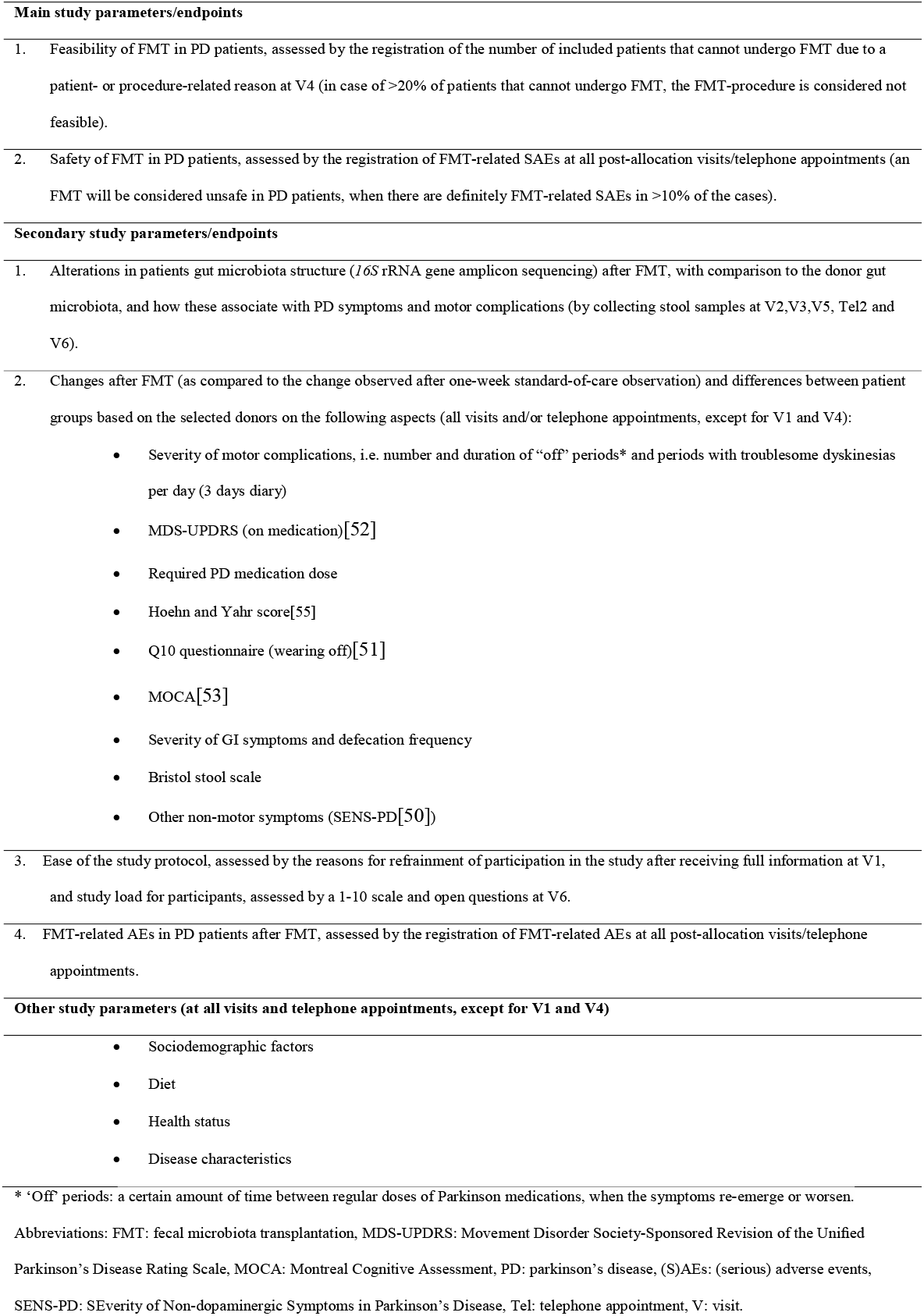
Study parameters/endpoints.

| **Main study parameters/endpoints** |  |
| --- | --- |
| 1. | Feasibility of FMT in PD patients, assessed by the registration of the number of included patients that cannot undergo FMT due to a patient- or procedure-related reason at V4 (in case of >20% of patients that cannot undergo FMT, the FMT-procedure is considered not feasible). |
| 2. | Safety of FMT in PD patients, assessed by the registration of FMT-related SAEs at all post-allocation visits/telephone appointments (an FMT will be considered unsafe in PD patients, when there are definitely FMT-related SAEs in >10% of the cases). |
| **Secondary study parameters/endpoints** |  |
| 1. | Alterations in patients gut microbiota structure (*16S* rRNA gene amplicon sequencing) after FMT, with comparison to the donor gut microbiota, and how these associate with PD symptoms and motor complications (by collecting stool samples at V2,V3,V5, Tel2 and V6). |
| 2. | Changes after FMT (as compared to the change observed after one-week standard-of-care observation) and differences between patient groups based on the selected donors on the following aspects (all visits and/or telephone appointments, except for V1 and V4):<br>Severity of motor complications, i.e. number and duration of “off” periods* and periods with troublesome dyskinesias per day (3 days diary)<br>MDS-UPDRS (on medication)[52]<br>Required PD medication dose<br>Hoehn and Yahr score[55]<br>Q10 questionnaire (wearing off)[51]<br>MOCA[53]<br>Severity of GI symptoms and defecation frequency<br>Bristol stool scale<br>Other non-motor symptoms (SENS-PD[50]) |
| 3. | Ease of the study protocol, assessed by the reasons for refrainment of participation in the study after receiving full information at V1, and study load for participants, assessed by a 1-10 scale and open questions at V6. |
| 4. | FMT-related AEs in PD patients after FMT, assessed by the registration of FMT-related AEs at all post-allocation visits/telephone appointments. |
| **Other study parameters (at all visits and telephone appointments, except for V1 and V4)** |  |
|  | Sociodemographic factors<br>Diet<br>Health status<br>Disease characteristics |

\* ‘Off’ periods: a certain amount of time between regular doses of Parkinson medications, when the symptoms re-emerge or worsen.
Abbreviations: FMT: fecal microbiota transplantation, MDS-UPDRS: Movement Disorder Society-Sponsored Revision of the Unified
Parkinson’s Disease Rating Scale, MOCA: Montreal Cognitive Assessment, PD: parkinson’s disease, (S)AEs: (serious) adverse events,
SENS-PD: SEverity of Non-dopaminergic Symptoms in Parkinson’s Disease, Tel: telephone appointment, V: visit.

### Data collection and management

All PD patients will receive a pseudonymized study ID after signing the informed consent form. All clinical data and samples will be stored linked to this study ID. This study ID is linked to patient identifying data in a separate document, which will be securely stored on another password-protected location than the clinical research data. Completed patient questionnaires and diaries will be collected on paper and stored in a secured environment at the LUMC. These data and the results of the investigator examinations during visits or phone interviews and the blood analysis will be entered into a password protected cloud-based database at the LUMC (Castor) with real-time edit checks and automatic data saving. This database is only accessible for the study investigators, DSMB, monitors, and authorities for inspection of research. The raw 16S sequencing data of the stool samples will be stored in a folder with restricted access, and will anonymously be submitted to a public repository (European Nucleotide Archive). Data collection and storage and overall study procedures will be monitored by independent LUMC study monitors. Furthermore, independent GRP audits are regularly performed in the LUMC (https://www.lumc.nl/research/grp-and-integrity/grp/).

### Safety considerations

Prior to the start of the study, an independent DSMB will be assembled, consisting of two FMT experts (one gastroenterologist and one infectious disease specialist), one neurologist and a statistician. The DSMB will regularly meet to discuss all aspects of subject safety. The DSMB will perform an interim analysis when the first six patients have completed their six weeks post-FMT follow-up. The results will be disclosed to the investigators. In case of an SAE or on request of the investigator, the DSMB will be consulted to evaluate the relation with FMT and/or the potential need to terminate the study. The study will be terminated when there are definitely FMT-related SAEs in >1 patients at the interim analyses and/or when the subjects health or safety is jeopardized according to the DSMB, medical ethical committee and/or investigator. The principal investigator can also decide to withdraw a subject from the study for urgent medical reasons. Furthermore, patients are free to interrupt their participation in the study at any moment. The LUMC has a liability insurance and an insurance to cover health problems of participants caused by the study.

FMT is routinely performed in patients with multiple relapsing CDI, for whom it is considered a relatively safe procedure. A study performed by the NDFB on FMT-treated patients with recurrent CDI revealed that approximately 21-33% of patients report mild gastrointestinal adverse events, such as abdominal pain and diarrhea, in the three weeks after FMT and at long-term follow-up[36]. Among patients that receive FMT for other indications than CDI, the percentage that develop these gastrointestinal adverse events is unknown. In 0-2% of patients receiving FMT via upper GI route, SAEs are reported which are probably or definitely related to the FMT or to the procedure [35, 36]. Described SAEs that are possibly attributable to FMT or to the procedure via upper GI route include aspiration pneumonia, septicemia or other infections, fever, systemic inflammatory response syndrome, peritonitis, upper GI hemorrhage or death[34-37, 59, 60]. Long-term SAEs are largely unknown, although one recent study suggests FMT does not cause long-term SAEs.[61] No clinical trials have been performed with FMT in PD patients so far. In the available case series (total of 33 patients), one patient reported an SAE (episodes of vasovagal pre-syncope),[46] while mild transient side effects related to the procedure were reported in two series [45, 47]. The incidence and type of FMT- or procedure-related problems and (S)AEs in this group is unknown, and will be the main objective of this pilot study.

(S)AEs after FMT will be monitored very closely by measurement of hemoglobin, platelets, inflammation parameters, liver enzymes, kidney function and electrolytes before and after FMT and by the use of standard questionnaires. Furhermote, the patient will be instructed to always contact the investigators immediately by phone or e-mail in case of any SAE (by phone outside working hours). Participation in the study will be recorded in the electronic medical record of the LUMC and patients will receive a information card with study information and contact details, enabling other physicians to contact the investigators. All (S)AEs will be followed until they have abated, or until a stable situation has been reached, also after withdrawal. In case of an SAE, the investigators will report this as soon as possible to the Parkinson working group and the DSMB. In case of definitely FMT-related SAEs, the Parkinson working group will decide whether it may be useful to perform an autologous rescue FMT and/or provide antibiotics, as this may potentially reverse the donor FMT effect.

In this pilot study, the upper GI route will be used for FMT. Aspiration of donor fecal material in patients without PD resulting in fatal aspiration pneumonia has been described in only a few cases [34, 35, 62]. PD patients with severe swallowing problems or decreased GI motility will be excluded from the study. In addition, fecal suspensions will be injected slowly and the patient will be positioned in upright position to prevent regurgitation. Domperidone will be used to prevent and/or treat nausea and to improve gastric motility.

### Data visualization and analysis

For this study, both an intention-to-treat principle (ITT) and a per-protocol analysis will be conducted. Continuous variables will be summarized with means (with standard deviation) or medians (with interquartile range) and categorical variables with frequencies and percentages. If possible, ordinal outcomes on one subject will be summed. A two-tailed p<0.05 will be considered statistically significant. For linear mixed models, data will be converted into a logarithmic form in case of a skewed distribution. The investigators will attempt to prevent or minimize missing values by calling patients before every visit to remind them they need to fill in questionnaires and/or collect feces and also every filled-in questionnaire will be checked on completeness. Linear mixed models and generalized estimating equation (GEE) take missing values into account, when data are missing at random. When applicable, Bonferroni corrections will be applied. After analysis of study results, unblinding of donor selection will be performed. This pilot study focusses on feasibility and safety as primary outcome. This study is not powered for the secondary outcomes.

The assessment of FMT feasibility and safety, the ease of the study protocol and FMT-related AEs will be descriptive.

The bacterial fraction of the gut microbiota will be profiled via 16S rRNA gene amplicon sequencing. DNA will be extracted from 0.1 gram feces using the Quick-DNA™ Fecal/Soil Microbe Miniprep Kit (ZymoResearch, CA, USA). The V3-V4 or V4 region of the 16S rRNA gene will be sequenced on an Illumina platform. Raw sequencing data will be processed using a validated computational pipeline (NG-Tax[63], Qiime2[64]) using the Silva 132 SSU database for taxonomic classification[65]. 16S rRNA gene amplicon sequencing sequence data of the gut microbiota of donors and patients of before and at several time points after FMT will be assessed for FMT-dependent changes in gut microbiota composition. Sequence reads will be clustered on similarity (100%[66]) and assigned to the nearest bacterial phylum/family/genus and the relative abundance will be determined. Differences in bacterial diversity within and between samples will be evaluated by calculating the alpha- and beta-diversity of each sample. FMT-dependent changes will be defined as an alteration of alpha- or beta-diversity towards that of the donor and/or taxa abundances that become more similar to the donor microbiota after FMT. For gut microbiota analysis or continuous variables in the clinical data, outcomes post-FMT at several time points will be compared to pre-FMT data by linear mixed models (including one or (a mean of) all pre-FMT and one or all post-FMT measurements). Continuous variables may be converted into categorical variables. For categorical variables, generalized linear mixed models and/or GEE will be used (including one or all pre-FMT and one or all post-FMT measurements). A donor effect can be added, when applicable (or the Metagenomics Longitudinal Differential Abundance Method will be used).

The main outcome point is 1 week after FMT. All changes in clinical values and microbiota recorded at this time point with respect to baseline, will be compared with changes recorded at the standard-of-care visit (1 week after baseline).

### Ethical considerations

The study will be conducted according to the principles of the Declaration of Helsinki[67] and in accordance with the Medical Research Involving Human Subjects Act (WMO)[68]. This study was approved by the Medical Ethical Committee Leiden Den Haag Delft (ref. P20.087). Potential protocol amendments will be notified to this committee. The study protocol was discussed with two “patient researchers” of the Dutch Parkinson patients association to review the study load, safety and patient-centered value.

### Status and timeline

The study is registered in the Netherlands Trial Register (under trial registration number NL9438). The study has started in December 2021 and is expected to end in December 2022.

## Discussion

Since there are no curative treatments available for PD and most PD patients with advanced disease experience less effectivity and/or adverse effects of PD medication, the development of new treatment strategies is highly desirable. Animal studies suggest a potential role of the gut microbiota in PD pathophysiology via de gut-brain axis and in the metabolism of levodopa, the mainstay of PD treatment. The most extreme form of modifying the gut microbiota is replacing the existing dysbiotic gut microbiota with a new normal microbiota from healthy donors. So far, no results of clinical trials on FMT in PD patients have been published and a pilot study to assess the safety and feasibility of FMT in PD patients appears a logical next step. The results may also provide some preliminary information on the efficacy of FMT in decreasing motor- and non-motor symptoms and motor complications, useful to design future studies. Analysis of the gut microbiota composition will reveal preliminary data on associated key taxa of the gut microbiota in PD patients with motor complications. This pilot study has strengths and limitations. Strengths of this study are the use of two different donors, the broad range of clinical rating scales for Parkinson symptoms and constipation, the strict surveillance of (S)AEs, the inclusion of a standard-of-care measurement for comparison of the recorded changes, the analysis of the gut microbiota at different time points and the storage of an autologous suspension for treatment of a potential FMT-related SAE. Limitations include the absence of a comparator arm with placebo treatment, which was deemed too burdensome for the patients, considering the main focus on safety as outcome. In case FMT appears feasible and safe in this patient group, a larger double-blind randomized clinical trial may be performed to further explore the potential benefits of FMT.

The study results will be disseminated through publication in peer-reviewed journals and through presentations at international conferences. Authorship criteria are based on the International Committee of Medical Journal Editors (ICMJE).

## Data Availability

The raw 16S sequencing data will anonymously be submitted to a public repository (European Nucleotide Archive) when the study is completed.

## Acknowledgements

The authors are indebted to the following people: all the participants of this study for their support; the “patient researchers” of the Dutch Parkinson patients association for their previous advice on the study design; trial manager Linda van Hulst for her help with the preparation of the pre-treatment medication; a private donor who has partly financially supported the study.

## Supporting information

**S1 File. SPIRIT 2013 checklist: Recommended items to address in a clinical trial protocol and related documents**.

**S2 File. Approved study protocol**

**S3 File. FMT4PD study group**

