## Supplemental file S2 for "Safety and feasibility of fecal microbiota transplantation for Parkinson’s disease patients: a protocol for a self-controlled interventional donor-FMT pilot study"

**Supplementary information file S2. The approved study protocol**

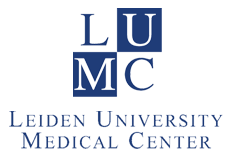

**RESEARCH PROTOCOL**

**Fecal Microbiota Transplantation for Parkinson’s Disease: a pilot study**

**(FMT4PD)**

(Version 3.0, 24-10-2020)

**Author**

K.E.W. Vendrik MD, MSc

Department of Medical Microbiology, Leiden University Medical Center

M.F. Contarino, MD, PhD

Department of Neurology, Leiden University Medical Center

**TABLE OF CONTENTS**

1.	INTRODUCTION AND RATIONALE	11

2.	OBJECTIVES	13

3.	STUDY DESIGN	13

4.	STUDY POPULATION	16

4.1	Population	16

4.2	Inclusion criteria	17

4.3	Exclusion criteria	17

4.4	Sample size calculation	17

5.	TREATMENT OF SUBJECTS	17

5.1	Investigational product/treatment	17

5.2	Use of co-intervention	18

5.3	Escape medication/procedures	18

6.	INVESTIGATIONAL PRODUCT/PROCEDURE	18

6.1	Name and description of investigational product(s)/procedure	18

6.2	Summary of findings from non-clinical studies	19

6.3	Summary of findings from clinical studies	24

6.4	Summary of known and potential risks and benefits	24

6.5	Description and justification of route of administration and dosage	24

6.6	Dosages, dosage modifications and method of administration	25

6.7	Preparation and labelling of Investigational Product	25

6.8	Drug accountability	26

7.	NON-INVESTIGATIONAL PRODUCT	26

7.1	Name and description of non-investigational product(s)	26

7.2	Summary of findings from non-clinical studies	26

7.3	Summary of findings from clinical studies	27

7.4	Summary of known and potential risks and benefits	27

7.5	Description and justification of route of administration and dosage	27

7.6	Dosages, dosage modifications and method of administration	27

7.7	Preparation and labelling of Non Investigational Medicinal Product	27

7.8	Drug accountability	28

8.	METHODS	28

8.1	Study parameters/endpoints	28

8.1.1	Main study parameters/endpoints	28

8.1.2	Secondary study parameters/endpoints	28

8.1.3	Other study parameters	28

8.2	Randomization, blinding and treatment allocation	28

8.3	Study procedures	29

8.4	Withdrawal of individual subjects	32

8.4.1	Specific criteria for withdrawal	32

8.5	Replacement of individual subjects after withdrawal	32

8.6	Follow-up of subjects withdrawn from treatment	32

8.7	Premature termination of the study	33

9.	SAFETY REPORTING	33

9.1	Temporary halt for reasons of subject safety	33

9.2	AEs, SAEs and SUSARs	33

9.2.1	Adverse events (AEs)	34

9.2.2	Serious adverse events (SAEs)	34

9.2.3	Suspected unexpected serious adverse reactions (SUSARs)	35

9.3	Annual safety report	35

9.4	Follow-up of adverse events	35

9.5	Data Safety Monitoring Board (DSMB)	35

10.	STATISTICAL ANALYSIS	36

10.1	Primary study parameters/endpoints	36

10.2	Secondary study parameters/endpoints	37

10.3	Other study parameters	38

10.4	Interim analysis	38

11.	ETHICAL CONSIDERATIONS	38

11.1	Regulation statement	38

11.2	Recruitment and consent	39

11.3	Objection by minors or incapacitated subjects	39

11.4	Benefits and risks assessment, group relatedness	39

11.5	Compensation for injury	40

11.6	Incentives	40

12.	ADMINISTRATIVE ASPECTS, MONITORING AND PUBLICATION	40

12.1	Handling and storage of data and documents	40

12.2	Monitoring and Quality Assurance	41

12.3	Amendments	41

12.4	Annual progress report	41

12.5	Temporary halt and (prematurely) end of study report	41

12.6	Public disclosure and publication policy	41

13.	STRUCTURED RISK ANALYSIS	42

13.1	Potential issues of concern	42

13.2	Synthesis	44

14.	REFERENCES	46

APPENDICES	52

Appendix A: Application form	52

Appendix B: FMT protocol of the NDFB	57

Appendix C: Product information of the fecal suspension for Fecal Microbiota Transplantation	60

Appendix D: Safe application of Faecal Microbiota Transplantation in the Netherlands …58

**LIST OF ABBREVIATIONS AND RELEVANT DEFINITIONS**

| **ABR** | **General Assessment and Registration form (ABR form), the application form that is required for submission to the accredited Ethics Committee; in Dutch: Algemeen Beoordelings- en Registratieformulier (ABR-formulier)** |
| --- | --- |
| **AE** | **Adverse Event** |
| **αSyn** | **alpha-synuclein** |
| **ASO** | **Parkinson’s disease mouse model with overexpression of αSyn** |
| **CDI** | ***Clostriodioides difficile* infections** |
| **CMAT** | **The Center for Microbiota Analyses and Therapeutics** |
| **CNS** | **Central nervous system** |
| **CV** | **Curriculum Vitae** |
| **DSMB** | **Data Safety Monitoring Board** |
| **ENS** | **Enteric nervous system** |
| **EudraCT**  **FMD** | **European drug regulatory affairs Clinical Trials**  **Fasting-mimicking diet** |
| **FMT** | **Fecal microbiota transplantation** |
| **GF** | **Germ-free** |
| **GI** | **Gastrointestinal** |
| **Hp** | ***Helicobacter pylori*** |
| **IC** | **Informed Consent** |
| **LUMC** | **Leiden University Medical Center** |
| **m** | **Month(s)** |
| **MDS-UPDRS** | **Movement Disorder Society-Sponsored Revision of the Uniﬁed Parkinson’s Disease Rating Scale** |
| **METC** | **Medical research ethics committee (MREC); in Dutch: medisch-ethische toetsingscommissie (METC)** |
| **mg** | **Milligram** |
| **ml** | **Milliliter** |
| **MOCA** | **Montreal Cognitive Assessment** |
| **MPTP** | **1-methyl-4-fenyl-1,2,3,6-tetrahydropyridine** |
| **NDFB** | **Netherlands Donor Feces Bank** |
| **PBS** | **Phosphate-buffered solution** |
| **PD** | **Parkinson’s disease** |
| **PI** | **Principal investigator** |
| **rCDI** | **Recurrent *Clostriodioides difficile* infections** |
| **(S)AE** | **(Serious) Adverse Event** |
| **SENS-PD** | **SEverity of Non-dopaminergic Symptoms in Parkinson’s Disease** |
| **SIBO** | **Small intestinal overgrowth** |
| **SIRS** | **Systemic inflammatory response syndrome** |
| **SPC** | **Summary of Product Characteristics; in Dutch: officiële productinformatie IB1-tekst** |
| **SPF** | **Specific-pathogen-free** |
| **Sponsor** | **The sponsor is the party that commissions the organisation or performance of the research, for example a pharmaceutical**  **company, academic hospital, scientific organisation or investigator. A party that provides funding for a study but does not commission it is not regarded as the sponsor, but referred to as a subsidising party.** |
| **SUSAR** | **Suspected Unexpected Serious Adverse Reaction** |
| **Tel** | **Telephone appointment** |
| **V** | **Visit** |
| **w** | **Week(s)** |
| **WMO** | **Medical Research Involving Human Subjects Act; in Dutch: Wet Medisch-wetenschappelijk Onderzoek met Mensen** |

**SUMMARY**

**Rationale:** The available literature suggests a role for the gut microbiota in the pathophysiology of Parkinson’s disease (PD). Changing the gut microbiota by means of fecal microbiota transplantation (FMT) could act on the pathophysiology of the disease and development of Levodopa-mediated motor complications in PD patients. In the proposed pilot study, FMT with feces from healthy donors will be performed for the first time in a study in PD patients. We hypothesize that FMT is feasible and safe in this patient group. In addition, we hypothesize that FMT will lead to a decrease of motor complications and PD symptoms in the short term, and an alteration of the intestinal microbiota composition towards that of the donor.
**Objective**:

*Primary objectives:*

1. Assess the feasibility of FMT in PD patients.
2. Assess the safety of FMT in PD patients.

*Secondary objectives:*

1. Explore whether FMT leads to alterations in motor complications (fluctuations or dyskinesias) and PD symptoms in the short term (up to three months post-FMT).
2. Determine alterations in gut microbiota composition and donor-recipient similarity, and their association with PD symptoms and motor complications.
3. Assess the ease of the study protocol.
4. Assess which FMT-related AEs are observed in PD patients after FMT

**Study design:** Single center prospective self-controlled interventional donor-FMT pilot study.

**Study population:** The study population will consist of 16 PD patients that use levodopa. Included PD patients should have idiopathic PD according to UK brain bank criteria with a disease duration of at least five years and should experience motor complications, despite using adequate PD medication. A written informed consent should be provided. Exclusion criteria are: Hoehn and Yahr scale stage 5, comorbidity or condition impairing ability to participate in the study according to the investigators, change in type or dose of PD medication in the previous three months, gastrointestinal (GI) infection or the use of antibiotics or probiotics in the previous three months, GI malignancy in the previous six months, known obstructions, paralysis or severe motility problems of the gastrointestinal tract, severe dysphagia with incapability of swallowing 2 liters of macrogol + electrolytes or inability to receive oral feeding, Inflammatory Bowel Disease, celiac disease, recent intraabdominal surgery(< 3 months) or intestinal resection in medical history, participation in another study within 16 weeks of screening visit, severe food allergy or allergy to medication that could be used by donors, (wish of) pregnancy, absence of contraception, lactation, immunocompromised state and use of immunosuppressants or opiates in the previous month. Patients should be able to understand and comply with study content and requirements, communicate in Dutch and be able to visit the Leiden University Medical Center (LUMC).

**Intervention**: FMT, with vancomycin and bowel lavage as pre-treatment and domperidone prior to FMT.

**Main study endpoints:**

1. Feasibility of FMT in PD patients: the number of included patients that cannot undergo FMT due to a patient- or procedure-related reason.
2. Safety of FMT in PD patients: FMT-related serious adverse events (SAEs).

**Nature and extent of the burden and risks associated with participation, benefit and group relatedness:** The participants will receive bowel lavage and antibiotics prior to FMT. They are not allowed to eat on the day of FMT prior to FMT. The FMT-procedure requires a gastroscopy to inject the fecal suspension directly into the horizontal duodenum or to insert a nasoduodenal tube with a pediatric gastroscope for later infusion of the fecal suspension, which are both minimally invasive procedures. The patient and the investigator or gastroenterologist can decide together which route is preferred. The nasoduodenal tube will remain in place until approximately 30 minutes after FMT. On the day of FMT, the patient will be in the hospital for approximately 2-4 hours. During this study, the patient has to visit the LUMC six times in total and will have two telephone appointments. Blood will be drawn three times. Physical examination, questionnaires, diary and collection of stool samples are repeated at each visit after screening (except for the FMT-visit).

FMT is a relatively safe procedure, but patients often experience mild self-limiting adverse events (AEs). The percentage of patients experiencing FMT-attributable AEs is 20-45%. In 0-5% of the patients, FMT-attributable SAEs are reported. The type and probability of specific procedure-related problems and (S)AEs in the group of PD patients is unknown. FMT in this pilot study will be performed via the upper GI route. Swallowing problems, delayed gastric emptying or decreased Gl motility may increase the risk of aspiration. However, we will exclude patients that cannot swallow 2 liters of laxatives. Importantly, nasoduodenal tube placement and nasoduodenal feeding are usually carried out without problems in PD patients.

The gut microbiota is considered to have a role in the pathophysiology of PD and in the metabolization of anti-PD medication. Based on previous studies, it is hypothesized that FMT with feces from healthy donors might improve the symptoms of PD, improve the effect of medication such as levodopa and limit their side effects, and/or slow down the disease progression. No studies have been performed with FMT in PD patients so far to confirm these findings. This study will provide crucial information about the safety and feasibility of this treatment in patients with PD, which, in the near future, could be further explored in larger trials aiming at determining the efficacy of FMT in PD patients. The participating patients will have the chance to experience this novel treatment and may possibly benefit from it.

A preliminary version of this study protocol was discussed with two Parkinson patients (patient-investigators), appointed by the Dutch Parkinson patients association (Parkinson vereniging), to review the study load, the safety and the patient-centered value of the study.

### 1. INTRODUCTION AND RATIONALE

PD is a progressive neurodegenerative disease that is characterized by the degeneration of neurons in the central nervous system (CNS), enteric nervous system (ENS) and peripheral autonomic nervous system, and the presence of Lewy bodies and Lewy neuritis in affected neurons^1^. An important factor in the etiology of PD may be the aggregation of the protein alpha-synuclein (αSyn), a major component of Lewy-bodies^2^. However, the etiology and pathogenesis of PD is still largely unknown. It is widely believed that there is a combination of genetic and environmental factors involved^3^.

GI symptoms (including obstipation and delayed transit) are frequently observed in PD patients and often precede the onset of motor symptoms, thus representing the first clinical manifestation of PD^4,5^. This suggests that the disease might be initiated in the gut. Concomitantly, several studies have demonstrated that alpha-synucleinopathy is present in the ENS and vagal nerves in an early phase of disease^6-11^. This led to the hypothesis that the disease may start in the gut, with a neurotrophic pathogen that is transported from the GI tract to the CNS by way of retrograde axonal and transneuronal transport through the vagal nerve.^6^ This neurotrophic pathogen might consist of misfolded αSyn molecular fragments^6,12^. The hypothesis is supported by studies suggesting that αSyn can spread from neuron to neuron^13^ and that αSyn forms could be transported from the gut to the brain^14-16^. It is further suggested that aggregation of αSyn in the brain and possibly the gut of PD patients is a consequence of inflammation‐induced oxidative stress^17-19^. Interestingly, PD patients have more inflammation of the colon, compared to healthy controls^20^. This finding suggests that there might be a role for peripheral inflammation in the initiation and/or the progression of PD.

The gut microbiota is the community of micro-organisms that resides in the gut. It has been hypothesized that the gut microbiota and their metabolites play an important role in the pathogenesis and course of PD. Several recent studies indicate that the gut microbiota and their metabolic products in PD patients are indeed different from healthy individuals^18,21-27^, although alpha-diversity (within-subject diversity) is similar to that of controls^23,26-28^. Other important findings are an overall more pro-inflammatory and less anti-inflammatory microbiota composition in PD patients^18^, with more genes involved in lipopolysaccharide biosynthesis^18^ and increased intestinal permeability^19^ compared to healthy controls. One study found that the increased relative abundance of *Enterobacterales* in PD patients was positively associated with the severity of postural instability and gait difficulty^23^. Two other studies suggested that gut bacterial tyrosine decarboxylases can metabolize levodopa to dopamine without being susceptible for aromatic amino acid decarboxylase inhibitors, such as carbidopa. Increased presence of gut bacterial tyrosine decarboxylases may thereby cause or worsen response fluctuations in levodopa/carbidopa-treated PD patients as dopamine cannot cross the blood-brain barrier^29,30^.

The prevalence of small intestinal bacterial overgrowth (SIBO) is increased in PD patients compared to healthy controls^31,32^ possibly due to a decreased GI motility in PD patients. SIBO is associated with impaired motor function and motor fluctuations^31-33^. Fasano *et al.*^32^ found that eradication of SIBO with rifaximin resulted in improvement of motor fluctuations, without affecting the pharmacokinetics of levodopa. By definition SIBO is associated with alterations of the gut microbiota. Furthermore, *Helicobacter pylori* (Hp) infections appear to be related to increased motor fluctuations in PD patients using Levodopa and treatment of Hp infections with antibiotics and omeprazole leads to improved motor fluctuations^34,35^. Pierantozzi et al^35^ observed increased levodopa absorption after Hp eradication therapy.

Probiotics may improve PD symptoms. One study showed an improvement in Movement Disorders Society Unified Parkinson Disease Rating Scale (MDS-UPDRS) score when PD medication was combined with probiotics. Other studies mainly observed alleviation of constipation^36^. All these studies underline a possible role of gut bacteria in the availability and/or absorption of PD medication.

A potential beneficial effect of FMT in PD patients is shown in several mouse studies ^37-39^. These are summarized in section 6.2 of this protocol. There is only one case report and one communication in a divulgative magazine describing the effect of FMT in PD patients with both showing improvement of PD symptoms after FMT ^40,41^. These are summarized in section 6.3 of this protocol.

FMT is a very effective treatment for recurrent (rCDI)^42-44^ and severe *Clostriodioides difficile* infections (severe CDI)^45^. At the moment, this is the only registered indication for FMT^46,47^. FMT is considered a safe treatment for patients with CDI^48^. Patients with CDI are shown to have a lower alpha-diversity of their microbiota^49,50^. FMT restores the reduced microbiota diversity and the disturbed metabolic capacity of the microbiota in these patients^51-53^. Data on other possible indications (e.g., hepatic encephalopathy, autism spectrum disorder and inflammatory bowel disease) are becoming available in experimental settings^54,55^.

The Netherlands Donor Feces Bank (NDFB), located in LUMC, provides ready-to-use quality assured fecal suspensions from healthy donors for FMT in patients with rCDI or severe CDI in the Netherlands. A total of 143 FMTs in 129 patients with recurrent or severe CDI were performed using a fecal suspension from the NDFB in the period May 2016 - August 2019 with a cure rate of 90% (manuscript in preparation).

Since there are no treatments available that cure PD or slow down the progression and most PD patients with advanced disease experience less effectivity and/or adverse effects of PD medication, the development of a new treatment strategy is crucial.

Changing the gut microbiota by means of an FMT could act on the pathophysiology of the disease and/or development of levodopa-mediated motor complications. Symptoms might decrease due to a direct effect of the changed gut microbiota on the gut-brain axis. They might be attenuated due to less production of pro-inflammatory cytokines with less intestinal inflammation and oxidative stress and subsequently less aggregation of αSyn in the ENS and CNS. Another important possibility is that FMT could lead to an increased absorption or less inhibition of PD medication in the gut due to the changed gut microbiota, resulting in an improved efficacy of the medication and less motor complications.

In the proposed pilot study, FMT will be performed with feces from healthy donors for the first time in a study in PD patients. Aim of the study is to demonstrate that FMT is feasible and safe in this patient group. In addition, we hypothesize that FMT will lead to a decrease of motor complications, PD symptoms in the short term and an alteration of the intestinal microbiota composition towards that of the donor and that the current study protocol is feasible and that the FMT-related AEs are comparable to what is found in other patient groups. In case FMT appears feasible and safe in this patient group, a future larger clinical trial may be performed to further explore the potential benefits of FMT.

### 2. OBJECTIVES

*Primary objectives:*

1. Assess the feasibility of FMT in PD patients.
2. Assess the safety of FMT in PD patients.

*Secondary objectives:*

1. Explore whether FMT leads to alterations in motor complications (fluctuations or dyskinesias) and PD symptoms in the short term (up to three months post-FMT).
2. Determine alterations in gut microbiota composition and donor-recipient similarity, and their association with PD symptoms and motor complications.
3. Assess the ease of the study protocol.
4. Assess which FMT-related AEs are observed in PD patients after FMT.

### 3. STUDY DESIGN

A single center prospective self-controlled interventional donor-FMT pilot study will be performed. Sixteen patients will be included. The follow-up period will be three months. The study site is LUMC. All FMTs will be performed at LUMC and the follow-up visits will also take place at LUMC. In figure 1 an overview of the study design is shown.

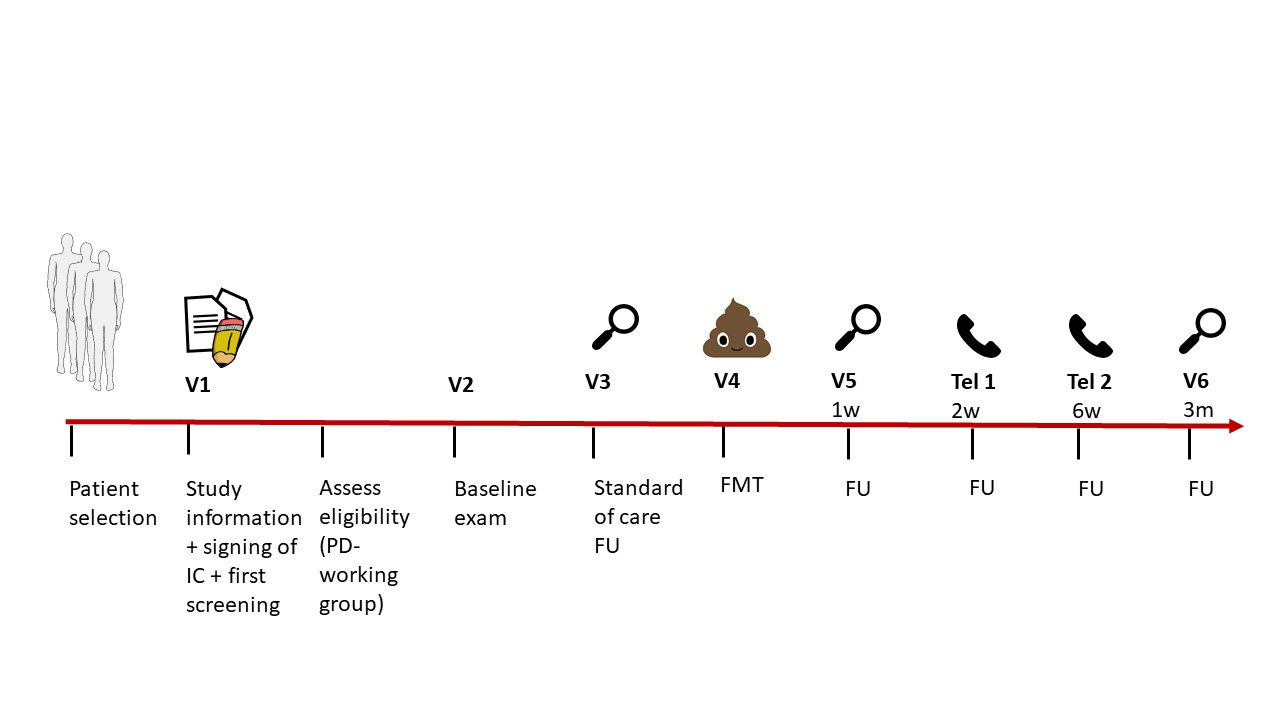

*Figure 1: Graphical abstract of study design. Abbreviations: FMT: fecal microbiota transplantation, FU: follow-up, IC: informed consent, PD: Parkinson’s disease.*

Description of the different steps in the study design

(all steps of the study and the various procedures to be performed are described in more detail in section 5, 6, 7 and 8 and in appendix B):

*Patient selection*

PD patients will be recruited in the first place from the LUMC, or if needed, PD patients will be searched by using advertisements. Selected patients will receive the patient information letter from the head of the LUMC Parkinson’s disease expertise center (a neurologist) from the LUMC (different from the principal investigator and data safety monitoring board member) with information about the study (including the informed consent form) and will be invited for visit 1 when interested.

*Visit 1: Information on the study, signing of informed consent and first screening*

During this visit the patient will be further informed on the study and questions can be asked. The investigators will determine whether the patient meets the inclusion and exclusion criteria and is able to participate in the study. In that case and if the patient is willing to participate, he/she will sign an informed consent form. Then, blood will be drawn to assess the baseline values and to assess whether there are comorbidities that may impair ability to participate in the study. When the patient needs additional time to consider participation in the study, the informed consent form can be signed during an extra visit at least one week later. This means that visit 1 will be postponed.

*Assess eligibility by Parkinson-working group*

The research physician and/or principal investigator (PI) will fill in an application form (Appendix A). This will be send to the Parkinson-working group, including several FMT-experts and one neurologist. They will evaluate the eligibility of the patient. Patients who are considered eligible will be included in the study and invited for visit 2.

*Visit 2: Baseline exam*

The baseline exam will be performed. The baseline exam includes:

- MDS-UPDRS IA, III and IV on medication
- Hoehn and Yahr
- SEverity of Non-dopaminergic Symptoms in Parkinson’s Disease (SENS-PD)
- Montreal Cognitive Assessment (MOCA)
- Baseline questionnaire
- Patient questionnaires
- Evaluation of the diary (3 days before visit until visit)
- Stool sample collection

*Visit 3: Follow-up standard of care one week after baseline exam*

The patients will be followed during one week of standard care before receiving an FMT with healthy donor feces. Visit 3 includes:

- MDS-UPDRS IA, III and IV on medication
- Hoehn and Yahr
- SENS-PD
- MOCA
- Patient questionnaires
- Evaluation of the diary (3 days before visit until visit)
- Registration of (S)AEs
- Stool sample collection

Visit 4: *FMT*

FMT via gastroscope or nasoduodenal tube with pre-treatment with vancomycin and macrogol + electrolytes (and bisacodyl in case of obstipation). On the day of FMT prior to FMT, the patient will receive one pill of domperidone. Problems during the FMT-procedure will be assessed.

*Visit 5: one week post-FMT exam*

- MDS-UPDRS IA, III and IV on medication
- Hoehn and Yahr
- SENS-PD
- MOCA
- Patient questionnaires
- Evaluation of the diary (3 days before visit until visit)
- Registration of (S)AEs
- Stool sample collection
- Blood sample collection

*Telephone appointment 1: two weeks post-FMT*

- Patient questionnaires
- Evaluation of the diary (3 days before telephone appointment until telephone appointment)
- MDS-UPDRS IA and IV
- Registration of (S)AEs

*Telephone appointment 2: six weeks post FMT*

- Patient questionnaires
- Evaluation of the diary (3 days before telephone appointment until telephone appointment)
- MDS-UPDRS IA and IV
- Registration of (S)AEs
- Stool sample collection

*Visit 6: three months post-FMT exam*

- MDS-UPDRS IA, III and IV on medication
- Hoehn and Yahr
- SENS-PD
- MOCA
- Patient questionnaires
- Evaluation of the diary (3 days before visit until visit)
- Registration of (S)AEs
- Stool sample collection
- Blood sample collection

*Table 1. Schedule of study procedures*

|  | **V1** | **V2** | **V3** | **V4** | **V5** | **Tel1** | **Tel2** | **V6** |
| --- | --- | --- | --- | --- | --- | --- | --- | --- |
| Time |  | Baseline | 1 w after baseline | FMT | 1 w after FMT | 2 w  After  FMT | 6 w after FMT | 3 m after FMT |
| Information on the study | X |  |  |  |  |  |  |  |
| Information on FMT | X |  |  |  |  |  |  |  |
| Signing informed consent | X |  |  |  |  |  |  |  |
| Screening | X |  |  |  |  |  |  |  |
| FMT |  |  |  | X |  |  |  |  |
| Patient questionnaires** |  | X | X |  | X | X | X | X |
| Diary (3 days before visit) |  | X | X |  | X | X | X | X |
| Baseline questionnaire* |  | X |  |  |  |  |  |  |
| MDS-UPDRS on medication*** |  | X | X |  | X | X | X | X |
| Hoehn and Yahr |  | X | X |  | X |  |  | X |
| SENS-PD |  | X | X |  | X |  |  | X |
| MOCA |  | X | X |  | X |  |  | X |
| Registration of (S)AEs |  |  | X | X | X | X | X | X |
| Stool sample |  | X | X |  | X |  | X | X |
| Blood sample | X |  |  |  | X |  |  | X |

* The baseline questionnaire includes questions on health status, disease-related variables and medication use (PD and non-PD).

**Patient questionnaires are filled in by the participant prior to a visit/telephone appointment and include questions on sociodemographic variables, health status, diet, constipation (Cleveland clinic constipation score^56^ and ROME IV criteria), SENS-PD, Q10 (wearing off), and MDS-UPDRS IB and II (and a study load questionnaire at V6).

*** MDS-UPDRS IA, III and IV (III not during telephone appointments).

Abbreviations: FMT: fecal microbiota transplantation, m: month(s), MDS-UPDRS: Movement Disorder Society-Sponsored Revision of the Uniﬁed Parkinson’s Disease Rating Scale, MOCA: Montreal Cognitive Assessment, (S)AEs: (serious) adverse events, SENS-PD: SEverity of Non-dopaminergic Symptoms in Parkinson’s Disease, Tel: telephone appointment, V: visit, w: week(s).

### 4. STUDY POPULATION

#### 4.1 Population

The study population will consist of 16 PD patients that are currently under treatment in the LUMC or PD patients that are recruited by advertisements on the website of the LUMC and/or of the Parkinson Association. We estimate that using this method one year is needed to find 16 eligible patients.

#### 4.2 Inclusion criteria

Clinical diagnosis of idiopathic PD according to UK brain bank criteria^57^.

PD disease duration of at least five years.

- Use of levodopa.
- Presence of motor complications (motor fluctuations or dyskinesias) despite adequate PD medication and regardless of severity.
- Written informed consent.

#### 4.3 Exclusion criteria

- Hoehn and Yahr scale stage 5 (most severe stage in scale for severity of PD motor symptoms).
- Comorbidity or condition impairing ability to participate in the study according to the investigators.
- Current use of probiotics or in the previous three months.
- Unstable PD with change in type or dose of PD medication in the previous three months.
- Symptoms of a GI infection during the previous three months.
- Current need of antibiotics or use in the previous three months.
- Current GI malignancy or in the previous six months.
- Known obstructions, paralysis or severe motility problems of the gastrointestinal tract
- Severe dysphagia with incapability of swallowing 2 liters of macrogol + electrolytes or inability to receive oral feeding.
- Known diagnosis of Inflammatory Bowel Disease (IBD)^58^ or celiac disease^59^.
- Intestinal resection in medical history.
- Recent intraabdominal surgery(< 3 months).
- Platelet count < 70x10^9^/L
- Participation in another study within 16 weeks of screening visit.
- Known severe food allergy or allergy to medication that a donor could have used (intake may lead to a life threatening situation).
- Immunocompromised state.
- Current use of immunosuppressants or opiates, or in the previous month.
- For women with child-bearing potential: Pregnancy; current wish to be pregnant or absence of contraception; lactation.
- Impaired ability to understand the study content and to give written informed consent.
- Unwilling or not capable to comply with the study requirements.
- Inability to communicate in Dutch.
- Inability to visit the LUMC.

#### 4.4 Sample size calculation

Since this is a pilot study, only 16 patients will be included. This is the number that is needed to have >80% chance that any FMT-related SAEs, that occur in >10% of the cases, might occur in the current study population. When FMT is performed in other diseases, SAEs definitely or probably related to FMT have been reported in 0-5% of the patients. The occurrence of FMT-related SAEs in >10% of the PD patients is deemed useful information that might change the design of a future randomized controlled clinical trial or might result in the choice not to perform such a clinical trial.

### 5. TREATMENT OF SUBJECTS

#### 5.1 Investigational product/treatment

Sixteen PD patients will be included, who will receive a donor FMT, randomized for feces of two healthy donors of the NDFB. Patients will receive the FMT via a gastroscope or nasoduodenal tube. The patients will be prepared for the FMT according to the standard protocol of the NDFB. This includes: preparation with 2 liters of laxatives (macrogol + electrolytes/Klean-prep) on the day prior to FMT, and vancomycin 250 milligram (mg) four times per day for five days pre-FMT until 24 hours before FMT. In case of obstipation, additional laxatives (Bisacodyl 2 times 5 mg ante noctem per day) will be administered in the two days prior to FMT to improve the efficacy of the bowel lavage. When this is not contraindicated, one pill of domperidone 10 mg will be self-administered orally on the day of FMT prior to FMT, to prevent nausea and to improve gastric motility. If the patient and/or physician (in case the patient agrees) prefer this, mild sedation by intravenous administration of midazolam before or during gastroscopy can be provided. More details are described in section 6 and 8 of this protocol.

#### 5.2 Use of co-intervention

Patients are not allowed to eat on the day of FMT prior to FMT. Female patients with child-bearing potential need to use adequate contraception during the study. There will be no other co-interventions during this study. PD patients are allowed to increase or decrease the dosage of medication or change the type of medication. This will be taken into account during analysis of the results.

#### 5.3 Escape medication/procedures

In case of nausea or vomiting after FMT, domperidone could be used, except when this is contraindicated.

In case of FMT-related SAEs, the Parkinson working group will decide whether it may be useful to perform an autologous rescue FMT and/or provide antibiotics, as this may potentially reverse the donor FMT effect. The Parkinson working group is a working group, which is assembled for this study and consists of two gastroenterologists, one infectious disease specialist, one medical microbiologist, and one neurologist (the PI). For the preparation of an autologous fecal suspension, before the baseline exam a stool sample should be delivered in a fecotainer to the NDFB within four hours after defecation. If PD patients are not able to bring the stool sample to the NDFB, the fecotainer with the stool will be picked up by an employee or student of the NDFB. This stool sample will be processed into an autologous fecal suspension for FMT (198 ml derived from 60 gram of feces), using methods described in standard operating procedures of the NDFB. The autologous fecal suspensions are stored in the freezer of the NDFB at -80°C.

### 6. INVESTIGATIONAL PRODUCT/PROCEDURE

#### 6.1 Name and description of investigational product(s)/procedure

A fecal microbiota suspension will be provided by the Dutch Donor Feces Bank (NDFB, housed at the LUMC). The NDFB is a non-profit stool bank for fecal transplantation with the primary aim of providing a standardized product for the treatment of patients with rCDI in the Netherlands. The NDFB participated in the development of international and European guidance documents for feces microbiota transplantation (FMT) and follows the recommendations issued therein.^60^

The working group of the NDFB consists of experts in the fields of microbiology, infectious diseases, gastroenterology, biobanking and methodology, and has extensive experience with FMT.^61,62^

The NDFB also supplies fecal suspensions for non-commercial research activities - provided that the scientific board agrees and all ethical permissions have been obtained - and participates in FMT-trials for ulcerative colitis, irritable bowel syndrome, non-alcoholic liver disease and eradication of multi-drug resistant organisms in kidney transplant patients.

FMT will be performed in all patients in this study. Defrosted ready-to-use fecal suspensions of 198 milliliters (ml), derived from feces of two healthy and rigorously screened donors, will be provided by the NDFB (<http://www.ndfb.nl/>). In the Netherlands, the fecal suspension is regarded as a transplant product and not as a medicinal product, food product or medical device.^61,63^ Fecal suspensions of two donors will be used in a randomized way. After exploring the existing literature, two donors are selected out of the donor pool of the NDFB, based on gut microbiota criteria that may be beneficial for PD patients. Two donors are selected to minimize the risk of no or a negative response due to donor specific characteristics and to get an idea on which donor gut microbiota characteristics are beneficial for PD patients. Importantly, it is unknown whether donors may develop PD in the future. However, donors with constipation are excluded and donors are asked whether there are any genetic diseases in the family. The NDFB decided not to ask specific questions to donors on risk factors for PD, such as decreased sense of smell, disturbed rapid eye movement sleep or family members with PD, since the knowledge of having an increased risk on PD may cause stress to the donors.

The used methods for donor screening are described by Terveer et al^61,62^ and in Appendix C. Under supervision of the Nederlandse Vereniging voor Medische Microbiologie (NVMM) and the Inspectie Gezondheidszorg en Jeugd the NDFB has drafted a guidance document for: “Safe application of Faecal microbiota Transplantation in the Netherlands” (Appendix D).

Feces donors of the NDFB are healthy individuals of between 18 and 60 years old that are rigorously screened via a questionnaire, interview, feces screening and blood screening. Donors do not have chronic diseases and do not use medication (except for sporadic use of some medication, like analgesics or antihistamines). Via the questionnaire and interview, the donors are screened on GI problems, diseases or characteristics associated with dysbiosis, risk behavior for infections, medical history, family history and medication use. The feces are tested every three months on (potential) virulent parasites, viruses (including the new coronavirus SARS-CoV-2) and bacteria (including multi-drug resistant organisms). Blood is tested every three months on sexual transmittable diseases or other via feces transmittable infections (including the new coronavirus SARS-CoV-2). The health of the donors is carefully monitored and feces and blood screening is repeated every three months, to test for new infections/colonization and to cover the window phase of some infections. All fecal suspensions are quarantined until a negative test result during re-screening and no development of new diseases between screening intervals. Donors are requested to contact the NDFB in case of a change in health or medication use and they will fill in a questionnaire at every donation with questions on their recent health status and risk factors for development of infections/colonization with (potential) pathogens (including multi-drug resistant organisms) or alterations in gut microbiota composition.

#### 6.2 Summary of findings from non-clinical studies

A summary of all FMT-studies in PD patients or PD animal models is provided in table 2 (for a complete overview see also Vendrik et al, Frontiers in Cell Infect Microbiol 2020^64^). Sampson *et al*.^65^ showed the importance of gut microbiota in the development of motor symptoms in a PD mouse model with overexpression of αSyn (ASO), concluding that gut bacteria are necessary to induce motor symptoms, alpha-synucleinopathy and neuro-inflammation. In this study, germ-free (GF) ASO mice showed less motor symptoms compared to specific-pathogen-free (SPF) ASO mice. When ASO mice received an FMT with feces from human PD patients, motor symptoms increased, compared to mice that received an FMT with feces from healthy human donors. The study clearly suggests that FMT with feces from healthy donors beneficially influences the course of PD. Meng-Fei Sun *et al*.^38^ used a 1-methyl-4-fenyl-1,2,3,6-tetrahydropyridine (MPTP)-induced PD mouse model and showed that mice that received a MPTP-injection had a better motor function after FMT with feces of healthy mice, compared to MPTP-injected mice that received no FMT. Furthermore, healthy mice that received feces from Parkinson mice performed worse compared to controls and in the traction test they performed even comparable to MPTP-injected mice. Zhou *et al*.^39^ observed less motor function decline and attenuated loss of dopaminergic neurons in the substantia nigra in PD mice that received a fasting-mimicking diet (FMD) compared to ad libitum-fed PD mice. Furthermore, they observed a higher (more favorable) striatal dopamine and serotonin concentration in PD mice that had received feces from FMD-fed control mice compared to phosphate-buffered solution (PBS)-gavaged or ad libitum microbiota-gavaged PD mice.

*Table 2. FMT in Parkinson’s disease*

| Study design | N | Follow-up after FMT | Neurological effects of FMT | GI effects of FMT | FMT-effects on microbiota | SAE after FMT (animals: other important effects) | Pre-treatment | Administration route | No of FMT | Amount of feces | Rationally selected feces donor | Year/  Reference |
| --- | --- | --- | --- | --- | --- | --- | --- | --- | --- | --- | --- | --- |
| Human  Case report | 1 | 3 m | UPDRS: decreased at 1 w after end of FMT-treatment, but became similar to pre-FMT at 3 m post-FMT.  Leg tremor almost disappeared at 1 w post-FMT but recurred in right lower extremity, more mild than pre-FMT, at 2 m post-FMT. | Wexner constipation score: decreased from 16 to 10.  PAC-QOL: decreased from 18 to 12 (8 at 1 w post-FMT).  Defecation time: Decreased from >30 to 5 min. | α-diversity: increased 1 w post-FMT, decreased after 3 m (OTU Number).  β-diversity: similar to donor at 1 w post-FMT, but similarity decreased later (w. UniFrac+PCoA).  Difference in individual taxa: yes. | No adverse effects | AB: NA  Bowel lavage: NA | TET tube, inserted into the ileocecal junction | 3 | 200 ml | No | 2019^40^ |
| Animal model:  Thy1-αSyn (ASO) mice  Relevant groups:  (all ASO or WT mice)  FMT:  1) GF+SPF-WT-FMT  2) GF+human PD-FMT  3) GF+human HC-FMT  No FMT:  4) GF  5) SPF  6) SPF+AB | 3-12 per group per analysis | 6-8 w (unclear for group 2 and 3) | Beam traversal, pole descent, adhesive removal, hindlimb clasping reflex score: ASO group 2 more motor symptoms vs ASO group 3. No effects in WT mice.  Beam traversal, pole descent, adhesive removal, hindlimb clasping reflex score: In ASO group 1 deterioration of motor symptoms and increased microglia cell body diameter, vs WT group 1 and 4. | No difference in constipation between group 2 and 3 in ASO or WT mice.  In ASO group 1 more constipation, vs WT group 1 and 5 and WT or ASO group 4 and 6. | α-diversity: NA.  β-diversity: most similar to donor,  mice with PD donors more similar to each other than to mice with HC donors. Difference between ASO and WT-mice post-FMT (w. en unw. UniFrac+ Bray-Curtis).  Difference in individual taxa: yes.  FMT with feces from SPF WT mice: NA. | NA | AB: NA  Bowel lavage: NA | Oral gavage | 1 | NA | Feces from 6 human PD patients, 6 human HCs or 3 SPF WT mice | 2016^65^ |
| Animal model:  MPTP-induced PD mice (i.p. injection)  Relevant groups:  (all SPF WT mice)  FMT:  1) MPTP+HC-FMT,  2) NS+PD-FMT,  3) NS+HC-FMT  No FMT:  4) No treatment,  5) MPTP+PBS 6) NS+PBS | 10-15 per group | 8 d after first FMT (until 1^st^ d after last treatment) | Worsened performance in pole descent and traction test and reduced striatal neurotransmitters in group 5 and 2 vs group 4, 6 and 3. Also improved (including no of dopaminergic neurons) in group 1 vs group 5.  Neuroinflammation: Decreased activated astrocytes and microglia in SN  and reduced expression of TLR4/TNF-α signaling pathway components  in gut and brain in group 1 vs group 5. | NA | α-diversity: Trend to increase in group 4 and little increase in group 1 vs 5 (Chao-1, phylog. div. whole tree).  β-diversity:  clustering of  group 1, group 4 and group 5 (w. UniFrac+ PCoA).  Difference in individual taxa: yes. | NA | AB: NA  Bowel lavage: NA | Gavage | 7 | 200 μL | Feces from normal control mice or MPTP-induced PD mice | 2018^38^ |
| Animal model:  MPTP-induced PD mice (i.p. injection)  Relevant groups:  FMT (AB-treated WT mice):  1) MPTP+AL-FMT  2) MPTP+FMD-FMT  No FMT (WT mice):  3) AB+MPTP+  PBS/G  4) AB+MPTP+  NF/HK | 8 per group | 8 d after first FMT (until 1^st^ d after last treatment) | Striatal DA and 5HT concentration of group 2 higher than group 1 and 3. 5HT concentration increased in group 1 compared with group 3.  5-HT concentration decreased in group 4, compared with group 2. | NA | NA | NA | AB: bacitracin gentamycin ciprofloxacin neomycin penicillin  Metronidazole  Ceftazidime  Vancomycin  streptomycin  Bowel lavage: NA | NA | 7 | 200 μL | Feces from normal mice treated with saline by intraperitoneal injection and fed ad libitum or fasting- mimicking diet | 2019^39^ |

Abbreviations: 5-HT: Serotonin or 5-hydroxytryptamine, AB: antibiotics, AB+MPTP+PBS/G: mice were treated with AB and MPTP intraperitoneal injection and 20% glycerol in sterile phosphate-buffered solution by gastric gavage, AB+MPTP+NF/HK: mice were treated with AB and MPTP intraperitoneal injection and heat-killed (HK) gut microbiota by gastric gavage from mice that were treated with normal saline by intraperitoneal injection and fasting-mimicking diet, ASO: alpha-synuclein overexpression, Chao1: estimates microbiota diversity from abundance data (measure of richness), DA: striatal dopamine, FMD: fasting-mimicking diet, fasting 3 days followed by 4 days of refeeding for three 1-week cycles, FMT: Fecal Microbiota Transplantation, GF: germ-free, GF+human HC-FMT: GF mice that receive FMT with feces from healthy controls, GF+human PD-FMT: GF mice that receive FMT with feces from human PD patients, GF+SPF-WT-FMT: GF mice that receive FMT with feces from SPF WT mice, GI: gastrointestinal, HC: healthy control, ns: non-significant, MPTP: 1-methyl-4-phenyl-1,2,3,6-tetrahydropyridine, MPTP+HC-FMT: mice that receive an MPTP injection i.p. and then an FMT with feces from normal control WT mice, MPTP+PBS: mice that receive MPTP i.p. and then PBS by gavage, NA: data not available, NS: normal saline, MPTP+AL-FMT: mice received MPTP and FMT with feces from mice that were treated with normal saline by intraperitoneal injection and were fed ad libitum, MPTP+FMD-FMT: mice that received MPTP and FMT with feces from mice that were treated with normal saline by intraperitoneal injection and fasting-mimicking diet, NS+HC-FMT: mice that received NS intraperitoneally and then an FMT with feces from normal control WT mice, NS+PBS: mice that received NS intraperitoneally and then PBS by gavage, NS+PD-FMT: mice that received NS intraperitoneally and then an FMT with feces from MPTP-mice, OTU: operational taxonomic unit, PAC-QOL: Patient Assessment of Constipation – Quality of Life, PBS: phosphate-buffered solution, PCoA: principal coordinates analysis, PD: Parkinson’s Disease, phylog. div: phylogenetic diversity, SAEs: serious adverse events, SCFA: short chain fatty acids, SPF: specific-pathogen-free, SPF+AB: SPF mice that receive antibiotics, F+SCFA: GF mice that receive oral SCFA, TET: Transendoscopic enteral tubing, Thy1-αSyn: alpha-synuclein-overexpression mouse model, unw: unweighted, UPDRS: Unified Parkinson’s Disease rating scale, w.: weighted, WT: wild-type

#### 6.4 Summary of findings from clinical studies

A summary of all FMT-studies in PD patients or PD animal models is provided in table 2. There is only one case report describing a PD patient that received FMTs in whom temporary improvement of leg tremors and other PD symptoms was observed one week after the third FMT^40^. However, leg tremors recurred at two months post-FMT and other PD symptoms had become similar to pre-FMT three months post-FMT. Constipation had also improved, which lasted until end of follow-up three months post-FMT. No adverse effects were observed. However, information on Parkinson symptom variability pre-FMT was missing. No further studies on FMT in PD were identified, except for one communication in a divulgative magazine in which improvement of PD symptoms after FMT was mentioned without further details^41^.

#### 6.5 Summary of known and potential risks and benefits

Benefits:

FMT is a very effective treatment for recurrent (rCDI)^42-44^ and severe *Clostriodioides difficile* infections (severe CDI)^45^, and for rCDI, cure rates of 80-95% are described in literature^42-44^. FMT restores the reduced microbiota diversity and the disturbed metabolic capacity of the microbiota in these patients^51-53^.

FMT may also be beneficial for several neurological indications where a role for the gut microbiota in disease pathogenesis is hypothesized (e.g., hepatic encephalopathy, autism spectrum disorder, multiple sclerosis). Publications on these indications are becoming available and FMT is currently being tested in larger populations^64,66^. Bajaj et al^54,67^ described reduced hospitalizations, improved cognition, and dysbiosis in patient with cirrhosis with recurrent hepatic encephalopathy after FMT from a rationally selected donor. In an open-label clinical trial of Kang et al^68,69^, gastrointestinal and behavioral ASD symptoms improved after FMT in 18 children with autism spectrum disorder and gastrointestinal symptoms, which persisted until two years after treatment. For other neurological indications the results of FMT are less clear^64^.

Potential benefits of FMT for PD been hypothesized based on data from previous studies (described in section 1). These include improvement of motor and non-motor symptoms (such as constipation), reduction of medication-induced motor complications, and ultimately slowing of disease progression. However, currently there is no published study yet demonstrating the benefit in PD patients.

Risks:

These are described in section 9.2 of this protocol.

#### 6.6 Description and justification of route of administration and dosage

The upper GI route is usually preferred by the NDFB over the lower GI route via colonoscope, because of the lower rate of SAEs^70^ (see also section 9.2). In this pilot study, the upper GI route will be used with infusion of the fecal suspension via gastroscope or nasoduodenal tube. Aspiration of donor fecal material resulting in a fatal aspiration pneumonia has been described, but is very rare (3 cases in the literature)^70-72^. PD patients may have swallowing problems, delayed gastric emptying and decreased GI motility, which may increase the risk on aspiration. However, nasoduodenal tube placement and nasoduodenal feeding are mostly carried out without problems in PD patients, which makes the likelihood of (S)AEs related to the infusion of a donor fecal suspension in these patients very low. In addition, particular attention to this aspect will be given when screening patients (see exclusion criteria) and the fecal suspension will be injected slowly at a rate of 10cc/min (approximately 1 hour after potential sedation) and in upright position of the patient to prevent regurgitation. In case of doubt on the position of a nasoduodenal tube, the position will be checked by X-ray. The alternative route for infusion of donor fecal suspensions is by colonoscopy. The burden for patients appears to be higher with this procedure. In addition, the amount of macrogol + electrolytes needed is higher (4 liters instead of 2 liters when performing FMT via upper GI). Higher amounts of macrogol + electrolytes could increase the risk on aspiration as well and, when there is inability to drink 4 liters of macrogol + electrolytes for the preparation of a colonoscopy, this might result in a less effective FMT. When this is not contraindicated, one pill of domperidone 10 mg will be self-administered orally on the day of FMT prior to FMT, to prevent nausea and to improve gastric motility. In case of nausea after FMT, domperidone could also be used.

#### 6.7 Dosages, dosage modifications and method of administration

The NDFB usually provides fecal suspensions of 198 ml, derived from 60 gram of healthy donor feces. Less than 50 gram of donor feces is proven to be less effective in literature^73,74^ and a surplus of feces increases the risk on regurgitation. The feces is diluted and filtered/sieved to facilitate the passing of the feces through the gastroscope or nasoduodenal tube. The patient and the investigator or gastroenterologist can decide together which administration route is preferred (e.g. dependent on the anatomy of the nose or stress of the patient). The fecal suspension is administered through a nasoduodenal tube (130 cm length and 3,3 mm diameter) or via a gastroscope. A nasoduodenal tube will be placed at the endoscopy department by the use of a pediatric gastroscope that is inserted through the nose. After this, the fecal suspension will be infused through the tube at the day care department. When infusion via nasoduodenal tube is preferred, standard treatment protocols for FMT via nasoduodenal tube of the NDFB are used (Appendix B). A nasoduodenal tube will remain in place until approximately 30 minutes after FMT. When the gastroscopy route is selected, the fecal suspension will be injected directly into the horizontal duodenum through a gastroscope at the endoscopy department.

#### 6.8 Preparation and labelling of Investigational Product

The NDFB follows the international and European guidelines^60^. Donor feces is collected using a Fecotainer to prevent environmental contamination and is processed to the end-product within 6 hours of defecation. The donor feces is processed to a ready-to-use fecal suspension with physiologic saline by homogenisation and sieving, allowing the suspension to pass the duodenal tube for clinical administration. Glycerol, in an end concentration of 10%, is added to allow optimal long-term storage at -80°C. Two RCTs and one meta-analysis showed non-inferiority and comparable cure rates for the treatment of rCDI with fresh or frozen fecal suspensions (stored at -80**°**C for up to 30 days)^75-77^.

Use of a frozen fecal suspension allows storage at -80**°**C for a longer period of time until the donor has been retested prior to actual use of the donor fecal suspension. This lowers the risk of transferring transmissible diseases by bypassing the window of detection phase of some transmissible infections (e.g. HIV, Hepatitis C). Storage duration at -80°C up to two years does not impact the clinical effectiveness of FMT for rCDI patients^78^. Fecal suspensions are therefore stored for a maximum of two years.

The fecal suspensions of 198 ml are stored with a unique, anonymized sample code in the centralized LUMC Biobank facility, which also participates in the national ‘Parelsnoer Institute’ (<https://www.health-ri.nl/parelsnoer>). A control sample of the original donor feces and an aliquot of the fecal suspension is stored separately from each processed and issued fecal suspension for biovigilance purposes to allow further investigations in the case of any complication. The issued fecal suspensions meet the pre-established quality criteria that have also been discussed in European context, tested and recorded in standard operation procedures.^60^

For more detailed information see <http://www.ndfb.nl/> and the publications by the NFDB ^61,62^

On the day of FMT the technician will transfer the fecal suspension into syringes of 50 ml, which only contain the study ID of the patient. Therefore, the physician that performs the FMT cannot see from which donor the fecal suspension is derived.

#### 6.9 Drug accountability

Fecal suspensions with the corresponding quality controls are stored in the LUMC Biobank in secured rooms. A technician of the NDFB will defrost the fecal suspension, transfer it to syringes and will add the study ID. The investigator will then take the fecal suspension to the patient. Data on which fecal suspension is administered, i.e. derived from which donor, the donation date, and the corresponding biobank-numbers, will be stored in a secured database, which is only accessibly to the persons that select the fecal suspensions for this study (and is not accessible for the investigators, the physician that performs the FMT or the research nurse).

### 7. NON-INVESTIGATIONAL PRODUCT

#### 7.1 Name and description of non-investigational product(s)

The patients will receive standard pre-treatment (with the same dosages) for FMT that the NDFB usually advices. Pre-FMT, the PD patients will receive vancomycin 250 mg four times per day orally for five days until 24 hours before FMT and bowel lavage by using two times 1 liter of macrogol + electrolytes (4 sachets of Klean-prep) on the day before FMT. Vancomycin is an antibiotic, that acts on Gram-positive bacteria and it belongs to the group of glycopeptides. It is a non-absorbable antibiotic (when taken orally). In case of FMT for rCDI, it is always used as pre-treatment. Macrogol + electrolytes is an osmotic laxative that is primarily used for constipation or bowel lavage as preparation for endoscopic procedures. In case of FMT, it is always used as pre-treatment. In case of obstipation, additional laxatives (bisacodyl two times 5 mg per day, both ante noctem) will be administered orally in the two days prior to FMT to improve the efficacy of the bowel lavage. When this is not contraindicated, one pill of domperidone 10 mg will be self-administered orally on the day of FMT prior to FMT, to prevent nausea and to improve gastric motility. In case of nausea after FMT, domperidone could also be used. Domperidone is a dopamine-antagonist that causes an increase in peristalsis of stomach and duodenum, an increase in pressure on the gastro-esophageal sphincter and relaxation of the sphincter of the pylorus. This leads to an increase in gastric emptying with prevention of vomiting. In contrast to several other antiemetics, this can safely be used in PD patients. Domperidon is frequently used in PD patients to prevent nausea, for example when starting new dopaminergic treatment.(e.g. domperidone is included as recommended adjuvant therapy in the brochure of apomorphine).

Furthermore, mild sedation by intravenous administration of 0.5–7,5 mg midazolam before or during gastroscopy can be provided, if the patient and/or physician (in case the patient agrees) prefers this. Only conscious sedation will be offered. Midazolam is a benzodiazepine that is frequently used to mildly sedate subjects during colonoscopy or gastroscopy.

Sedation and observation during and after sedation will be performed according to the LUMC protocol for sedation and analgesia in endoscopic procedures <http://iprova.lumc.nl/iDocument/Viewers/Frameworks/ViewDocument.aspx?DocumentID=0dda4f8d-1b57-4028-a235-202b19cd2fe6&NavigationHistoryID=22823639&PortalID=110&Query=sedatie+endoscopie>.

#### 7.2 Summary of findings from non-clinical studies

Patients with CDI that undergo an FMT usually receive antibiotics, mostly vancomycin, and bowel lavage prior to FMT.

In animal studies with PD mouse models, only one study mentioned pre-treatment with antibiotics, which included vancomycin but also other antibiotics^39^. This study showed a positive result of FMT on striatal DA and 5HT concentration. However, two other studies did not mention AB pre-treatment and also showed positive results^38,65^. No animal studies used bowel lavage prior to FMT. Furthermore, there is only one published case report in humans which the author did not mention the use of antbiotics or bowel lavage prior to FMT^40^.

For CDI, when FMT is given via upper GI, the main reason for the bowel lavage is primarily that the autologous microbiota is washed out, which may improve the engraftment of the donor fecal suspension. However, there is no data available that compares FMT with and without prior bowel lavage. Since there is mainly evidence that FMT with prior bowel lavage is effective^42-44^ and no data available on the effectiveness of FMT without prior bowel lavage, bowel lavage is almost always administered before FMT. Furthermore, the risks of bowel lavage are scarce.

In CDI patients, the reason for pre-treatment with antibiotics is principally to treat the CDI, since *C. difficile* is susceptible for vancomycin. Importantly, it also reduces overall bacterial load before FMT and is therefore thought to improve engraftment. For CDI, FMT serves mainly to prevent recurrences and therefore accompanying prior treatment with antibiotics is essential. It is not known whether antibiotics should be given prior to FMT for other indications. There is no data available that compares FMT with and without prior antibiotics in CDI or other indications. Results from studies in animals performed by Vedanta Biosciences in collaboration with the NDFB have shown that the administration of vancomycin before FMT leads to better engraftment of the donor fecal suspension with respect to no pre-treatment (unpublished data). Since the NDFB has a lot of experience with vancomycin pre-treatment and has shown good results with this, this pre-treatment was deemed the safest for PD patients.

Since PD patients have delayed gastric emptying and decreased GI motility and since transient and mild nausea is not a rare observation immediately after FMT^70,71^, domperidone is administered on the day of FMT prior to FMT when this is not contraindicated. This may prevent potential nausea and vomiting post-FMT.

Summary of Product Characteristics (SPC):

Vancomycin:

<https://www.geneesmiddeleninformatiebank.nl/smpc/h11984_smpc.pdf>

Macrogol + electrolytes:

<https://www.geneesmiddeleninformatiebank.nl/smpc/h15354_smpc.pdf>

Midazolam:

<https://www.geneesmiddeleninformatiebank.nl/smpc/h22594_smpc.pdf>

Domperidone:

<https://www.geneesmiddeleninformatiebank.nl/smpc/h07678_smpc.pdf>

#### 7.3 Summary of findings from clinical studies

*As described in section 7.2*

#### 7.4 Summary of known and potential risks and benefits

Risks are described in section 7.2

#### 7.5 Description and justification of route of administration and dosage

As described in section 7.1 and 7.2.

#### 7.6 Dosages, dosage modifications and method of administration

As described in section 7.1 and 7.2.

#### 7.7 Preparation and labelling of Non Investigational Medicinal Product

Preparation and labelling of the non-investigational medicinal products will be done according to the relevant GMP guidelines.

#### 7.8 Drug accountability

Drug accountability is not applicable for non-investigational products.

Vancomycin, macrogol + electrolytes and domperidone and, if used, bisacodyl, will be provided by the department of Clinical Pharmacy & Toxicology and will be self-administered by the patient. Midazolam, if used, will be stored at the department of Endoscopy and will be administered by a physician.

### 8. METHODS

#### 8.1 Study parameters/endpoints

##### 8.1.1 Main study parameters/endpoints

1. Feasibility of FMT in PD patients, assessed by the registration of the number of included patients that cannot undergo FMT due to a patient- or procedure-related reason.
2. Safety of FMT in PD patients, assessed by the registration of FMT-related SAEs.

##### 8.1.2 Secondary study parameters/endpoints

1. Alterations in gut microbiota structure (16S rRNA gene amplicon sequencing) after FMT, with comparison to the donor gut microbiota, and how these associate with PD symptoms and motor complications.
2. Changes after FMT (as compared to the change observed after one-week standard-of-care observation) and differences between patient groups based on the selected donors on the following aspects:

- Severity of motor complications, i.e. number and duration of off periods and periods with troublesome dyskinesias per day (3 days diary)
- MDS-UPDRS (on medication)
- Required PD medication dose
- Hoehn and Yahr score
- Q10 questionnaire (wearing off)
- Montreal Cognitive Assessment (MOCA)
- Severity of GI symptoms and defecation frequency
- Bristol stool scale
- Other non-motor symptoms (SENS-PD)

3. Ease of the study protocol, assessed by the reasons for refrainment of participation in the

study after receiving full information at V1, and study load for participants, assessed by a 1-10 scale and open questions.

4. FMT-related AEs in PD patients after FMT, assessed by the registration of FMT-related AEs.

##### 8.1.3 Other study parameters

- Sociodemographic factors
- Diet
- Health status
- Disease characteristics

#### 8.2 Randomization, blinding and treatment allocation

All subjects will receive an FMT. Before the FMT, there will be an observation period of standard of care, which means this pilot-study is self-controlled. Therefore, no subject randomization will be performed.

However, the donor selection will be randomized. Two healthy donors from the NDFB will be selected for this study, based on available literature. One or two employees of the NDFB that are not involved in this trial will use Castor to produce a randomization list for the two donors. This randomization list will not be disclosed to the investigators/physicians that are involved in this trial. The employees that produce the randomization list will inform the technician that prepares the fecal suspension to the patient. The syringes that contain the fecal suspension will not contain any donor identifying information (only the study ID).

Indications for breaking the randomization code could be infections (only when leading to SAEs that are probably or definitely related to FMT) in the PD patients post-FMT and when the Data safety monitoring Board (DSMB) deems it necessary. Then, a relation of the SAE with the selected donor feces could be examined.

#### 8.3 Study procedures

*Patient selection*

PD patients that are currently under treatment in the LUMC and who, based on the available clinical data, meet the inclusion and exclusion criteria will be selected. PD patients from the LUMC will be searched and selected in HIX by Parkinson nurses and the head of the LUMC Parkinson’s disease expertise center (a neurologist) by using CTcue. When this method does not provide enough patients, the study could be advertised on the website of the LUMC and of the Parkinson Association. Selected patients will receive the patient information letter from the head of the LUMC Parkinson’s disease expertise center (different from the PI and DSMB member) with information about the study (including the informed consent form) in a for the subject understandable language. Patients who are interested in taking part in the study will visit the neurology outpatients clinic of the LUMC for the first visit. During the first visit the patient will be further informed on the content of the study, the study load and the potential risks of FMT by on of the investigators and will have the chance to ask all the questions that may arise from reading the information letter. If the patient agrees with participating, the informed consent form will be signed in front of the investigator. If the patient needs additional time to consider participation in the study, the informed consent form can be signed during an extra visit at least one week later. This means that visit 1 will be postponed.We estimate that one year is needed to find 16 eligible patients.

*Independent FMT-expert*

When the patients have questions considering the participation in the study, they can contact an independent FMT-expert or an independent PD-expert. The independent experts will be selected before the start of the study and contact information will be mentioned in the informed consent form.

*Visit 1: Information on the study, signing of informed consent and first screening*

During this visit the patient will be further informed on the content of the study, the study load and the potential risks of FMT and will have the chance to ask all the questions that may arise from reading the information letter. The research physician and/or the PI will determine whether the patient meets the inclusion and exclusion criteria at that moment and is able to participate in the study. If necessary, the patient will also visit the gastroenterologist. The investigator will make sure that the patients receive complete, adequate written and oral information regarding the nature, aims, possible risks and benefits of the study. lt will be explained to the patients that they are free to interrupt their participation in the study at any moment without any consequences. If the patient is willing to participate and meets the inclusion and exclusion criteria, he/she will sign an informed consent form during visit 1. The investigator will make sure that the patients have a copy of the information sheet and informed consent form. The informed consent procedure will follow the SOP for informed consent of the LUMC (iProva). After signing of the informed consent, blood will be collected to assess the baseline values and to assess whether there are comorbidities that may impair ability to participate in the study.

In addition, patients will receive instructions concerning stool sample collection at home and the filling in of patient questionnaires and a diary during the study.

*Assess eligibility by Parkinson-working group*

When the patient is eligible according to the research physician and/or PI and the informed consent form is signed, the application form will be filled in (Appendix A). This will be send to the Parkinson-working group, including several FMT-experts and one neurologist (the PI). They will evaluate the eligibility of the patient (indication and possible contraindications).

Patients who are considered eligible will be invited for visit 2. These patients will receive feces collection material, patient questionnaires and a diary by post and they will be asked to fill in these patient questionnaires and the diary in the three days prior to visit 2. They will also be asked to bring a stool sample (all feces from one defecation) in a fecotainer to the NDFB within four hours after defecation before visit 2. If PD patients are not able to bring this stool sample to the NDFB, the stool will be picked up by an employee or student of the NDFB. This stool sample will be processed into an autologous rescue fecal suspension for FMT (further described in section 5.3).

*Baseline exam (V2)*

If the patient is eligible according to the Parkinson working group, the patient will visit the LUMC for visit 2 which includes the baseline exam.

The day before the baseline exam, the patients will fill in patient questionnaires on paper with questions on motor symptoms and non-motor symptoms from the week previous to the visit (Table 1). Patient questionnaires include questions on sociodemographic variables, health status, diet, constipation (Cleveland clinic constipation score^56^ and ROME IV criteria), ease of the study protocol, disease-related variables, medication use (PD and non-PD), SENS-PD, MDS-UPDRS IB and II, and Q10 (wearing off). In the three days before the baseline exam, the patient will fill in a diary on a daily basis to describe the motor complications during the day. During the baseline visit, the patients will hand over the filled-in diary and the patient questionnaires. The baseline questionnaire will be filled in by the investigators and/or a research nurse by asking questions to the patient about health status, disease-related variables and medication use (PD and non-PD). In addition, the MDS-UPDRS IA, III and IV, SENS-PD, MOCA and Hoehn and Yahr score will be assessed by asking questions to the patient and by physical examination. During visit 2, patients will be asked to fill in some patient questionnaires and a diary and to collect a stool sample in the three days before visit 3. Patients will be instructed to report all SAEs during the study immediately to the investigators.

*Follow-up standard of care (V3)*

To assess the variability of the study endpoints and to provide self-control data, a full evaluation will be performed 1 week after V2, following regular care. Diaries, patient questionnaires, MDS-UPDRS IA, III and IV, Hoehn and Yahr, MOCA and SENS-PD score assessment and the collection of a stool sample will be repeated (using feces collection tubs, as the fecotainer is only used for the first stool sample). Furthermore, the development of (S)AEs, using a standardized form, and changes in health and medication will be assessed (Table 1).

Information and instructions on the FMT procedure and the pre-treatment will be discussed again with the patient, including advantages and disadvantages of sedation before FMT. Vancomycin, macrogol + electrolytes and domperidone and, if used, bisacodyl, will be given to the patient with instructions, as these will be administered before visit 4.

*Fecal microbiota transplantation (V4)*

During visit 4, the FMT will be performed. The 16 selected patients will receive an FMT with healthy donor feces in the hospital (without overnight stay).

The FMT-procedure and the pre-treatment and medication that are used in this study are described in detail in section 6 and 7 of this protocol. After the FMT, the patient can go home when the observation period of at least two hours at the day-care department is over and the potential sedation has worn off. On the day of FMT, the patient will be in the hospital for approximately 2-4 hours. Furthermore, the development of (S)AEs will be assessed, using a standardized form. Patients will be asked to fill in some patient questionnaires and a diary and to collect a stool sample in the three days before visit 5.

*Follow-up after FMT (V5, V6, Tel1, Tel2)*

The post-FMT follow-up will be performed one week post-FMT, two weeks post-FMT, six weeks post-FMT and three month post-FMT. This includes two visits, at one week (V5) and three months post-FMT (V6), and two telephone appointments, at two weeks (Tel1) and six weeks (Tel 2) post-FMT.

During follow-up visits, the development of (S)AEs, the MDS-UPDRS IA, III and IV, SENS-PD, MOCA and Hoehn and Yahr score, and changes in health and medication will be assessed, blood will be drawn and patients will hand over the paper patient questionnaires, the diary and a stool sample (Table 1).This will be done by the investigators, supported by a research nurse. During each visit when blood is drawn, five tubes with in total approximately 30 ml of blood will be collected. Two tubes (approximately 7.5 ml) will be used for this study and three other tubes (approximately 22.5 ml) will be stored in the LUMC Biobank Parkinson for future research purposes. The regulations of the LUMC Biobank Neurologische Ziekten will be applicable.

At every contact the patient will be instructed to always contact the investigators in case of any SAE.

During the telephone appointments, the standardized (S)AE questionnaire and MDS-UPDRS IA and IV will be filled in by the investigators with the answers of the patient. The patient will also fill in the diary and the patient questionnaires before the telephone appointments and will bring them with them during visit 6. At six weeks post-FMT, the patient will also collect a stool sample, which will be sent to the LUMC by post.

During each visit/telephone appointment, the patient will be asked to fill in some patient questionnaires and a diary before the next visit/telephone appointment and, when applicable, to collect a stool sample in the three days before the next visit/telephone appointment. During the last visit, the study load will be assessed.

*Fecal sampling*

During this study, stool samples are collected for analysis and evaluation of the FMT treatment effect and (S)AEs (Table 1). The stool sample before the baseline exam will be used for the preparation of an autologous fecal suspension for an autologous FMT (described in section 5.3). The remainder of the stool sample will be stored for microbiota analysis and culturing purposes (preferably 4 gram) and for storage in the LUMC Biobank Parkinson (preferably 4 gram) and a part as safety aliquot of the fecal suspension (2 ml fecal suspension and 2 gram of the original stool sample) for when later analysis is needed in case of an (S)AE. If the baseline exam stool sample does not contain at least 33 gram, the patient will be asked to collect another stool sample. For the remaining stool samples during this study, at least 2 gram is required. The one week post-baseline exam, one week post-FMT and three months post-FMT stool samples will be handed over at the visits. Patients will be requested to collect stool samples of each defecation from three days before the visit, or earlier if the patient has severe constipation, until the visit, and to store it in the fridge and bring the most fresh stool sample during the visit for optimal quality of the feces. At six weeks post-FMT, patients will be requested to send a stool sample by post, preferably as soon as possible after defecation, or if not possible, with storage in the fridge until transport.

Every stool sample will be analysed by the patient using the Bristol stool scale to describe the consistency of the feces before and after FMT. Microbiota analysis will be performed on all stool samples, and culturing when deemed necessary, to assess the changes in the microbiota composition and diversity after FMT. Additional analysis on the stool samples will be performed, when the Parkinson Working Group or data safety monitoring board deems this necessary for safety reasons, e.g. due to an (S)AE.

All fecal suspensions (autologous and of the healthy donors) are stored in a -80°C freezer of the NDFB or biobank. All stool samples will be aliquoted and stored in a -80°C freezer of the NDFB and in the LUMC Biobank Parkinson. When possible, at least two times 1 gram feces is stored with 10% glycerol as cryoprotectant (for culturing purposes) and at least two times 1 gram feces is stored without glycerol (for microbiota analysis by 16S rRNA gene amplicon sequencing) in the NDFB freezer. In addition, when there is more feces left and if participants give permission for LUMC Biobank Parkinson storage, two aliquots of 1 gram with 10% glycerol and two aliquots of 1 gram without glycerol will be stored in the LUMC Biobank Parkinson for future research purposes. The regulations of the LUMC Biobank Neurologische Ziekten will be applicable.

The bacterial fraction of the gut microbiota will be profiled via 16S rRNA gene amplicon sequencing, giving insights in the gut microbiota’s structural features, including its composition, diversity and bacterial networks, which can be associated with clinical data. In addition, stored stool samples can be retrieved whenever needed for further microbiome analyses of interest (metagenomics, metatranscriptomics, metaproteomics and metabolomics).

To assess the fecal microbiota, DNA will be extracted from 0.1 gram feces using the Quick-DNA™ Fecal/Soil Microbe Miniprep Kit (ZymoResearch, CA, USA). The V3-V4 or V4 region of the 16S rRNA gene will be sequenced on an Illumina platform (in paired-end modus, 150-300 bp). Raw sequencing data will be processed using a validated computational pipeline (NG-Tax, Qiime2) using the Silva 132 SSU database for taxonomic classification.

*Blood sampling*

Blood will be drawn at the screening visit, at one week post-FMT and three months post-FMT, to assess whether there is comorbidity that may impair the ability to participate in the study and to asssess alterations in hemoglobin, platelets, inflammation parameters, liver enzymes, kidney function and electrolytes after FMT for safety reasons. During each visit, five tubes with in total approximately 30 ml of blood will be collected. This will consist of two tubes for blood and serum analysis of in total approximately 7.5 ml and, if participants give permission for the LUMC Biobank Parkinson, also three tubes of in total approximately 22.5 ml for the LUMC Biobank Parkinson. If participants give permission for the LUMC Biobank Parkinson, their blood samples (and some DNA from the blood) will be stored for future (yet unknown) analysis.

#### 8.4 Withdrawal of individual subjects

Subjects can leave the study at any time for any reason if they wish to do so without any consequences. The investigator can decide to withdraw a subject from the study for urgent medical reasons.

##### 8.4.1 Specific criteria for withdrawal

Subjects are excluded during the study when they develop a contraindication for FMT prior to FMT.

#### 8.5 Replacement of individual subjects after withdrawal

When there is a withdrawal before the FMT, individual subjects will be replaced. When the patient is not willing/able to participate in the follow-up visits and telephone appointments after FMT, individual subjects will not be replaced. Subjects will be analysed in an intention to treat analysis and per protocol analysis.

#### 8.6 Follow-up of subjects withdrawn from treatment

Individual subjects that are withdrawn from the study after receiving FMT, will be periodically contacted by telephone to assess the development of (S)AEs. All AEs and SAEs will be followed until they have abated, or until a stable situation has been reached. Depending on the event, follow up may require additional tests or medical procedures as indicated, and/or referral to the general physician or a medical specialist.

#### 8.7 Premature termination of the study

In accordance to section 10, subsection 4, of the WMO, the sponsor will suspend the study if there is sufficient ground that continuation of the study will jeopardise subject health or safety. The sponsor will notify the accredited medical research ethics committee (METC) and the data safety monitoring board (DSMB) without undue delay of a temporary halt including the reason for such an action. The study will be suspended pending a further positive decision by the accredited METC. The investigators will take care that all subjects are kept informed. The METC, DSMB and/or the sponsor will decide whether the study should be terminated prematurely.

Furthermore, the DSMB will assess the potential need to terminate the study after the interim analysis and in case of an SAE or on request of the sponsor (described in section 9.5). When the study is terminated, no further FMT-procedures will be performed.

### 9. SAFETY REPORTING

#### 9.1 Temporary halt for reasons of subject safety

In accordance to section 10, subsection 4, of the WMO, the sponsor will suspend the study if there is sufficient ground that continuation of the study will jeopardise subject health or safety. The sponsor will notify the accredited METC and the DSMB without undue delay of a temporary halt including the reason for such an action. The study will be suspended pending a further positive decision by the accredited METC. The investigators will take care that all subjects are kept informed.

#### 9.2 AEs, SAEs and SUSARs

FMT is considered the preferred treatment for patients with multiple relapsing CDI. In this population, it is considered a relatively safe procedure, but (S)AEs have been described.

No studies have been performed with FMT in PD patients so far. The type and probability of specific procedure-related problems and (S)AEs in this group is unknown, and will be the main objective of this pilot study.

To assess the safety of FMT in PD patients, (S)AEs after FMT will be monitored very closely and hemoglobin, platelets, inflammation parameters, liver enzymes, kidney function and electrolytes will be assessed before and after FMT.

From every patient that receives a donor FMT, a ready-to-use autologous rescue fecal suspension will be prepared prior to FMT and stored in a -80°C freezer. In case of FMT-related SAEs, the Parkinson working group will decide whether it may be useful to perform an autologous rescue FMT and/or provide antibiotics, as this may potentially reverse the donor FMT effect (described in section 5.3).

During the follow-up visits, the patients will be questioned on (S)AEs. Before the FMT, the patient will also be instructed to always contact the investigators immediately in case of any SAE. The investigators will report all (S)AEs in the medical records and case report forms of the patient and in the (S)AE register. For each (S)AE the following details will be recorded in the (S)AE register:

1. SAE or AE

2. Description

3. Date and time of occurrence

4. Duration

5. Relationship with the intervention

6. Action taken

7. Outcome

Participation in the study will be recorded in the electronic patient file of the LUMC, which means that physicians will receive information on the study with contact details of the PI and research physician, when the patient is admitted to the LUMC or in case of an outpatient visit in the LUMC. In addition, the patient will receive a card with information on the study and contact details to be used in case of emergency which needs to be provided to physicians in case of an admission or outpatient visit in another hospital than the LUMC. In case of an SAE, physicians are requested to report this within three days to the investigators. The investigators will report SAEs in the medical records of the patient, case report form of the patient and the (S)AE register. The investigators will report this as soon as possible to the Parkinson working group and the DSMB. The DSMB will assess whether it could be related to the FMT and whether the study should be paused or terminated prematurely. In case of a clinically relevant increase or decrease in certain blood values after FMT or in case of doubt on the clinical relevance of blood values, the investigators will report this to the DSMB. They decide whether it is an SAE and whether it is FMT-related. All not serious AEs will be communicated to the investigators within seven days. When the AE was not expected, the investigators will discuss this with the Parkinson Working group.

The investigators will report an SAEs through the web portal *ToetsingOnline* to the accredited METC that approved the protocol, within seven days of first knowledge for SAEs that result in death or are life threatening followed by a period of maximum of eight days to complete the initial preliminary report. All other SAEs will be reported within a period of maximum 15 days after the investigators have first knowledge of the SAEs.

A member of the NDFB is always available for consultation in case of any possibly FMT-related (S)AEs or possible FMT-related problems.

The medical advisory board of the NDFB will be informed regularly on the progress of the study.

##### 9.2.1 Adverse events (AEs)

AEs are defined as any undesirable experience occurring to a subject during the study, whether or not considered related to FMT. All AEs reported spontaneously by the subject or observed by the investigators or his staff will be recorded.

During standard FMT procedures, patients can experience mild self-limiting AEs of the GI tract shortly after FMT. The percentage of patients experiencing FMT-attributable AEs varies among studies. They occur in approximately 20-45% of the patients. In literature and in a large cohort of 130 patients treated by the NDFB, the most common AEs when performing FMT via the upper GI route are abdominal discomfort (including abdominal pain), increased stool frequency, flatulence, bloating and cramps. AEs due to upper GI endoscopy include nasal stuffiness, sore throat and rhinorrhea ^48,70-72,79^. All these AEs are often mild and transient.

##### 9.2.2 Serious adverse events (SAEs)

An SAE is any untoward medical occurrence or effect that:

- results in death;
- is life threatening (at the time of the event);
- requires hospitalisation or prolongation of existing inpatients’ hospitalisation;
- results in persistent or significant disability or incapacity;
- is a congenital anomaly or birth defect; or
- any other important medical event that did not result in any of the outcomes listed above due to medical or surgical intervention but could have been based upon appropriate judgement by the investigators.

An elective hospital admission will not be considered as an SAE.

In 0-5% of patients receiving FMT, SAEs are reported which are probably or definitely related to the FMT or to the procedure ^70-72^. In the systemic review of Wang et al^70^, SAEs were found in 2.0% of the patients that received FMT via the upper GI routes and 6.1% of the patients that received FMT via lower GI routes. FMT via lower GI route gives an increased risk on perforation, GI hemorrhage and sedation-associated risks^48,70-72,79^. Therefore, FMT will be performed via the upper GI route in the current study. Described SAEs that are possibly attributable to FMT or to the procedure via upper GI route include aspiration pneumonia, septicemia or other infections, fever, peritonitis, upper GI hemorrhage or death^48,70-72,79^. One study showed a transient increase of neutrophils, decreased lymphocytes and increased CD3+/CD4+ and CD4+/CD8+ ratios in three healthy subjects that received capsules with feces from healthy donors, but these effects were mostly transient^80^. One patient developed a systemic inflammatory response syndrome (SIRS). These results suggest a transient systemic acute response to antigenic exposure with leukocytosis.

All these SAEs due to FMT are uncommon. In the systematic review of Wang et al^70^ one death related to FMT was described in 1089 patients (0.09%), caused by aspiration during sedation of colonoscopy^81^. The other 37 deaths after FMT were possibly or unrelated to FMT. Another review by Baxter et al^71^ found a death rate that was potentially attributable to FMT of 0.3% (3/1174 patients), due to polymicrobial septic shock with decompensated toxic megacolon in a patient that received FMT via gastric tube^82^, aspiration during sedation to deliver a colonoscopic FMT^81^ (same case as the above mentioned by Wang et al^70^) and aspiration pneumonia with septic shock after an upper-GI FMT under general anesthesia^83^. Beurden et al^72^ reviewed 39 FMTs via nasoduodenal tube performed in the Amsterdam Medical Center in the Netherlands and reported one patient that died (1/39), due to pneumonia, possibly caused by aspiration.

##### 9.2.3 Suspected unexpected serious adverse reactions (SUSARs)

Not applicable, since FMT is not considered a medicine.

#### 9.3 Annual safety report

Not applicable, since FMT is not considered a medicine.

#### 9.4 Follow-up of adverse events

All AEs and SAEs will be followed until they have abated, or until a stable situation has been reached. Depending on the event, follow up may require additional tests or medical procedures as indicated, and/or referral to the general physician or a medical specialist. AEs and SAEs will be reported in the case report form of the patient and the (S)AE register till end of the study.

#### 9.5 Data Safety Monitoring Board (DSMB)

Prior to the start of the study, a DSMB will be assembled. The DSMB will consist of at least two independent FMT-experts (one gastroenterologist and one infectious disease specialist), one independent neurologist (different from the previous mentioned independent FMT-expert and neurologist) and an independent statistician.

An open progress meeting with the DSMB will be held at the start of the study, at least once a year during the study and at the end of the study, in which the DSMB monitors recruitment figures and losses to follow-up, evidence for treatment harm, compliance with previous DMC recommendations, the need for termination of the trial and breaking of the randomization code of donor selection, the need for additional data analyses, and advises on protocol modifications suggested by investigators and assesses the impact and relevance of external evidence. A closed DSMB meeting will be planned when the first six patients have had their six weeks post-FMT follow-up (safety interim analysis). Additional DSMB meetings will be organized in case of an SAE and on request of the sponsor to evaluate the relation with FMT and/or the potential need to terminate the study. This will be open or closed, dependent on the wish of the DSMB.

The DSMB will support the Parkinson working group with an interim analyses on the 6 weeks post-FMT follow-up when the first six patients have completed their six weeks post-FMT follow-up to monitor safety. In case of an SAE and on request of the sponsor they will also be consulted shortly after an event (at least within 2 weeks) to evaluate the relation with FMT and/or the potential need to terminate the study. The study will be terminated when there are definitely FMT-related SAEs in >1 patients at the interim analyses at six weeks post-FMT in the first six patients or when there is another reason for premature termination of the study according to the DSMB. The advice(s) of the DSMB will only be sent to the sponsor of the study (with a copy to the coordinating research physician). Should the sponsor decide not to fully implement the advice of the DSMB, the sponsor will send the advice to the reviewing METC, including a note to substantiate why (part of) the advice of the DSMB will not be followed.

The DSMB members have no conflict of interest, as they are not involved in the design or execution of this study (except for the statistician, who will only provide advice on statistics in this study) and they have no financial relation with the NDFB or other parties involved in this study.

### 10. STATISTICAL ANALYSIS

General considerations:

This pilot study focusses on feasibility and safety as primary outcome. The sample size is low and this study is not powered for the secondary outcomes, which means that the statistics will be less reliable for these. In case FMT appears feasible and safe in this patient group, a future larger clinical trial may be performed to further explore the potential benefits of FMT.

For this study, both an intention-to-treat principle (ITT) and a per-protocol analysis will be conducted. Since the secondary outcomes aim at exploring any effect of FMT, no formal hypothesis testing will be performed.

ln general, continuous variables will be summarized with standard descriptive statistics including means (with standard deviation) or medians (with interquartile range). Categorical variables will be summarized with frequencies and percentages. Ninety-five percent confidence intervals or interquartile ranges will be provided for descriptive statistics, dependent on whether there is a skewed distribution. If possible, ordinal outcomes on one subject will be summed (e.g. all questions on depression in questionnaires). These outcomes can be considered numeric variables and in this way the power can be increased.

After analysis of study results, unblinding of donor selection will be performed.

#### 10.1 Primary study parameters/endpoints

1. Feasibility of FMT in PD patients, assessed by the registration of the number of included patients that cannot undergo FMT due to a patient- or procedure-related reason.

This will be descriptive and will be assessed by the registration of the number of included patients that cannot undergo FMT. All reasons for rescheduling or aborting a FMT will be recorded. In case of >20% of patients (>3 patients) that cannot undergo FMT due to a patient- or procedure-related reason, the FMT-procedure is considered not feasible.

2. Safety of FMT in PD patients, assessed by the registration of FMT-related SAEs.

The nature and number of SAEs and the relation with FMT will be described. This also includes alterations in hemoglobin, platelets, inflammation parameters, liver enzymes, kidney function and electrolytes after FMT, indicative of FMT-related SAEs. An FMT will be considered unsafe in PD patients, i.e. a larger phase 2 clinical trial with the same procedure will not be recommended, when there are definitely FMT-related SAEs in >10% of the cases, i.e. >1 patients, at the end of the study.

#### 10.2 Secondary study parameters/endpoints

1. Alterations in gut microbiota structure (16S rRNA gene amplicon sequencing) after FMT, with comparison to the donor gut microbiota, and how these associate with PD symptoms and motor complications.

Microbiota analyses will be performed by the Center for Microbiota Analyses and Therapeutics (CMAT), that is well equipped to study dysbiosis, gut microbiota composition and its alterations after FMT.

Statistical analyses and data visualization will be performed in R using packages phyloseq, vegan, ggplot2, DESeq2 and Microbiome, among others. 16S rRNA gene amplicon sequencing sequence data of donor gut microbiota and gut microbiota of the patients of before and several timepoints after FMT will be assessed for FMT-dependent changes in gut microbiota composition and engraftment of donor bacteria. Sequence reads will be clustered on similarity (97-100%) and assigned to the nearest bacterial phylum/family/genus and the relative abundance will be determined. Differences in bacterial diversity within and between samples will be evaluated by calculating the alpha- and beta-diversity of each sample. FMT-dependent changes will be defined as an alteration of alpha- or beta-diversity towards that of the donor and/or taxa abundances that become more similar to the donor microbiota after FMT. Engraftment of donor bacteria will be assessed by: beta-diversity (similarity/distance measure between donor-recipient microbiota) and by assessing the percentage of taxa (from total of taxa) post-FMT that are derived from the donor, are inherent to the patient (based on pre-FMT sample), and are shared (based on pre-FMT and donor sample) (with the assumption that the bacteria are not acquired from the environment). A minimum threshold of 0.1 % relative species abundance will be used in determining engraftment. Outcomes post-FMT at several timepoints will be compared to pre-FMT data by linear mixed models when normally distributed and data will be converted into logarithmic form in case of a skewed distribution. A two-tailed p<0.05 will be considered statistically significant. When applicable, Bonferroni corrections will be applied to correct for multiple testing. Linear mixed models takes missing values into account, when the data is missing at random. The investigators will attempt to prevent missing values or, if not possible, to minimize the amount of missing values. For outcomes that are considered to have the potential to be different between the patient group that received feces from one donor and the patient group that received feces from another donor, a donor effect will be added to the linear mixed models (or a MetaLonDA analysis will be performed: Metagenomics Longitudinal Differential Abundance Method).

2. Changes after FMT (as compared to the change observed after one-week standard-of-care observation) and differences between patient groups based on the selected donors on the following aspects:

- Severity of motor complications, i.e. number and duration of off periods and periods with troublesome dyskinesias per day (3 days diary)
- MDS-UPDRS (on medication)
- Required PD medication dose
- Hoehn and Yahr score
- Q10 questionnaire (wearing off)
- Montreal Cognitive Assessment (MOCA)
- Severity of GI symptoms and defecation frequency
- Bristol stool scale
- Other non-motor symptoms (SENS-PD)

For continuous variables, outcomes post-FMT at several timepoints will be compared to pre-FMT data by linear mixed models when normally distributed and data will be converted into logarithmic form in case of a skewed distribution. For categorical variables, generalized linear mixed models will be used. A two-tailed p<0.05 will be considered statistically significant. When applicable, Bonferroni corrections will be applied to correct for multiple testing. Linear mixed models takes missing values into account, when the data is missing at random. The investigators will attempt to prevent missing values or, if not possible, to minimize the amount of missing values. For outcomes that are considered to have the potential to be different between the patient group that received feces from one donor and the patient group that received feces from the other group, a donor effect will be added to the (generalized) linear mixed models.

3. Ease of the study protocol, assessed by the reasons for refrainment of participation in the

study after receiving full information at V1, and study load for participants, assessed by a 1-10 scale and open questions.

This will be descriptive. PD patients that do not want to participate in the study after receiving information at V1 are asked why not and at the end of the follow-up, participants will be asked to rate the study load (scale from 1 to 10), elaborate on the part of the study which they found the most a burden and to elaborate on how they experienced the FMT-procedure.

4. FMT-related AEs in PD patiente after FMT, assessed by the registration of FMT-related AEs.

This will be mainly descriptive, based on the registration of AEs. The nature and number of AEs and the relation with FMT will be described.

Furthermore, blood results of one week and three months post-FMT can be compared to pre-FMT by using linear mixed models for normally distributed numerical variables (converted into logarithmic form in case of a skewed distribution) or by generalized estimating equation (GEE) for numerical variables that are converted into categorical variables. These models take missing values into account, when the data is missing at random. The investigators will attempt to prevent missing values or, if not possible, to minimize the amount of missing values. A two-tailed p<0.05 will be considered statistically significant.

#### 10.3 Other study parameters

As the study group is small and correcting for confounders would decrease the power and as this is a pilot study focussed on safety and feasibility, we will not correct for potential confounders.

#### 10.4 Interim analysis

The DSMB will perform an interim analysis when the 6^th^ patient has received the six weeks follow-up. Further details are described in section 9.5. The nature and number of AEs and SAEs and the relation with FMT will be described by the DSMB. The study will be terminated prematurely when there are definitely FMT-related SAEs in >1 patients at the interim analyses at six weeks post-FMT in the first six patients, or when there is another reason for premature termination of the study according to the DSMB.

### 11. ETHICAL CONSIDERATIONS

#### 11.1 Regulation statement

The study will be conducted according to the principles of the Declaration of Helsinki (amended by 64th WMA General Assembly, Fortaleza, Brazil, published in JAMA November 27, 2013 Volume 310, Number 20) and in accordance with the Medical Research Involving Human Subjects Act (WMO).

#### 11.2 Recruitment and consent

Described in section 8.3

#### 11.3 Objection by minors or incapacitated subjects

Not applicable

#### 11.4 Benefits and risks assessment, group relatedness

Potential benefits:

PD is a progressive disease. No cure or medication that slows down the progression is available. Only PD symptoms can be treated with medication. In an advanced stage of the disease, PD medication may become less effective or motor complications may occur, such as motor fluctuations or dyskinesias, despite using adequate PD medication. For some of these patients deep brain stimulation may help, but many patients still have PD symptoms after the procedure and a large portion of patients is not eligible. Finding a new treatment strategy is crucial for these patients. The gut microbiota is considered to have a role in the pathophysiology of PD and in the metabolization of anti-PD medication. FMT is the most effective gut microbiota intervention and may serve as a new treatment for PD. Several studies suggest that FMT with feces from healthy donors might improve the symptoms of PD, improve the effect of medication such as levodopa and limit their side effects, and/or slow down the disease progression. However, apart from one case report, no evidence is available in humans with PD. This study will provide crucial information about the safety and feasibility of this treatment in patients with PD, which, in the near future, could be further explored in larger trials aiming at determining the efficacy of FMT in PD patients. If FMT appears effective, patients that participate in this study may experience a decrease in PD symptoms and side-effects of PD medication and maybe even a reduced disease progression. Furthermore, they will contribute to an increase in knowledge on the pathophysiology of PD.

Potential risks:

These are described in section 9.2 of this protocol.

Study load:

Prior to FMT a bowel lavage is needed. To this end, the patient is requested to drink 2 liters of macrogol + electrolytes in a relatively short time period. It is usually spread over the day before FMT. Furthermore, the patient is not allowed to eat on the day of FMT prior to FMT, which is a standard regimen before gastroscopy. Patients have to take vancomycin for five days and one pill of domperidon before FMT (and in case of obstipation, bisacodyl for two days ante noctem): this is usually not considered a burden.

The FMT-procedure requires a gastroscopy to inject the fecal suspension directly into the horizontal duodenum or to insert a nasoduodenal tube (130 cm length and 3,3 mm diameter) through the nose with a pediatric gastroscope for later infusion of the fecal suspension, which are both minimally invasive procedures. The patient and the investigator or gastroenterologist can decide together which administration route is preferred (e.g. dependent on the anatomy of the nose or stress of the patient). The nasoduodenal tube will remain in place until approximately 30 minutes after the FMT. The patient can choose to receive mild sedation (midazolam) before or during the gastroscopy. The injection of the fecal suspension through the nasoduodenal tube or gastroscope is usually not considered a burden. On the day of FMT, the patient will be in the hospital for approximately 2-4 hours including an observation period of at least two hours at the day-care department.

The patient has to visit the centre six times in total and will have two telephone appointments. PD patients could be less mobile, which could make a visit to the hospital difficult. Therefore, the number of visits was minimized.

Blood will be drawn three times, which patients might find unpleasant.

The tests in this study, questionnaires, diaries, and the collection of stool samples are usually not considered a burden.

A preliminary version of this study protocol was discussed with two Parkinson patients (patient-investigators), appointed by the Dutch Parkinson patients association (Parkinson vereniging), to review the study load, the safety and the patient-centered value of the study.

#### 11.5 Compensation for injury

The sponsor/investigator has a liability insurance which is in accordance with article 7 of the WMO.

The sponsor (also) has an insurance which is in accordance with the legal requirements in the Netherlands (Article 7 WMO). This insurance provides cover for damage to research subjects through injury or death caused by the study.

The insurance applies to the damage that becomes apparent during the study or within four years after the end of the study.

#### 11.6 Incentives

Not applicable

### 12. ADMINISTRATIVE ASPECTS, MONITORING AND PUBLICATION

#### 12.1 Handling and storage of data and documents

All PD patients will receive a pseudonymized study ID by the investigators when they have signed the informed consent form. This study ID will start with X, and then the year of inclusion and the rest will consist of the number of inclusion. The study ID will not contain any patient identifying or clinical data.

All clinical data, blood and stool samples and fecal suspensions will be stored linked to this pseudonymized study ID. This study ID will also be linked to patient identifying data in a separate document (subject identification code list). The patient identifying data (linked to the study ID) will be stored on another location than the clinical research data (linked to the study ID). Patient identifying data will be stored for safety reasons. The informed consent form will inform the patients on this.

All clinical research data will be stored in a password protected web-based database (Castor) at the LUMC. Questionnaires and diaries will be on paper (because some patients might be of older age and not familiar with computers). This data will also be entered into Castor. The paper questionnaires will be stored in a secured environment at the LUMC, containing only the pseudonymized study ID. The patient identifying data will be password-protected and stored in a datasafe on secured servers of the LUMC. Only the responsible investigators that are involved in this study will have access to the patient identifying information.

If sent by e-mail, patient data will be sent linked to the pseudonymized patient number and via secure routes.

The autologous fecal suspension of the PD patients and quality control stool samples will be stored in freezers of the NDFB in secured rooms, labelled with the study ID and date of donation. The PD patient stool samples (labeled with the study ID and date of donation) for this study will be stored in freezers of the NDFB in secured rooms of the LUMC. Sample collection, processing and storage related data will be stored in a for CMAT specialised metadata standard using SampleNavigator. The raw 16S sequencing data of the stool samples will be stored on LUMC’s high performance computer cluster in a folder with restricted access, and will anonymously be submitted to a public repository European Nucleotide Archive.

The results of the blood analysis will be located in the Electronic Patient File of the LUMC and will be entered into Castor.

The blood and fecal samples that will be provided to the LUMC Biobank Parkinson will be handled confidentially and coded. They will be stored in secured rooms in the LUMC Biobank Parkinson. The regulations of the LUMC Biobank Neurological Diseases will be applicable to the LUMC Biobank Parkinson. In case no permission is provided for storage in the LUMC Biobank Parkinson, the autologous fecal suspension with quality control stool samples and the other stool samples will still be stored in freezers of the NDFB for 20 years for safety reasons (in case of an SAE, these can be tested to find out whether the SAE is related to FMT and the autologous suspension can be administered to the patient). The blood and serum samples will still be stored at the department of Clinical Chemistry and Laboratory Medicine of the LUMC. For the duration of the storage of the blood and serum samples needed for this study, the policy of the clinical chemistry of the LUMC will be followed, which includes 24 hours for EDTA tubes and 6 days for Serum gel tubes.

All data on the subjects and the fecal samples for this study will be destroyed 20 years after end of the study.The coded feces and blood samples in the LUMC Biobank Parkinson will be stored for indefinite duration as these serve for future research purposes.

#### 12.2 Monitoring and Quality Assurance

Monitoring will be executed by internal monitors of the LUMC according to the monitor plan.

#### 12.3 Amendments

Amendments are changes made to the research after a favourable opinion by the accredited METC has been given. All amendments will be notified to the METC that gave a favourable opinion.

All substantial amendments will be notified to the METC and to the competent authority.

Non-substantial amendments will not be notified to the accredited METC and the competent authority, but will be recorded and filed by the sponsor.

#### 12.4 Annual progress report

The sponsor/investigator will submit a summary of the progress of the trial to the accredited METC once a year. Information will be provided on the date of inclusion of the first subject, numbers of subjects included and numbers of subjects that have completed the trial, SAEs/ serious adverse reactions, other problems, and amendments.

#### 12.5 Temporary halt and (prematurely) end of study report

The investigator/sponsor will notify the accredited METC of the end of the study within a period of eight weeks. The end of the study is defined as the last patient’s last visit.

The sponsor will notify the METC immediately of a temporary halt of the study, including the reason of such an action.

In case the study is ended prematurely, the sponsor will notify the accredited METC within 15 days, including the reasons for the premature termination.
Within one year after the end of the study, the investigator/sponsor will submit a final study report with the results of the study, including any publications/abstracts of the study, to the accredited METC.

#### 12.6 Public disclosure and publication policy

The results of this investigator-initiated study will be sent in for publication to peer-reviewed journals, despite of the results. Furthermore, this clinical trial will be registered in a public trial registry before the first patient is recruited.

### 13. STRUCTURED RISK ANALYSIS

#### 13.1 Potential issues of concern

a. Level of knowledge about mechanism of action

Available literature on the possible role of the gut microbiota in PD and evidence on the efficacy of FMT in PD is described in section 1, 6.2 and 6.3 of this protocol.

These studies suggest that changing the gut microbiota by means of an FMT could act on the pathophysiology of the disease and/or development of levodopa-mediated motor complications. Symptoms might decrease due to a direct effect of the changed gut microbiota on the gut-brain axis. They might be attenuated due to less production of pro-inflammatory cytokines with less intestinal inflammation and oxidative stress and subsequently less aggregation of αSyn in the ENS and CNS. Another important possibility is that FMT could lead to an increased absorption or less inhibition of PD medication in the gut due to the changed gut microbiota, resulting in an improved efficacy of the medication and less motor complications.

b. Previous exposure of human beings with the test product(s) and/or products with a similar biological mechanism

In the last two decades, an increasing amount of FMT-studies have been published. FMT has been studied in patients with recurrent and severe CDI ^42-45^, neurological disorders^68,69,84-89^, inflammatory bowel disease^58^, irritable bowel syndrome^90,91^, pouchitis^92^, metabolic syndrome^93^, hepatic encephalopathy^54,67^, chronic hepatitis B infection^94^, graft versus host disease^95^, chronic intestinal pseudo-obstruction^96^, small-intestinal bowel overgrowth^96,97^, microscopic colitis^97,98^, multi-drug resistant organisms^99,100^, and sepsis^101^. Overall these studies show a beneficial effect of FMT on the disease with a good safety-profile. Some studies already observed an effect after a few days. There were a few studies that observed only a transient effect or effects were clearly better after repeated FMTs.

These studies included several diseases with colitis, such as rCDI and ulcerative colitis. FMT is a very effective treatment for rCDI^42-44^ and severe CDI^45^, with cure rates of 80-95% for rCDI ^42-44^. For ulcerative colitis, a decreased or absent intestinal inflammation was observed after FMT in several patients^102^. Studies in rCDI and IBD suggest that an FMT could lead to a decrease of intestinal inflammation, which may also be the case in PD, with potentially less aggregation of αSyn in the ENS and CNS as a result. However, one study showed an increased systemic inflammation in three healthy subjects that received capsules with feces from healthy donors, but these effects were mostly transient^80^. One patient developed SIRS.

For PD patients, there is only one case report (126) and one communication in a divulgative magazine that described the effect of FMT (described in more detail in section 6.3).

c. Can the primary or secondary mechanism be induced in animals and/or in *ex-vivo* human cell material?

Mouse studies with animal models for colitis have shown that FMT may reduce intestinal inflammation ^103,104^. Furthermore, the potential beneficial effect of FMT in PD is shown in several animal studies with PD animal models, which are described in section 6.2.

d. Selectivity of the mechanism to target tissue in animals and/or human beings

FMT in patients with rCDI and a reduced gut microbiota diversity leads to improvement of gut microbiota diversity after FMT. The gut microbiota of patients alters towards the gut microbiota composition and alpha-diversity of the gut microbiota of the donor after infusion of the feces of the donor^42^. In PD, alpha-diversity (within-subject diversity) appears similar to that of controls^23,26-28^, but the gut microbiota composition differs^18,21-23^. PD patients have more pro-inflammatory gut bacteria, compared to healthy controls. After receiving feces from a healthy donor, we expect the gut microbiota composition to change towards that of the donor and we expect to observe a decrease in pro-inflammatory gut bacteria. We hypothesize that the concomitant decrease in intestinal inflammation may decrease αSyn pathology in the ENS and CNS or the altered gut microbiota may change the availability and/or pharmacokinetics of PD medication. The selected donors for this study will be rationally selected based on available literature.

The only case report of PD showed an increase in alpha-diversity seven days post-FMT and a decrease after 90 days. The gut microbiota composition changed towards that of the donor. However, this is n=1.

By altering the gut microbiota composition several pathways may be altered, including immunological, endocrine, metabolic and/or neural pathways. Therefore, FMT is likely not selective for the target tissue. However, previous studies in rCDI have shown that it is a safe treatment ^48,70-72,79^ and animal studies with PD mouse models have shown a beneficial effect of healthy donor FMT^38,39,65^.

e. Analysis of potential effect

As mentioned in section 6.2, several animal studies have suggested a beneficial effect of healthy donor FMT in PD. However, no studies have been performed with FMT in human PD patients (except for one case report).

FMT appears a safe treatment for patients with rCDI (more details in section 9.2).

Since there are no treatments available that cure PD or slow down the progression and most PD patients with advanced disease experience less effectivity and/or adverse effects of PD medication, the development of a new treatment strategy is crucial. As animal studies have already shown a beneficial effect, a pilot study with a low sample size that assesses the safety and feasibility of FMT in PD patients appears a logical next step.

f. Pharmacokinetic considerations

Not applicable as FMT is no medication.

g. Study population

In- and exclusion criteria are described in section 4.

By excluding PD patients with Hoehn and Yahr stage 5, PD patients that have severe swallowing problems and PD patients who are not capable of understanding and complying with the study requirements, subjects with the most severe stage of PD are filtered out. By excluding patients with a change in type or dose of PD medication in the previous three months, we aim to include patients who have a relatively stable disease. By excluding patients with a disease duration of less than five years, patients with atypical parkinsonisms will be most likely excluded. Pregnant or lactating women or women with a pregnancy wish will be excluded and women with child bearing potential will be asked to use efficient contraception.

h. Interaction with other products

The use of antibiotics for (systemic) infections during or after FMT could affect the microbiota and thus diminish the effect of FMT. Furthermore, FMT may alter availability and pharmacokinetics of anti-PD (or other) medication, e.g. by decreasing the bacterial tyrosine decarboxylase load in the gut of patients. Then, Levodopa may be less frequently converted to dopamine outside of the brain. Dopamine cannot pass the blood-brain-barrier and, therefore, more Levodopa will be available for the brain. Several other mechanisms for altering availability and pharmacokinetics of medication are theoretically possible.

i. Predictability of effect

The alteration of the gut microbiota of the PD patients could be considered a biomarker for the engraftment of the donor microbiota. Furthermore, blood analysis components could be a biomarker for some (S)AEs.

We do not use biomarkers for measuring the effect of FMT on PD symptoms since this is a pilot study which primarily focusses on safety and feasibility. Effects of FMT will be assessed by using MDS-UPDRS, SENS-PD, MOCA and Hoehn and Yahr scores, a diary and questionnaires. Safety will be assessed by registration of FMT-related SAEs. Feasibility of FMT will be assessed by the registration of the number of included patients that cannot undergo FMT due to a patient- or procedure-related reason.

Several mouse studies and one case report show a positive effect of healthy donor FMT. It is important to note that publication bias may contribute to the lack of reported negative studies.

We expect the symptoms of the PD patients to decrease after FMT due to a decrease of intestinal inflammation with a subsequent decrease in αSyn pathology in gut and brain or due to increased availability and altered pharmacokinetics of PD medication.

j. Can effects be managed?

Patients will be monitored closely. The patient will be instructed to always contact the investigators in case of any (S)AE and (S)AEs will be assessed during follow-up visits/telephone appointments. In case of doubt on the condition of the patient, a physician will see the patient as soon as possible. Physicians will be aware of the participation in the study, as this is stated in the electronic patient file and on a card which participants will carry with them and which includes information on the study and contact details to be used in case of emergency. A DSMB will support the study by monitoring the safety of the participants and by performing an interim analysis.

From every patient that receives a donor FMT, a ready-to-use autologous rescue fecal suspension will be prepared and stored prior to FMT. In case of FMT-related SAEs, the Parkinson working group will decide whether it may be useful to perform an autologous rescue FMT and/or provide antibiotics, as this may potentially reverse the donor FMT effect.

In previous studies, patients who received FMT via lower GI routes were more likely to develop SAEs than those who received FMT by upper GI routes^48,70-72,79^. In this pilot study, the upper GI route will be used. Gastroscopy and placement of a nasoduodenal tube are very common interventions in hospitals. The experts on the endoscopy department of the LUMC are experienced with performing these procedures. In case of doubt, the position of a nasoduodenal tube will be checked by X-ray. To prevent aspiration, the fecal suspension will be infused slowly and the patients will be kept in an upright position during and after infusion. After FMT, the patients will be monitored for at least two hours before being discharged. When this is not contraindicated, one pill of domperidone 10 mg will be self-administered orally on the day of FMT prior to FMT, to prevent nausea and to improve gastric motility. In case of nausea after FMT, domperidone could also be used.

#### 13.2 Synthesis

Since there are no treatments available that cure PD or slow down the progression and most PD patients with advanced disease experience less effectivity and/or adverse effects of PD medication, the development of a new treatment strategy is crucial. Animal studies suggest a potential role of the gut microbiota in disease pathophysiology and a potential beneficial effect of a healthy donor FMT in mouse models of PD. However, no studies have been performed with FMT in human PD patients (except for one case report that shows a beneficial effect) and a pilot study with a low sample size that assesses the safety and feasibility of FMT in PD patients appears a logical next step.

In previous studies with FMT for rCDI, the percentage of patients experiencing FMT-attributable AEs was approximately 20-45%. However, these are mostly mild and self-limiting. Furthermore, only 0-5% of patients developed SAEs which were probably or definitely related to the FMT or to the procedure^70-72^. SAEs that are possibly attributable to FMT or to the procedure via upper GI route include aspiration pneumonia, septicemia or other infections, fever, peritonitis, upper GI hemorrhage or death^48,70-72,79^. More details are described in section 9.2. FMT appears a safe treatment for patients with rCDI. However, the type and probability of specific procedure-related problems and (S)AEs in PD patients is unknown, and will be the main objective of this pilot study.

Risks are minimized by several measures. By excluding PD patients with Hoehn and Yahr stage 5, PD patients that have severe swallowing problems and PD patients who are not capable of understanding and complying with the study requirements, subjects with the most severe stage of PD are filtered out. By excluding patients with a change in type or dose of PD medication in the previous three months, we aim to include patients who have a relatively stable disease. By excluding patients with a disease duration of less than five years, patients with atypical parkinsonisms will be most likely excluded. Pregnant or lactating women or women with a pregnancy wish will be excluded and women with child bearing potential will be asked to use efficient contraception. Furthermore, patients will be monitored closely. The patient will be instructed to always contact the investigators in case of any (S)AE and (S)AEs will be assessed during follow-up visits/telephone appointments. All (S)AEs will be followed until they have abated, or until a stable situation has been reached. In case of doubt on the condition of the patient, a physician will see the patient as soon as possible. Physicians will be aware of the participation in the study, as this is stated in the electronic patient file and on a card which participants will carry with them and which includes information on the study and contact details to be used in case of emergency. A DSMB will support the study by monitoring the safety of the participants and by performing an interim analysis. We cannot exclude that PD symptoms and/or disease progression might increase after FMT, although this phenomenon has not been observed in the FMT-studies with mouse models of PD and the case report on FMT in a PD patient. From every patient that receives a donor FMT, a ready-to-use autologous rescue fecal suspension will be prepared and stored prior to FMT. In case of FMT-related SAEs, the Parkinson working group will decide whether it may be useful to perform an autologous rescue FMT and/or provide antibiotics, as this may potentially reverse the donor FMT effect.

In previous studies, patients who received FMT via lower GI routes were more likely to develop SAEs than those who received FMT by upper GI routes^48,70-72,79^. In this pilot study, the upper GI route will be used. Gastroscopy and placement of a nasoduodenal tube are very common interventions in hospitals. The experts on the endoscopy department of the LUMC are experienced with performing these procedures. In case of doubt, the position of a nasoduodenal tube will be checked by X-ray. To prevent aspiration, the fecal suspension will be infused slowly and the patients will be kept in an upright position during and after infusion. After FMT, the patients will be monitored for at least two hours before being discharged. When this is not contraindicated, one pill of domperidone 10 mg will be self-administered orally on the day of FMT prior to FMT, to prevent nausea and to improve gastric motility. In case of nausea after FMT, domperidone could also be used. Vancomycin, Kleanprep, domperidone, bisacodyl and midazolam are not mentioned in section 13.1, as these are used within its indication.

We think the risks on worsening of PD symptoms, increased disease progression or other SAEs are low since:

- A beneficial effect of healthy donor FMT in PD is suggested in animal studies.

- FMT is considered a safe treatment for other indications.

- Fecal material from healthy donors is used that have been rationally selected according to stringent safety criteria of the NDFB and based on available literature.

In conclusion, we believe that the potential scientific benefit will outweigh the risks of FMT treatment.

### APPENDICES

#### Appendix A: Application form

[Available upon request: please contact the corresponding author to request access to this information]

#### Appendix B: FMT protocol of the NDFB

[Available upon request: please contact the corresponding author to request access to this information]

#### Appendix C: Product information of the fecal suspension for Fecal Microbiota Transplantation

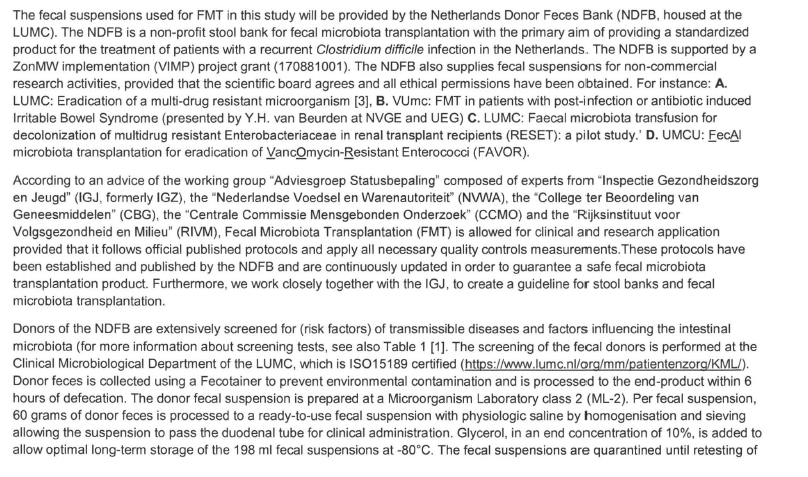

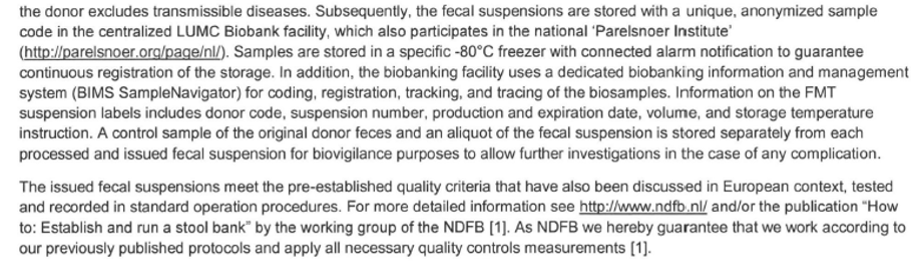

**Table 1:**

| **Exclusion criteria by first screening questionnaire***  Age <18 or ≥ 55^a^, BMI <18.5 or > 25, high risk faecal- and or blood transmittable diseases, recent antibiotic use (<6 months), gastrointestinal complaints (for example diarrhoea, obstipation or irritable bowel like symptoms), recent travel to endemic areas of gastrointestinal pathogens, (first degree relative with) inflammatory bowel disease, known systemic infection, liver diseases like hepatic encephalopathy or Non Alcoholic Fatty Liver Diseas, History of cancer, including GI malignancy or polyposis, first degree relative with a GI malignancy < 60 years or family history of genetically-driven cancer, metabolic syndrome, substantial comorbidity, chronic medication use, autism, auto-immune disorders, neurological/neurodegenerative disease, atopic diseases, frequent healthcare contacts^b^. | |
| --- | --- |
| **Laboratory screening serum****   - Hepatitis A (IgM + IgG)^c^ - Hepatitis B (HBsAg + anti-Hbcore) - Hepatitis C (anti-HCV) - Hepatitis E (IgM + IgG)^c^ - HIV (anti-HIV, type 1 and 2) - Lues; *Treponema pallidum* (Ig) - Cytomegalovirus (IgM + IgG)^c^ - Epstein Barr Virus (IgM + IgG)^c^ - HTLV^d^ - *Strongyloïdes* (IgG1/IgG4)^e^ - *Coronavirus (IgM + IgG)* | **Laboratory screening faeces****   - *Clostridioides diffic*ile (PCR) - *Helicobacter pylor*i (antigen test) - Bacterial gastro-enteritis: (PCR): *Salmonella* spp. *Campylobacter* spp., *Campylobacter jejuni, C. coli*, *Shigella* spp., *Yersinia enterocolitica* and *Y. pseudotuberculosis*, *Aeromonas* spp., *Plesiomonas shigelloides,* and Shiga Toxin producing *E.coli* - Antibiotic resistant bacteria (culture); ESBL and/or carbapenemase producing bacteria, Aminoglycoside AND quinolone resistant Enterobacteriacese, vancomycin resistant enterococci and methicillin resistant *Staphylococcus aureus* - Viral pathogens (PCR): Norovirus serotype I+II, Astrovirus, Sapovirus, Rotavirus, Adenovirus 40/41, Adenovirus non-40/41, Enterovirus, Parechovirus, Coronavirus - Parasites (PCR): *Giardia lamblia*, *Entamoeba histolytica, Cryptosporidium parvum and C. hominis, Microsporidium spp*, Cystoisospora belli, Cyclospora cayetanensis. *Strongyloïdes* ^e^ - Microscopy for ova, cysts and larvae: for example: *Blastocystis* sp. |
| **Questionnaire recent health status: During donation of faeces*****  Stool frequency/pattern, general health, use of antibiotics, travel history, sexual behaviour | |

Donor screening by questionnaire, when donors pass the questionnaire, laboratory screening of faeces will follow. Faeces is first screened for the presence of *Dientamoeba fragilis* and *Blastocystis* sp*.* When negative, other pathogens are investigated, after which screening of serum is performed. If a donor is suitable for donation, before every donation a questionnaire about the recent health status should be filled in. ^a^ Or 60 years when no colon cancer is detected during the national colon cancer screening programme. ^b^ Donors are not allowed to have frequent healthcare contacts (working in direct patient care or laboratory handling of infectious agents). ^c^ In case of rescreening, only repeat when prior sero-negative, to detect seroconversion and subsequent potential transmission via faeces. ^d^ In case of rescreening only when travelled outside Europe, ^e^ In case of rescreening only when travelled to Middle and South America, Africa or Asia. *: In case of abnormalities in the interview or questionnaire, individuals are usually definitely rejected as a donor. **: When donors pass the questionnaire and interview, but a pathogen is detected by faeces examination or serological screening, the decision whether to definitely of temporarily exclude a donor depends on the detected pathogen. ***: When a deviation is found in the questionnaire on recent health status during donation of faeces, the donor is usually temporarily excluded. In case donors experience a transient mild illness, such as a common cold or diarrhea, they are mostly temporarily excluded from donation, but faecal suspensions of the period within the three-month interval can still be used after a negative rescreening.

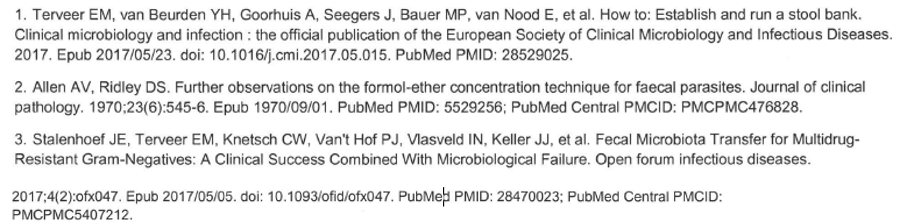

#### Appendix D: Safe application of Faecal Microbiota Transplantation in the Netherlands

Prof. dr. Ed. J. Kuijper, drs. E.M Terveer, prof.dr. H. Verspaget and dr. Josbert Keller

**On behalf of the Nederlandse Donor Feces Bank (NDFB) Working Group:**

Dr. M. P. Bauer, infectious diseases specialist, LUMC

Prof. M.A. Benninga (MB), pediatrician and gastroenterologist, Department of Pediatric Gastroenterology and Nutrition, Amsterdam UMC, location AMC

Drs. E. Boeije-Koppenol, psychologist and coordinator NDFB, LUMC

E.K.L. Berssenbrugge, technician, NDFB, LUMC

Dr. Y.H. van Beurden, gastroenterologist, Amsterdam UMC, location VUmc

Dr. A. Goorhuis (AG), infectious diseases speclalist, Amsterdam UMC, location AMC

Dr. J.J. Keller (JK), gastroenterologist, Haaglanden Medical Centre, The Haque

Prof. dr. E.J Kuijper (EK), medical microbiologist, Dep. Med. Microbiol, LUMC

Dr. P.P. H le Brun (PB), pharmacist, Department of Clinical Pharmacy and Toxicology, LUMC

drs. E. van Lingen, PhD student gastroenterology and coordinator NDFB, LUMC

LUMC

Prof. CJ. Mulder, Gastroenterology, Amsterdam UMC, location VUmc

Dr. E. van Nood (EvN), infectious diseases specialist, Erasmus Medical Centre

Drs. R.E Ooijevaar, (RO), PhD student Gastroenterology, Amsterdam UMC, location VUmc

Dr. J. van Prehn, medical microbiologist, LUMC

Prof. dr. C.I.J Ponsioen (CP), gastroenterologist, Department of Gastroenterology and Hepatology, Amsterdam UMC, location AMC

Drs. B. Rethans (BR), PhD student Gastroenterology, Amsterdam UMC, location AMC

Drs. E.M Terveer (ET), medical microbiologist and coordinator NDFB, Dept. Med Microbiol,

Prof.dr.ir. H.W. Verspaget (HV), Department of Biobanking, LUMC

Prof. dr. C.M.J.E. Vandenbroucke-Grauls, Medical Microbiologist, Amsterdam UMC, location VUmc

Drs. K.E.W.Vendrik, PhD student Medical Microbiology and coordinator NDFB, LUMC

**Approved version by the Netherlands Donor Feces Bank (NDFB), Leiden; 10 October, 2019.**

Content

#### 14.1 Introduction

Prof.dr. Ed J. Kuijper and dr. Josbert Keller

The “Adviesgroep Statusbepaling” with experts from the “Inspectie Gezondheidszorg en Jeugd (IGJ)”, “Nederlandse Voedsel en Warenautoriteit (NVWA)”, “College ter Beoordeling van Geneesmiddelen (CBG)”, “Centrale Commissie Mensgebonden Onderzoek (CCMO)” and the National Institute for Public Health and the Environment (“RIVM”) assesses the legal status of medical products whose status is not clear. They have recently discussed if donor faecal microbiota transplantation (FMT) used to treat multiple recurrent *Clostridiodes difficile* infections (rCDI) meets the definition of a drug as expressed in Article 1 of the Medicines Act and should therefore be considered a medicine. **The Advisory Group has concluded that the product cannot be classified as a medicine**, because the precise mechanism of action is not known. Based on the different effects currently attributed to FMT, it cannot be classified under the legal definition of a drug. However, the IGJ considers **careful and safe application of FMT** essential. In addition to the efficacy and safety of FMT in the treatment of rCDI, the product must also meet appropriate quality and safety requirements for possible application in other diseases. Therefore, the IGJ has asked the field – i.e. those who apply treatment with donor faeces - to establish a framework of standards, in order to guarantee safe application of FMT in the Netherlands.

In compliance with this request, the Nederlandse Donor Feces Bank (NDFB) composed a **national multidisciplinary committee** to develop a guidance document and contacted two European Societies (“European Society for Clinical Microbiology and Infectious Diseases” and the “United European Gastroenterology”) to harmonize the activities with other donor feces banks in Europe. This resulted in the development of this guidance document (not a guideline), and in two European guidance documents that will be completed by the end 2019/early 2020.

The NDFB working group formulated well-built questions in accordance with the PICO processs and summarized the literature according to

<https://acpjc.acponline.org/Content/123/3/issue/ACPJC-1995-123-3-A12.htm>.

The following topics were discussed;

| **TOPIC** | **Lead** | **Contributors** |
| --- | --- | --- |
| Actual and theoretical risks of FMT | AG | RO, EvN, BR, EK |
| Q1: What donor factors influence the outcome of faecal microbiota transplant? | RO | CP, MB, BR, JK, ET |
| Q2: What recipient factors influence the outcome of faecal microbiota transplant? | EvN | CP, MB, EK, JK |
| Q3: Where and under which conditions should donor stool samples be processed and stored? | HV | PB, BR (CP), ET, EK, JK |
| Q4: What factors related to the preparation of the transplant influence the outcome of faecal microbiota transplant? | ET | CP, MB, EK, JK |
| Q5: How should FMT be administered to patients? | JK | CP, MB, EvN, AG |
| Q6: What is the general approach to follow-up post-FMT? | EK | HV,CP, MB, JK |

Four documents were used as basis:

Cammarota G, Ianiro G, Tilg H, Rajilic-Stojanovic M, Kump P, Satokari R, et al. European consensus conference on faecal microbiota transplantation in clinical practice. Gut. 2017. Epub 2017/01/15. doi: 10.1136/gu

Terveer EM, van Beurden YH, Goorhuis A, Seegers JFML, Bauer MP, van Nood E, Dijkgraaf MGW, Mulder CJJ, Vandenbroucke-Grauls CMJE, Verspaget HW, Keller JJ, Kuijper EJ.. How to: Establish and run a stool bank. Clin Microbiol Infect. 2017;23:924-930.

Mullish BH, Quraishi MN, Segal JP, McCune VL, Baxter M, Marsden GL, Moore DJ, Colville A, Bhala N, Iqbal TH, Settle C, Kontkowski G, Hart AL, Hawkey PM, Goldenberg SD, Williams HRT. The use of faecal microbiota transplant as treatment for recurrent or refractory *Clostridium difficile* infection and other potential indications: joint British Society of Gastroenterology (BSG) and Healthcare Infection Society (HIS) guidelines. Gut. 2018;67:1920-1941

Cammarota G, Ianiro G, Kelly CR, Mullish BH, Allegretti JR, Kassam Z, Putignani L, Fischer M, Keller JJ, Costello SP,Sokol H, Kump P, Satokari R, Kahn SA,Kao D,Arkkila P,Kuijper E, Vehreschild MJGT, Pintus C, Lopetuso LR, Masucci L, Scaldaferri F, Terveer EM, Nieuwdorp M, Lopez Sanroman A, Kupcinskas J, Hart A, Tilg H, Gasbarrini A. International Consensus Conference on stool banking for faecal microbiota transplantation in clinical practice. Revised version submitted and accepted for publication in Gut, September 2019

#### 14.2 Actual and theoretical risks of FMT

Dr. A. Goorhuis and dr. Y van Beurden

FMT is a powerful treatment option against rCDI. After publication of the first randomized trial showing the efficacy of FMT in patients with rCDI,^1^ it has been implemented as a standard therapy for this condition, when antibiotic therapy alone has failed to prevent rCDI. Both European^2^ and American^3^ guidelines include FMT for the management of rCDI. Less evidence exists for the use of FMT as direct therapy for severe CDI, refractory to antibiotic treatment, though several studies have reported positive results.^4,5,6,7^ In this case, FMT is not applied to prevent recurrent disease, but in the management of severe disease refractory to antibiotic therapy, to combat toxin-producing *C. difficile* directly*.* Although the use of FMT for treatment of rCDI has gained consensus worldwide, for FMT to be used safely, several issues should be addressed, such as the route of FMT delivery and the indication as direct treatment of severe CDI. The route of FMT delivery can either be proximal, i.e. FMT administration per nasoduodenal tube or capsules (the latter are not available in the Netherlands), or distal (i.e. FMT per colonoscopy or enema). To date, there is no consensus which of the two routes is generally preferable, as both are safe and the success rates of FMT via both routes seem comparable, although the long-term effects are unknown. However, specific safety concerns apply to each of the two routes, these will be discussed below. The second issue, also discussed below, pertains to the efficiency and safety of FMT as direct treatment of severe CDI. One randomized controlled trial has been performed comparing single versus multiple FMT infusions by colonoscopy in 56 patients with refractory severe CDI. Administration of multiple FMTs had a high success rate, but the study was not blinded and not designed to assess the efficacy of FMTs in treating severe CDI.^4^ A few case series also indicate that FMT can be life-saving in the clinical setting of therapy-refractory severe CDI, and avoids the need for surgical intervention.^5,6,7^

Commonly, post-FMT adverse events in patients with rCDI are mild and transient, such as diarrhoea, cramping, flatulence and belching, constipation; however, rare serious adverse events, including fever, bacteraemia, intestinal perforation, aspiration pneumonia, and death, have been described.^8,9,10^

**Route of FMT administration**

FMT administration per nasoduodenal tube (proximal route)

The proximal route of FMT administration is currently the standard route of administration in the Netherlands, whereas the distal route is more often applied in the USA and across southern Europe. Therefore, the majority of clinicians in the Netherlands have gained experience with this mode of delivery of donor stools.

In a recent study, complications and safety of FMT per nasoduodenal tube were assessed in 39 patiens.^8^ No long term side-effects were observed during a 6-month follow-up period. Serious adverse events (SAEs) were observed in nine patients within 12 weeks after FMT. In total, SAEs occurred in 9 (23%) patients, of which 4 (10%) were deemed procedure-related, and 4 (10%) were non-procedure-related. One patient (3%) died 15 days after FMT due to pneumonia. A causal relation with FMT could not be excluded. This patient had a swallowing disorder and was fed through a PEG-tube. The FMT had been administered through a nasoduodenal tube, which was placed adjacent to the PEG-tube. In the three-hour observation period after the procedure, the patient experienced mild and transient regurgitation complaints, but no signs of aspiration. One week after FMT, she developed pneumonia and died, despite antibiotic treatment. Although no causative organism was identified, aspiration of donor feces could have been the cause of this pneumonia. This case has led to an amendment in the national FMT-protocol, which now designates swallowing disorders as contra-indication for proximal FMT, because of an increased aspiration risk. The other 4 procedure-related events comprised regurgitation and/or vomiting of donor faeces. In retrospect, these events were partly preventable. The first patient had a pre-existing bowel condition that compromised the speed of bowel passage, the second patient had consumed a considerable amount of food within one hour after FMT, the third patient developed abdominal cramps during FMT, but the procedure was not terminated, and the fourth patient had a history of a congenital syndrome and mental retardation, including a swallowing disorder, for which she was tube-fed. None of these patients developed further complications. The 4 other SAEs that were not attributable to FMT comprised hospital admissions within 12 weeks after FMT, for reasons unrelated to FMT.

Based on our experience to date, we have reduced the total amount of donor feces suspension to be administered by the proximal route from 500 ml to 200 ml.

Recommendations to avoid regurgitation or vomiting and subsequent aspiration are:

- Reduce stress/anxiety
- Do not increase the speed with which FMT is infused, and pause or stop the infusion if necessary
- Avoid food or fluid ingestion shortly (<1 hour) after FMT
- Proximal FMT is contra-indicated in patients with pre-existing abdominal conditions that compromise bowel passage
- Take specific measures for patients who are fed through a PEG tube, such as consultation of a gastroenterologist, who can pass a jejunal extension through a PEG tube
- Assess aspiration risk in each patient; if risk is increased, this is a contra-indication for proximal FMT; consider administration of FMT via colonoscopy
- A swallowing disorder is a contra-indication for proximal FMT
- During FMT, monitor continuously for symptoms of abdominal discomfort or nausea, and discontinu the procedure when symptoms develop
- Administer metoclopramide if nausea develops
- Keep the patient under hospital observation for at least three hours after Post FMT

FMT administration per colonoscopy (distal route)

The distal route of FMT is used more frequently in southern Europe and the USA. In the Netherlands, this route is usually only used when contra-indications exist for the proximal route. The main reason is that the distal route is more invasive and colonoscopic delivery requires specific expertise of a gastroenterologist. Lower routes of delivery include both enemas and colonoscopy. FMT administered with enemas is less effective than colonoscopic FMT and should therefore be reserved as last resort option, especially since FMT per colonoscopy has also been proven safe and effective. The advantage of distal FMT over proximal FMT is the opportunity to directly inspect the intestinal mucosa, which offers the opportunity to assess the presence of pseudomembranes and to grade the severity of disease. Distal FMT can also be of pivotal importance in cases when the CDI diagnosis is uncertain. Disadvantages of distal FMT are that the procedure is more invasive, and that it requires the specialist care of a gastroenterologist, which is not necessary for proximal FMT. Furthermore, distal FMT carries the risk of incremental damage to an already diseased colonic wall, with an increased risk of bowel perforation, especially in patients with severe colitis. However, in experienced hands, colonoscopy is generally safe in these patients.

**Recommendation to administer FMT: The proximal route of FMT administration is currently the standard route that is used in the Netherlands, but the distal route can be used in patients with a contra-indication for FMT per proximal route.**

**FMT as treatment of severe CDI**

Several recent studies and experience of experts in the field indicate that FMT can be life-saving in the clinical setting of refractory severe FMT, avoiding surgical intervention.

Subjects with severe CDI are particularly frail, not only because of colitis, but also because they usually suffer from multiple comorbidities, and are critically ill. Importantly, the clinical condition of these patients can deteriorate rapidly. There is no universal consensus regarding the exact timing of FMT in patients with severe CDI, but it should be considered when the infection is refractory to antibiotic therapy. Indeed, the establishment of a FMT program was shown to reduce the rates of surgical procedures for severe CDI.^11^ Moreover, FMT appeared to decrease mortality in patients with severe CDI refractory to antibiotic therapy,^4,5,6,7^ and could be considered as a therapeutic option for this condition. In the absence of clear guidelines regarding the role of FMT in the treatment of severe refractory CDI, the risk of performing FMT in these patients should be weighed individually, by a multidisciplinary team of experts, preferably consisting of an infectious diseases specialist, a clinical microbiologist, a gastroenterologist and a surgeon.

**Recommendation: In patients with severe CDI when the infection is refractory to antibiotic therapy, FMT should be considered by a multidisciplinary team of experts, preferably consisting of an infectious diseases specialist, a clinical microbiologist, a gastroenterologist and, if required, a surgeon.**

#### 14.3 Q1a. What donor factors should be considered before approval as a stool donor?

Drs. R.E Ooijevaar and drs .E. M. Terveer

*General safety remarks*

Transplantation of fecal microbiota from one individual into a diseased individual poses the theoretical risk of transmission of pathogens and/or the transfer of a perturbed microbiota, leaving the recipient susceptible to several disorders. An extensive screening of potential fecal microbiota donors should be performed. We propose a 3-step screening method of all potential new donors. Following initial approval of a new donor, rescreening should also be regularly performed before releasing donor material for patient care.

*Initial screening algorithm of new donors for transmissible pathogens*

The first step in screening of a potential donor consists of an individual interview combined with a questionnaire to assess the risk of the presence of transmissible pathogens based upon behavior, medical and travel history, and current medication.^105^ The second step is screening of a fecal sample for the presence of possible transmissible pathogens. To reduce screening costs we recommend to first screen for pathogens most prevalent according to the local epidemiology. In the Netherlands, we therefore propose to first examine the fecal sample for *Blastocystis hominis* and *Dientamoeba fragilis*, although no consensus exists among stool banks whether *Blastocystis hominis* carriage should lead to exclusion of donors.^105-107^ Subsequently we propose testing the fecal sample for several other potentially transmissible pathogens (Table 2). General blood testing by complete blood cell count with differential analysis,creatinine and aminotransferases (ALT) can also be considered.

*Screening of active donors*

Prior to every donation a short questionnaire should be filled out by the donor to assess the recent health status. All donations should be quarantined until rescreening has been performed (window of detection phase). Alternatively, a different approach can be applied when fresh stool samples are used. Upon approval through rescreening the donor material can be released for patient care or study purposes. We propose a timely rescreening of each active donor within 1 to 6 months, depending on the number of donations or risk of transmissible diseases (foreign travel, number of recent sexual contacts). For studies with fresh donor stool samples, a quarantine period is not feasible and regular screening with an appropriate risk factor analysis will be sufficient.

**Recommendation: Extensive screening by questionnaire and a personal interview concerning risk factors for transmissible diseases should be mandatory for every new potential donor. A short questionnaire about the recent health status should be completed for each separate donation by active donors.**

**Recommendation: Rescreening should be performed within 1 to 3 months on frozen or fresh donor material, before releasing donor material for patient care or study purposes.**

*Disorders associated with dysbiosis of the gut microbiota*

Dysbiosis of the commensal gut microbiota has been described and linked to several disorders other than recurrent *Clostridioides difficile* infection (r)(CDI) (Table 3).^108-112^ The transfer of disorder-associated microbiota might leave recipients susceptible to development of the respective disorder. It is not always understood if dysbiosis is the driving step in pathophysiology or is caused by a disorder. However due to safety precautions potential donors with (a high risk of) one of these disorders should be excluded from the donor program. The list of disorders associated with dysbiosis should regularly be updated according to the latest literature to uphold the highest safety standards. A list of currently known disorders associated with dysbiosis of the gut microbiota is shown in table 3.

Long-term safety data is still lacking so no firm conclusions on screening protocols can be drawn. New insights might provide changes to the protocols used for screening in the future.

**Recommendation: Donors with (or at high risk of developing) a disorder associated with dysbiosis of the gut microbiota should be excluded from the donor program. The list of disorders should be regularly be updated.**

**Recommendation: The screenings protocol should be adapted immediately upon new insights.**

*Age and body mass index*

Currently there is no consensus on age restrictions for potential donors. Throughout several studies donors between 16 and 60 years old have been used. The gut microbiota decreases in stability and diversity in the elderly of 60 years and older.^113,114^ Furthermore the increased odds for undetected comorbidity such as colon cancer should lead to the exclusion of donors over 60 years of age. To further lower theoretical risks of transmissible disorders, the age limit could be set to 55-60 years (expert opinion).

One case study reports weight gain in a recipient following FMT from an overweight donor.^115^ An association of FMT with weight gain and an increased Body Mass Index (BMI) has not been described in literature since. However, a recent large retrospective cohort study found that a single FMT did not cause weight gain in the recipients.^116^ In concordance with most stool banks, donors who fall outside of the normal BMI range of 18 -25 should be excluded from the donor program because of the high risk of a disturbed microbiota, until further prospective studies confirm otherwise.^105-107,117^

**Recommendation: Potential stool donors should be between >= 16 and <= 60 years of age. Expert opinion: not above age 60.**

**Recommendation: Potential stool donors should have a BMI between 18 and 25.**

*Related and unrelated donors*

The rise of centralized stool banks has made FMT treatment with unrelated donors more readily available.^105-107^ Studies that used related donor for FMT treatment did not show lower efficacy in curing rCDI, but nowadays unrelated donors are usually preferred because of their independence from recipients. Related donors might underreport their own risk factors and risk behavior. Moreover, unrelated donors allow for more rapid transplantation when needed, because of their pre-screening.

**Recommendation: Both related and unrelated stool donors should be considered acceptable. When possible, FMT is best sourced from a centralized stool bank, from a healthy unrelated donor.**

#### 14.4 Q1b. What donor factors influence the efficacy of FMT for treating (r)CDI ?

Drs. R.E Ooijevaar and drs .E. M. Terveer

*Antibiotics and other medication*

The human microbiota is mainly formed by environmental factors, and medication seems to play a major role.^118,119^ Antibiotics disrupt the commensal microbiota and leave the recipient susceptible to disease, such as CDI.^120-122^ This perturbation can remain detectable up to six months after administration of antibiotics, but the majority of those treated with antibiotics have regained their pretreatment microbiota composition within 4 weeks.^120^ When the microbiota recovers following a course of antibiotics, the new healthy state is not necessarily the same as prior to the administration of antibiotics.^120,121^. Most stool banks use a donor exclusion period of 3 months after antibiotic use ^123^. Prolonged use of proton pump inhibitors also perturbs the microbiota and is associated with an increased risk for CDI.^124,125^ Donors using regular medication are excluded as most non-antibiotic drugs also have extensive impact on the microbiota.^118^

**Recommendation: Active donors receiving antibiotics regardless of indication should be excluded from donation for a period of at least 3 to 6 months. Regular medication use is an exclusion criterion**

*Metabolomic and metagenomic composition*

In general a rich, diverse and abundant microbiota is considered healthy and therefore suitable for donation.^126^ A recent study tried to identify metabolomic and metagenomic factors which could be associated with a higher efficacy in treating rCDI.^127^ A metabolomic and metagenomic analysis of donor stool from 40 unique donors used to treat more than 1400 rCDI patients was performed. Donors were divided into two groups based on their efficacy of curing rCDI (>80% vs 70-80%). Analysis of donor stool did not show a difference in metabolomic or metagenomic profile between these groups.^127^ In addition, Barnes et al. showed that selecting a donor based on microbiota metrics (high diversity, balanced constitution of Bacteriodetes vs Firmicutes, and concentration of fecal butyrate) did not result in a higher cure rate of rCDI with a single infusion.^128^ These results suggest that recipient factors might be more important in curing rCDI with FMT. Interestingly, taxonomic composition of donor microbiota however might play a role in indications other than rCDI, such as ulcerative colitis.^129,130^

**Recommendation: Optimal donor stool selection for the treatment of rCDI based on metabolomic and metagenomic stool profile does not seem feasible yet. Recipient factors seem more important in curing rCDI.**

**Recommendation: Donor selection based on microbiota metrics can be relevant in other diseases than rCDI (e.g. ulcerative colitis).**

**Table 1: Recommended feces screening**

| **Bacteria** | **Test suggestion** | **Exclusion**** |
| --- | --- | --- |
| *Clostridioides difficile* | PCR | Yes |
| *Helicobacter pylori* | Antigen test | Yes |
| *Salmonella* spp. | PCR^1^ | Yes |
| *Campylobacter* spp. | PCR^1^ | Yes, when *C. lari, C. upsaliensis* or *C. fetus* |
| *Campylobacter jejuni/coli* | PCR^1^ | Yes |
| Shiga toxin producing *E. coli* | PCR^1^ | Yes |
| *Shigella* spp. | PCR^1^ | Yes |
| *Yersinia enterocolitica* | PCR^1^ | Yes |
| *Y. pseudotuberculosis* | PCR^1^ | Yes |
| *Aeromonas* spp | PCR^1^ | Yes |
| **Antibiotic-resistant bacteria** |  |  |
| ESBL*/Carbapenemase-producing bacteria | Culture | Yes |
| Vancomycin-resistant enterococci | Culture | Yes |
| Methicillin-resistant *Staphylococcus aureus* | Culture | Yes |
| Other MDRO defined as resistant to both aminoglycosides and fluoroquinolones | Culture | Yes |
| **Viruses** |  |  |
| Norovirus I+II | PCR | Yes |
| Astrovirus | PCR | Yes |
| Sapovirus | PCR | Yes |
| Rotavirus | PCR | Yes |
| Adenovirus 40/41 | PCR | Yes |
| Adenovirus non-40/41 | PCR | Yes |
| Enterovirus | PCR | Yes |
| Parechovirus | PCR | Yes |
| **Parasites**** |  |  |
| *Blastocystis hominis**** | Microscopy (not PCR) | Yes |
| *Dientamoeba fragilis* | Microscopy/PCR | Yes |
| *Giardia lamblia* | PCR | Yes |
| *Entamoeba histolytica* | PCR | Yes |
| *Cryptosporidium parvum* | PCR | Yes |
| *Cryptosporidium hominis* | PCR | Yes |
| *Microsporidium spp* | PCR | Yes |
| *Strongyloïdes stercoralis* | PCR | Yes |

^1:^ if PCR is positive, followed by culture

*: extended spectrum beta-lactamase

**: temporary exclusion. For *Entamoeba histolytica* and *Strongyloides stercoralis,* treatment is needed. For the other indications a rescreening can be performed within 1 – 6 months.

***; preliminary data from the NDFB indicate that FMT of Blastocystes-positive donors determined by PCR do not result in gastrointestinal symptoms or decreased efficacy after transfer to patients. For ulcerative colitis, several data indicate that *Blastocystis hominis* is inversely associated with UC and that successful donors harbor *Blastocystis hominis* far more often than non-successful donors. Confirmation and further studies are necessary to establish the role of screening for Blastocystes in the setting of UC. Until so far, microscopy is used as indicator of a high load, which is recommended as criterion for exclusion.

**Table 2: Recommended serum screening**

| **Pathogen** | **Test** |
| --- | --- |
| Hepatitis A | (IgM +) IgG |
| Hepatitis B | HBsAg + anti-Hbcore |
| Hepatitis C | Anti-HCV |
| Hepatitis E* | (IgM +)IgG |
| HIV | HIV antigen and antibody (HIV-combo test) |
| Lues (*Treponema pallidum*) | TPPA |
| Cytomegalovirus | (IgM +) IgG |
| Epstein Barr Virus | (IgM +) IgG |
| *Strongyloïdes*^1^ | IgG1 + IgG4 |

^1^: If potential donor has a history of travel to Middle and South America, Africa, or Asia

*, In doubt, a PCR on stool (and/or blood) will be performed.

**Table 3: Disorders associated with dysbiosis of the gut microbiota**

| **Gastrointestinal disorders with dysbiosis** | **Additional risk factors of diseases** |
| --- | --- |
| Inflammatory bowel disease | **Or first degree relatives** |
| Irritable bowel syndrome |  |
| Metabolic syndrome / Steatosis hepatis |  |
| Liver cirrhosis |  |
| Microscopic colitis |  |
| Colon carcinoma | **Or first degree relative with colon carcinoma <50 years** |
| Colo- or ileostomy |  |
| **Psychiatric disorders** |  |
| Autism |  |
| Depression |  |
| **Neurinflammatory disorders** |  |
| Parkinson’s disease |  |
| Multiple sclerosis |  |
| **Other** |  |
| Graft-versus-host disease |  |
| Atopy |  |
| Obesity |  |
| Auto-immune disease |  |
| Malignant disease |  |

#### 14.5 Q2 What recipient factors influence the outcome of FMT?

Dr. E. van Nood

Introduction

Recipient factors that influence the outcome of FMT related to selecting and preparing the patient

Ongoing antibiotic use

Correct diagnosis

Whole bowel lavage

Patient characteristics

Recipient factors that influence the outcome of FMT related to infusing the product

Duodenal route

Colonic route

Capsules

Enema

Recipient factors that influence the outcome of FMT related to the product infused

General

Special groups

(anaphylactic) Food Allergies

Celiac disease/Lactose intolerance

Pregnant women

Patients on vasopressive medication/ICU

Patients with decompensated liver cirrhosis

**Introduction**

This chapter deals specifically with recipient factors that can influence the outcome of FMT.

Although relatively simple to perform, questions regarding both short-term and long-term safety as well as the complex and rapidly evolving regulatory landscape have limited widespread use of FMT.[1] Adverse events of FMT have not been well studied. It is therefore even more difficult to identify risks for certain groups. Although there are publications that address donor screening, there are less studies that try to identify factors in recipients that predict a negative outcome.

In a systematic review that included 50 publications, the incidence of adverse events was 29 percent. Of the 78 types of adverse events, the most frequently reported was abdominal discomfort. [2] A total of 44 types of serious adverse events occurred in 9.2 percent of patients. The incidence of serious adverse events among 1089 patients included death, infection, and relapse of inflammatory bowel diseases in 3.5, 2.5, and 0.6 percent, respectively. No specific recipients characteristics can be extracted due to variation between patients. Another study [3] identified procedural adverse events, infectious events, and events per recipient group, but suggests that comparison between patients with such a heterogeneous range of conditions risks to confound true adverse effects with conditions that are part of the natural progression of disease.

As the patients that are treated with FMT have varying underlying conditions, with varying morbidity, it is difficult to distinguish whether events that occur post FMT are true adverse events, or a symptom of the underlying disease (eg in inflammatory bowel disease). This influences clear identification of adverse events, let alone selective recipient identification. Most of the risks are theoretical, as there are limited data on observed side effects, so both the known actual and theoretical risks are evaluated.

With the above mentioned limitations, there are three groups of identifiable (theoretical) recipient factors that can influence the outcome of FMT. Firstly, factors that are related to the actual physical process of infusing feces. These factors mostly affect short term safety. This applies to selection and preparation of the patient and infusion of feces. Recipients who cannot receive proper preparation of FMT, or have a higher chance of experiencing difficulties during infusion of feces have an increased risk of (serious) adverse events, thereby negatively influencing outcome (eg in the case of limited passage of feces in the gastrointestinal tract such as ileus). Secondly, several morbidity factors are identified in recipients that can potentially increase the risk of negative effects of the products infused. Food allergies are one example, and the risk of more severe infection in selected recipient groups (eg a primary CMV infection in an immunocompromised host should be taken into account) together with the risk of other long term side effects. Long term risks relate primarily to potential long term effects on the bowel or the immune system. Thirdly, other recipient factors that can positively or negatively influence the outcome of FMT itself. With this third point there is no true adverse event, but the chance of failure of FMT is increased. For example, ongoing antibiotic use, age, number of admissions prior to FMT can all negatively influence the outcome of FMT, with increased risk of failure.

**1. Recipient factors that can negatively influence the outcome of FMT which are related to the process of selecting and preparing the patient.**

**Ongoing antibiotic use**

An important predictor of failure of FMT (and of recurrence of CDI in general) in several studies is antimicrobial exposure pre-FMT or continuing antibiotic use during or directly following FMT.[4].Therefore, to optimize chance of success, all antibiotics should be stopped prior to infusion of feces. [5] Antimicrobial stewardship after FMT should be implemented to prevent disruption of the new microbiota and development a new CDI.

**Recommendation: Antibiotic use after FMT should be avoided (if possible) due to the increased risk of failure of FMT.**

**Correct diagnosis**

The diagnosis for which the patient receives FMT should be as clear as possible. Testing for CDI is warranted. This is particularly true for recurrent *Clostridium difficile* infection. If patients are misdiagnosed as having CDI, and are given FMT, our experience is that their chance of failure is higher. [6,7] Diagnosis of CDI should be made according to ECCMID guidelines where possible [7].

**Recommendation: FMT should be given to the right patients, therefore proper CDI diagnostics are mandatory. CDI testing should be performed according to ESCMID guideline**

**Whole bowel lavage**

The efficacy of FMT may depend upon the technique used to cleanse the colon before administration of the fecal enema [8]. Historically, feces have been administered to patients by enema or colonoscopy, the latter warranting a whole bowel lavage. This is done by administering a macrogol solution, which is taken the day prior to the infusion of feces. The solution is given orally, and has a total volume of 2 liters. If recipients have an ileus, mechanical obstruction or perforation, a whole bowel lavage is contraindicated. Some macrogol solutions contain aspartame, which is contraindicated in patients with phenylketonuria. Allergy for polethyleenglycol, the basic ingredient of macrogol solutions, is also a contraindication.

**Recommendation:** **Bowel lavage should be administered prior to FMT via the lower gastrointestinal route, and should be considered prior to FMT via the upper route, therefore in case of allergy for substances related to polyethylenic glycol the upper route would be preferred.**

**Patient characteristics**

Several factors have been identified that can negatively influence the outcome of FMT, but might not lead to exclusion of patients. For CDI, previous CDI-related hospitalization is a negative predictor for success in one small study (OR 1.43, 95% CI: 1.18-1.75); with each additional hospitalization, the odds of failure increased by 43%.[5] Furthermore severe and severe-complicated infection and inpatient status during FMT were strongly associated with early failure of a single FMT for CDI. Another study of over 200 patients observed an increased failure rate in female patients (P=0.016), previous hospitalization (P=0.006), and surgery before FMT (P=0.005). [9] However, these are factors that can be observed, but will probably not lead to denying FMT.

**2 Recipient factors that can negatively influence the outcome of FMT that are related to the process of infusing feces.**

Several routes of administration of fecal intestinal microbiota have been reported, (but the optimal protocol for FMT is unclear and probably both routes are comparable. [10,11,12] A pooled analysis of 182 cases of rCDI treated with FMT showed that colonoscopic FMT has a slightly higher cure rate than nasogastric FMT (93 versus 85 percent), although the difference was not statistically significant [13] . Both routes are used depending on patient clinical characteristics, but the majority of patients treated for rCDI in the Netherlands are given FMT through the upper gastrointestinal route. [12]

**Recommendation: Recipients should be evaluated for the optimal route of receiving FMT. If the upper gastrointestinal route is not feasible, the colonoscopic route can be chosen and vice versa.**

**Upper gastrointestinal/duodenal route.** Feces can be given through a duodenal tube. If potential recipients of feces are dealing with passage problems of the upper gastrointestinal tract (fistulas, perforation, ileus) infusion of feces using the duodenal route is not feasible. If patients are nauseated or prone to vomit, or if the duodenal tube cannot be positioned appropriately, the lower route is be preferred.[14,15] With regard to placement of duodenal tube in recipients there are only few contraindications. If placement of duodenal tube using mild sedation (eg midazolam) is preferred, general criteria and protocols should be provided. Patients with serious cardiac or pulmonary conditions should not be given sedation unless protocols as developed in the hospital are respected.

**Recommendation: Patients with known obstruction of the gastrointestinal tract (eg ileus) should not receive FMT through the upper gastrointestinal route.**

**Colonic route** Colonoscopy must be performed cautiously to minimize the risk of perforation. (16; 17) If the colonic route is chosen, all patients with already existing perforation should be excluded. Patients with severe colitis should be identified to take extra caution, in order to prevent perforation. Most patients who undergo colonoscopy also receive mild sedation (eg midazolam), for which the local protocols apply.

| **Patient category** | **Drawback/contraindication** | **consequence** |
| --- | --- | --- |
| Ileus | Hampered passage of whole bowel lavage/feces | Only rectal |
| Perforation | Intra-abdominal spill of whole bowel lavage/feces | No FMT |
| Nausea/vomiting | Increased risk of vomiting if upper GI tract is used for infusing feces | Only rectal |
| Serious lung/cardiac problems | Potential complications of sedation | If sedation warranted with local protocols |
| Allergy macrogol/polythyleenglycol |  | No whole bowel lavage |
| PKU | Macrogol with aspartame not to be given | Whole bowel lavage without aspartame |
| subtotal colectomies | Less effective, even in CDI |  |

**Recipient factors that can negatively influence the outcome of FMT that are related to the infused product.**

Patients with specific characteristics (old age, immunocompromised state, decompensated liver cirrhosis, pregnancy etc) all have their own (most theoretical) risks.

Literature is limited for most groups, although data on immunocompromised patients is steadily increasing. If recipients are more likely to develop side effects or adverse events following FMT this might not implicate that FMT should not be given. As mentioned earlier, patients who have had several recurrences of CDI seem to have a higher chance of FMT failure. But this particular group of patients is also far less likely to respond to any other possible therapy. Therefore, if the risk is acceptable, FMT should still be considered

**Special groups**

**Immunocompromised patients**

FMT was not widely used in immunocompromised patients at first, owing to concern for donor-derived infection. In the last 10 years however, the group of patients who are given FMT is steadily growing, both because rCDI occurs more often in patients with solid organ transplants, and because there is growing interest in influencing dysbiosis and influencing graft-versus-host-disease in hematologic patients. A smaller retrospective study compared outcome and adverse effects between immunocompetent and immunocompromised patients. It describes the absence of increased risk of adverse events. The predictor of failure in their study was antimicrobial exposure pre-FMT [4]. Most studies are small. In a retrospective analysis of FMT in 99 immunocompromised patients, only a few SAEs or related adverse events were observed. [18] .With the increased incidence of FMT the data suggesting that FMT in immunocompromised patients is safe are vastly growing, but again mainly limited to case reports and cohorts. Side effects vary from small intestinal bacterial overgrowth [19], to infectious complications and transient side effects that are also observed in the immunocompetent group. On June 13, 2019, the Food and Drug Administration (FDA) issued a safety alert concerning the risk of serious adverse reactions due to transmission of multi-drug resistant organisms (MDRO) through FMT to two immunocompromised patients. [20] One of the individuals died, but the report did not provide information on the cause of death. For reasons not specified, the donor had not been screened for MDRO. The FDA required inclusion of MDRO screening in all active and future FMT-based study protocols. Although the NDFB screens regularly for MDRO and only releases fecal suspensions after a quarantine period, the NDFB decided to slightly adapt the protocol and to use only feces suspensions for treatment of severe immunocompromised patients that have been screened directly for MDRO and other microorganisms according to the existing protocol. The precise status of immunocompromised patients will be determined by the expert group of the NDFB when a FMT is requested. A separate protocol has been made and is in accordance with a recent proposal of UEG, supervised by prof. Vehreschild (Cologne). For FMT studies in other diseases than rCDI, a special recommendation should be made by a group of experts including members of the NDFB.

**Recommendation: FMT should be offered with caution to immunosuppressed patients, in whom FMT appears efficacious without significant additional adverse effects.**

**Norovirus transmission** possibly associated with FMT has been reported in two cases, both of which were positive for norovirus on PCR. Although one donor had a negative test, and the other donor was not tested at all, there was a correlation in time. [21] These patients were not immunocompromised. However, norovirus can give a more serious clinical course in immunocompromised patients, and therefore extra caution should be undertaken when giving FMT to this group of patients during norovirus season.

**Gram-negative bacteraemia** occurred in several cases after FMT [22] with two of the patients dying. *Escherichia coli* bacteraemia occurred 24 h after colonoscopic FMT in a 61-year-old man with concomitant Crohn’s disease and diverticulitis who had had six *prior E. coli* bacteraemias in the preceding three and a half years. The authors postulated that altered intestinal permeability was the cause. Another report mentions a patient who died 48 hours after FMT for refractory CDI with toxic megacolon and shock with positive bloodcultures of *Pseudomonas aeruginosa, Eschericha coli,*and *Lactobacillus casei.* [23]*.* Following this SAE, the authors modified the FMT consent form to include the possibility of post-FMT colitis, sepsis, and death.

**Other viral infections**, such as **CMV and occasionally EBV** can be transmitted through FMT. [24] In immunocompromised recipients who are CMV naïve, infection can lead to a primary CMV infection, with potential deleterious consequences. It is therefore advised to match CMV in donor and recipient, in order to prevent adverse events.

**Recommendation: If FMT is administered to an immunocompromised recipient, standard protocols should deal with CMV (and occasionally EBV) status of donor and patient.**

**(anaphylactic) Food Allergies**

If recipients have anaphylactic allergic reactions in their previous medical history, our advice is not to use FMT. The responsibility of delivering donor feces that is 100% clean of the allergen, which can act as a potential lethal product for the recipient cannot be accepted. In milder allergies, FMT can be given with extra focus on possible avoidance of products that cause allergy. A case of ‘hives’ occurred in a patient with history of medication allergies during the seven-day follow up period after colonoscopic FMT of anonymous donor faeces. [25]

**Recommendation; FMT should not be offered to recipients with a history of anaphylactic food allergies.**

**Celiac disease/Lactose intolerance**

Recipients with known other food allergies, celiac disease or lactose intolerance can receive FMT. A donor can be selected who is willing to eat gluten free for some time, in order to deliver feces that has no additional risk for exacerbation of underlying disease.

**Recommendation; FMT can be offered to patients with celiac disease or known lactose intolerance or mild food allergies; special donor preparations can be considered.**

**Pregnant women**

To our knowledge only one case is described in which a pregnant woman received FMT. [26] With uncertainties on the effect on pregnancy we believe that FMTs should not routinely be offered to pregnant women.

**Recommendation; FMT should preferably not be given to pregnant women.**

**Patients in ICU or with refractory severe CDI**. Recurrent CDI or refractory CDI in the intensive care unit (ICU) has been treated with FMT in a limited number of patients. [22, 27] Although there is limited data with the risk of publication bias, the results appear favourable. With the increase in interest for dysbiosis in ICU, several patients with conditions other than CDI have been treated with FMT. [28] A large retrospective French analysis amongst 111 patients with severe CDI, revealed that early FMT dramatically reduces mortality and should be proposed as a first-line treatment for severe CDI. [29] Further studies are needed to clarify complications and contraindications. We would advise to use caution in performing FMT in the ICU or for patients with severe refractory CDI.

**Recommendation; FMT should be given with caution to patients with refractory severe CDI or patients in ICUs.**

**Decompensated liver cirrhosis**: In patients with advanced cirrhosis on lactulose and rifaximin, FMT restored antibiotic-associated disruption in microbial diversity and function. [30] However, there is an increased risk of translocation in patients with ascites, which warrants caution.

**Recommendation:** **There is no evidence that FMT is not safe in patients with liver cirrhosis. However, FMT should be given with caution in patients with decompensated chronic liver disease.**

**Children:** The role of fecal microbiota transplant (FMT) in the treatment of pediatric inflammatory bowel disease (IBD) or irritable bowel syndrome (IBS) is unknown though it is considered as an effective and safe treatment for children with rCDI. [31, 40, 41] One study showed twenty-one subjects who received a single FMT for active IBD, with a median age of 12 years, of whom 57% and 28% demonstrated clinical response at 1 and 6 months post-FMT, respectively. Adverse events attributable to FMT were mild to moderate and self-limited. [32] In a phase 1 pilot study, 10 children and young adult patients (aged 7 to 21 years) with mild to moderate UC received fresh fecal enema daily for five days. [33] At baseline, pediatric UC activity index (PUCAI) ranged from 15 to 65. Clinical response (>15 reduction in PUCAI) within one week occurred in seven of nine (78 percent) children, including three (33 percent) who had clinical remission (PUCAI <10) and six (67 percent) who maintained clinical response at one month. As compared with baseline, median PUCAI significantly improved after FMT. There were no adverse events.

**Recommendation: FMT is a safe and effective treatment for children with rCDI.**

**Older people:** In a case review of all FMT recipients aged 65 or older, mortality was high, but FMT was not a causative factor in these events. [34].

**Recommendation: There is no reason to withhold FMT for the elderly population**.

**Patients with active IBD:**

Smaller studies looking at safety mostly address short term safety in patients with ulcerative colitis [35] or Crohn’s disease, and most studies do not report on serious adverse events in study periods that vary from weeks to months. [36] In the larger studies no real adverse events were noted [37 and 38]. There is some concern that use of FMT in inflammatory bowel disease can paradoxically increase disease activity, mainly seen in patients with Crohn’s disease. This limited experience suggests that FMT may cause overstimulation of the immune system leading to a flare of the IBD. [39] In patients with concomitant IBD and CDI (where FMT was administered primarily for CDI), clinical deterioration occurred in six cases.[3]. However, a beneficial effect of FMT was described in several studies addressing the effects of FMT for ulcerative colitis. [37,38] In general, FMT in patients with ulcerative colitis and CDI appears safe and effective. Whether patients should be pretreated with eg prednisolone in combination with vancomycin, or should receive upfront FMT is not known. Current studies are focusing on identifying a favorable microbiota composition in donors used to treat IBD via FMT. Ideally, if such a donor could be identified, it would also be the preferred donor to treat rCDI in IBD patients.

**Recommendation: With lack on data on the optimal protocol for FMT for IBD, FMT is preferably given in research setting.**

| **Patient category** | **Potential drawback** | **consequence** |
| --- | --- | --- |
| Immunocompromised patients | Increased risk for infection | Screen and match |
| Anaphylactic food allergy | Anaphylactic reaction | No FMT |
| Food allergy | Mild | Consider to instruct donor |
| Preexistent celiac disease | Exacerbation celiac disease | Consider gluten free donor |
| Preexistent lactose intolerance | Exacerbation lactose intolerance | Consider lactose free donor |
| Pregnant patients | Unknown effect of FMT on child | Depending on underlying condition |
| Decompensated liver disease | Potential translocation |  |
| Children | Unknown long term effects | With caution |
| Elderly | More comorbidities | With caution |
| Active IBD | Flare IBD |  |
| Severe disease | translocation | With caution |

16. L Potakamuri, L Turnbough, A Maheshwari, S Kantsevoy, A Ofosu, Effectiveness of Fecal Microbiota Transplantation for the Treatment of Recurrent *Clostridium difficile* Infection: Community Hospital Experience: Presidential Poster

American Journal of Gastroenterology 108, 2013

17. Kelly C, de Leon L Successful Treatment of Recurrent *Clostridium difficile* Infection with Donor Stool Administered at Colonoscopy: A Case Series, , American Journal of Gastroenterology: October 2010 - Volume 105 - Issue - p S135, Abstracts: COLON

18. Kelly CR, Ihunnah C, Fischer M, Khoruts A, Surawicz C, Afzali A, Aroniadis O, Barto A, Borody T, Giovanelli A, Gordon S, Gluck M, Hohmann EL, Kao D, Kao JY, McQuillen DP, Mellow M, Rank KM, Rao K, Ray A, Schwartz MA, Singh N, Stollman N, Suskind DL, Vindigni SM, Youngster I, Brandt L. Fecal microbiota transplant for treatment of *Clostridium difficile* infection in immunocompromised patients. Am J Gastroenterol. 2014 109:1065-71

19. Webb BJ, Brunner A, Ford CD, Gazdik MA, Petersen FB, Hoda D. Fecal microbiota transplantation for recurrent *Clostridium difficile* infection in hematopoietic stem cell transplant recipients.Transpl Infect Dis. 2016;18:628-33.

20. https://www.fda.gov/vaccines-blood-biologics/safety-availability-biologics/important-safety-alert-regarding-use-fecal-microbiota-transplantation-and-risk-serious-adverse

21. Schwartz M, Gluck M, Koon S. Norovirus gastroenteritis after fecal microbiota transplantation for treatment of *Clostridium difficile* infection despite asymptomatic donors and lack of sick contacts. Am J Gastroenterol. 2013;108:1367

22. Trubiano JA, Gardiner B, Kwong JC, Ward P, Testro AG, Charles PG. Faecal microbiota transplantation for severe *Clostridium difficile* infection in the intensive care unit. Eur J Gastroenterol Hepatol. 2013;25:255-7.

23. Paola R. Solari, Patrick G. Fairchild, Luis Junco Noa, Mark R. Wallace. Tempered Enthusiasm for Fecal Transplant. Clin Infect Dis. 2014;59:319.

24. Hohmann EL, Ananthakrishnan AN, Deshpande V. Case Records of the Massachusetts General Hospital. Case 25-2014. A 37-year-old man with ulcerative colitis and bloody diarrhea. N Engl J Med. 2014;371:668-75

25. Kahn SA, Goeppinger SR, Vaughn BP, Moss AC, Rubin DT, Tolerability of Colonoscopic Fecal Microbiota Transplantation in IBD, Gastroenterology, May 2014 Volume 146, Issue 5, Supplement 1, Page S-581, Mo1187

26. Saeedi BJ, Morison DG, Kraft CS, Dhere T. Fecal Microbiota Transplant for *Clostridium difficile* Infection in a Pregnant Patient. Obstet Gynecol. 2017;129:507-509. .

27, You DM, Franzos MA, Holman RP. Successful treatment of fulminant *Clostridium difficile* infection with fecal bacteriotherapy. Ann Intern Med. 2008;148:632-3.

39. Wurm P, Spindelboeck W, Krause R, Plank J, Fuchs G, Bashir M, Petritsch W, Halwachs B, Langner C, Högenauer C, Gorkiewicz G. Antibiotic-Associated Apoptotic Enterocolitis in the Absence of a Defined Pathogen: The Role of Intestinal Microbiota Depletion. Crit Care Med. 2017;45::e600-e606.

40. Davidovics ZH et al. *Clostridium difficile* Infection and Other Conditions in Children: A Joint Position Paper From the North American Society for Pediatric Gastroenterology, Hepatology, and Nutrition and the European Society for Pediatric Gastroenterology, Hepatology, and Nutrition. J Pediatr Gastroenterol Nutr. 2019 Jan;68(1):130-143.

41. Barfield E, Small L, Navallo L, Solomon A. Going to the Bank: Fecal Microbiota Transplantation in Pediatrics. Clin Pediatr (Phila). 2018 Apr;57(4):481-483.

#### 14.6 Q3: Where and under which conditions should  donor stool samples (fresh or frozen) be processed and stored

dr P.P. H le Brun

As long as the status of FMT is not defined, production and quality control are based on the GMP for non-sterile production. This includes protocols for fresh and frozen donor feces, since many studies still use fresh donor feces (1). The following conditions are based on the current EU GMP.

**Production:**

Standard operating procedures for collection and processing are available. All handling of materials and products, such as receipt and quarantine, sampling, storage, labelling, dispensing, processing, packaging and distribution are done in accordance with written procedures or instructions and, where necessary, recorded. Fresh stool samples will be processed within 2 hours after delivery. Incoming materials are checked and labelled and put in quarantine. Contamination of a starting material or of a product by another material or product is prevented.

Materials are registered with a data-system for coding, registration, tracking and tracing of the samples and faecal suspensions. A storage time will have to be defined based on experimental data.

All products are labelled with the identity and a unique code, traceable to the donor. Further shelf life and storage condition are part of the label

**Personnel:**

A proper job description is described including competences training and re-training. The different duties for production, quality control and release are described. Personnel is also trained in hygiene including hand wash procedure. No person affected by an infectious disease or having open lesions on the exposed surface of the body is engaged in the production process. Eating, drinking, chewing or smoking, or the storage of food, drink, smoking materials or personal medication in the production and storage areas is prohibited. Direct contact is avoided between the operator’s hands and the exposed product as well as with any part of the equipment that comes into contact with the products.

**Premises and equipment:**

Processing is done in a controlled but not classified facility. The lay out is preferably in accordance with GMP class D with BSL-2 facilities. Lay out of the room is such that cross contamination is prevented and in such a way as to allow the production to take place in a logical order corresponding to the sequence of the operations. Entrance of unauthorised personnel is prevented. Walls and floor are smooth and easy to be cleaned. There is a gowning area for personnel including a gowning procedure. Premises are cleaned with soap and a sporicidal disinfectant after individual stool processing. As much as possible disposable materials will be used or otherwise autoclavable materials.

For storage of faecal suspensions a storage at -80°C, in a freezer with connected alarm notification in a room separate from the processing area, is mandatory. Storage areas are of sufficient capacity to allow orderly storage of the various categories of materials and products: starting and packaging materials, intermediate, bulk and finished products, products in quarantine, released, rejected, returned or recalled.

**Quality control:**

In process checks and a QC release procedure for original stool AND faecal suspension are mandatory. Quality Control is concerned with sampling, specifications and testing and release procedures. All procedures are described in a quality manual.

1. Lai CY, Sung J, Cheng F, Tang W, Wong SH, Chan PK, Kamm MA, Sung JJY, Kaplan G, Chan FKL Ng SC. Systematic review with meta-analysis: review of donor features, procedures and outcomes in 168 clinical studies of faecal microbiota transplantation. Aliment Pharmacol Ther. 2019 Feb;49(4):354-363.

#### 14.7 Q 4: What factors related to the preparation of the transplant influence the outcome of FMT?

Drs. E.M. Terveer, drs. B. Rethans, prof. C Ponsioen, prof. M Benninga, prof. H. Verspaget, dr. Josbert Keller, prof. Ed Kuijper

**Disclaimer**: The general steps recommended in this statement are based on what has been described, but never rigorously tested. There are no reported studies comparing different preparation protocols of fecal suspensions, but the protocols used in different studies are comparable and allow good/moderate evidence of suitable protocols for preparation of fecal suspensions for FMT treatment of recurrent *Clostridioides difficile* infection (rCDI).

In line with the Good Manufacturing Practices (GMP) as defined by the WHO, donor feces collection and preparation for FMT should follow a standard protocol to ensure “that products are consistently produced and controlled to the quality standards appropriate to their intended use and as required by the marketing authorization”

**Recommendation. Donor stool collection and preparation for FMT should follow a standard protocol.**

Stool collection

- There is very little evidence or guidance for the collection of donor feces. To promote standardised practice and a safe and effective product, clear (preferably written) instructions should be provided to the donor for feces collection and delivery procedures.
- To prevent environmental/cross-contamination, feces is collected by the donor in a fecal container (e.g., Fecotainer).
- Until further processing/handing the stool sample can be stored at room temperature (0°C–30°C). If this takes more than 30 min, temporary storage in a cooler bag or refrigerator is preferred. Research showed that fecal storage without stabilisation buffer significantly changes taxa abundances from 30 minutes onwards ^131-134^.

**Recommendation. Stool should be collected in a clean container and stored at room temperature for no longer than 30 min. If longer, a cooler bag or refrigerator should be used.**

Timeline of processing the feces to a fecal suspension

- It is generally believed that a high viability of bacteria in stool increases the chance of a successful FMT. As the majority of fecal bacteria are anaerobic, feces should be processed as soon as possible to minimise sample degradation and alteration over time, which may occur due to the complex metabolic and environmental requirements of the fecal microbiota.
- **A period of 6 hours** has been generally applied across many successful studies of FMT treatment in (r)CDI and randomized controlled trials (RCTs) in particular ^42-44,72,97,135-151^. Although no formal comparative study has been performed, in studies which use a longer period between collection and processing ^76,152-154^ (i.e., processing feces within 24 to 48h), the cure rate of FMT seems lower than in studies where processing of feces is performed within a short time interval (within 8h), with cure rates of 74% and 85%, respectively. For other indications, such as the treatment of Inflammatory Bowel Disease (IBD) or Irritable Bowel Syndrome (IBS), there are no firm data. It may, however, be that processing time for these indications is more critical. Because this is not yet known, we do not recommend a different processing time, this might change in the future.
- As the preparation of donor feces takes time, it is advised to donors to submit their feces **within 2 hours after defecation**.

**Recommendation. Stool should be processed to a fecal suspension within 6 hours.**

(An)aerobic conditions of fecal preparation

- There are no comparative trials of anaerobically versus aerobically prepared FMT for treatment of rCDI, IBD or IBS. The vast majority of fecal suspension preparations has been undertaken aerobically. Three small observational studies (n=86) have been performed with anaerobic processing of the feces ^155-157^, with a rCDI cure rate of 80% (all studies taken together, first infusion). This is not significantly different from the cure rate of the standard aerobic processing, with cure rates of 76% ^106,155-157^ (first infusion). Therefore, for rCDI there appears to be no clear need to process donor feces anaerobically, a method which introduces additional complexity and costs.
- The discrepancy between infusing healthy microbiota, which consists largely of anaerobic bacteria, and aerobic processing of feces could be due to the fact that a considerable part of the bacterial genera produce resilient spores allowing interindividual transfer of at least a proportion of oxygen-sensitive intestinal bacteria ^158^. Given that these spore-forming bacteria typically represent about one-third of gut bacteria ^158^ and that disorders accompanied by dysbiosis, such as IBS or IBD, are typically defined by lower abundance of anaerobic bacteria ^159-161^, it provides rationale to expect that the anaerobic processing of samples could be relevant for FMT success in the treatment of these disorders. However, at present data are too scarce to recommend a strict anaerobic protocol for processing donor feces for the treatment of IBD.

**Recommendation. Aerobically and anaerobically prepared fecal suspensions are both considered suitable for FMT**

Amount of feces

- Most RCTs and case series report variable amounts of stool used for preparation of a fecal suspension. The majority of studies use ≥ 50 gram of feces. Two systematic reviews and one study in IBS patients recommend the use of ≥ 50 gram of feces ^73,74,162^ since decreased cure rates were observed when using < 50 gram. Gough et al., observed a fourfold increase in recurrence rates if < 50 gram of stool was used ^73^. Yet, this report was published already in 2011. Moreover, the second systematic review concludes this recommendation based on 2 case-series (with capsules) ^74^.
- We performed also an analysis of cure rates of all rCDI studies reporting the fecal amount used; studies which use less than 50 gram (most use ≥ 30 gram) reported a cure rate of 82% (404/505, 12 studies). When more than 50 gram was used a cure rate of 86% was observed (964/1118, 25 studies).
- Concerning cost-effectivity, use of 30 gram of feces could be considered since many experts use 30 gram with good clinical results.

**Recommendation. It is preferred to use approximatey 50 g of stool to prepare a fecal suspension for rCDI treatment.**

Diluent for feces to prepare fecal suspension

- A comparative study with different fecal suspension diluents has not been performed. The majority of studies have used preservative-free sterile 0.9% saline as diluent for processing feces for FMT suspensions ^42-44,72,75,106,136,138-141,143-146,149,150,152-157,163-176^. Some studies, however, also use fresh water ^97,137,142,177^, with similar cure rates (success rate first infusion, transfer via enema excluded) of 89% (4 studies) versus 83% when saline was used. If enemas are included in the analysis of water-processed FMT suspensions a cure rate of only 58% (7 studies) was observed ^76,137,142,177-179^. It is unclear if this drop in cure rate is only caused by the transfer via enema (which is a less effective method, and often needs repeated enema transfers), or by the use of water in the feces processing. Theoretically saline should be superior to water as saline enables better preservation of microorganisms ^180^.
- The initial volume of diluent used to process the fecal suspension varies between studies, with ranges in ratio ‘feces to diluent’ from 1:1 to 1:10. A clear difference in outcome between the different ratios is not observed. When a lower dilution factor 1:1 – 1:6.7 is compared with a higher dilution of 1:10, cure rates (success rate first infusion, transfer via enema excluded) of 84% and 80%, are respectively observed.
- The amount of diluent depends on the route of administration, as the total amount of fecal suspension for the upper GI route is usually less (<200 ml) than the lower GI route (200-500 ml). In addition, a smaller amount of diluent maximises the amount of feces in the fecal suspension (and bacteria/ml). The suspension should, however, not be too viscous to be able to deliver via a naso-duodenal tube or biopsy channel of a colonoscope. Therefore, the optimal balance between above considerations is a ‘feces : diluent’ ratio of 1 : 3 to 5. (expert opinion)

**Recommendation. Sterile 0.9% saline should be used as diluent to prepare the fecal suspension.**

**Recommendation. To prepare the fecal suspension a ‘feces to diluent’ of 1 : 3-5 should be used.**

Homogenisation and filtration of the fecal suspension

- Fecal suspensions can be homogenised by a variety of methods such as in blenders ^43,106,138-140,145,146,148,149,152,155,156,163,164,167,169,174,175^, in stomacher bags ^136,153,172^, with mortar and pestle ^105^, or with wooden spatulas ^76,147,178^, with no apparent major variation in efficacy. Of utmost importance is the use of **sterile or clean material**, which implies that all material should be autoclaved or disposable. Possible disadvantages of blenders are difficulties with appropriate sterilisation and possible aerolisation of the feces suspension.
- To prevent clogging of the tube/biopsy channel during the administration procedure the fecal suspension should be filtered. Filtration can be performed by a gauze, filter paper, strainers or sieves. To prevent external contamination either a closed system or an open system in a flow cabinet should be used.
- When infusing the suspension via colonoscopy or enema a filtration step is not needed if the fecal suspension is homogenised in a blender. A possible theoretical advantage of unfiltered feces is the preservation of fibrous material, as many short-chain fatty acid producing colonic bacteria require fibre as substrate ^181^. In clinical studies regarding treatment of rCDI, no disadvantage of fecal suspensions that are homogenised by blender versus other methods is proven.
- 15 studies report using a blender, with a combined cure rate of 84% (820/973), 14 studies report using other methods with a combined cure rate of 70% (528/755). Leaving out studies which use enema as delivery mode, the cure rates were comparable, 84% and 89%, respectively.
- To reduce the infused volume concentration, concentration by centrifugation is allowed. Many studies, especially when preparing a fecal suspension for the upper GI, use a centrifugation step without effect on the outcome. An exception is one study that used multiple centrifugation and washing steps and showed a markedly low cure rate of 68% ^144^.

**Recommendation. Feces should be homogenized and the suspension filtered when applied via the upper GI route. Of utmost importance is the use of sterile or clean material, which implies that all material should be autoclaved or disposable.**

Fresh or Frozen and storage period

- Two RCTs and one meta-analysis showed non-inferiority and comparable cure rates for the treatment of rCDI with fresh or frozen (-80**°**C) fecal suspensions ^75-77^. Use of a frozen fecal suspension allows storage for a longer period of time until the donor has been retested prior to actual use of the fecal suspension. This lowers the risk of transferring diseases by bypassing the window of detection phase of some transmissible infections (e.g. HIV, Hepatitis C). In addition, having well-screened donor fecal suspensions in storage will allow more rapid transplantation when needed, bypassing the logistical difficulties of screening and preparing a fresh FMT suspension.
- Storage at -80^o^C rather than at -20^o^C is recommended to minimise sample degradation.
- When a frozen fecal suspension is prepared, an appropriate cryoprotectant should be added prior to freezing. Cryopreservation is a process of preservation of the biological and structural functions of tissues or cells by cooling to sub-zero temperatures. This minimises the risk of cellular damage from intracellular freezing and protects cells against slow-cooling (solution effects) injury ^182^. In most studies the cryoprotectant **glycerol** is used for FMT preparation in a final concentration of 10 to 15% ^97,106,136,144,147,155,156,164,168,172,174,175,183^. Viability of six representative groups of fecal bacteria after 2 months of storage at -80°C in normal saline with or without 10% glycerol did not differ from baseline. However, at 6 months the aerobes, total coliforms and lactobacilli were significantly reduced by >1 log ^156^ in the fecal suspensions stored without glycerol.
- Clinical success of frozen fecal suspensions is reported after up to 6-10 months of storage at -80°C ^97,105,136,147,155,156,174,175^, but this could in theory be much longer. However, there have been no comparative clinical trials investigating storage duration. OpenBiome and the NDFB have positive experiences with storage of up to 2 years ^184^**.** Material stored for < 6 months (83.8%, N=1473) was comparable in effectiveness to material stored for 6-12 months (83.8%, N=439) and for >12 months (83.3%, N=12), suggesting that frozen storage duration does not significantly impact the rate of clinical cure ^184^.
- To ensure the maximum safety and quality of the fecal suspension, it is mandatory to specify a maximum storage time with an expiry date.
- A side effect of large amounts of glycerol in the bowel is a mild alteration in serum glucose. This is not observed when less than 0.75 gr glycerol /kg body weight is used. For a person weighing 70 kg, this results in a glycerol limit of 52.5 gram (approximately 500 ml FMT suspension). A calculation of the maximum fecal suspension volume and possible adjustment should be made when infusing large volumes to diabetic patients with low weight.
- A potential side effect of glycerol is its laxative effect.

**Recommendation. The use of banked frozen fecal suspensions (-80°C) is considered preferable to fresh preparations.**

**Recommendation. Glycerol at a final concentration of 10% should be added to a fecal suspension prior freezing.**

**Recommendation.** **Fecal suspensions stored at -80°C appear safe and effective up to a shelf life of 24 months. A date of expiration should be registered on the product.**

Thawing of donor feces suspension

- There are little published data addressing optimal thawing of frozen fecal suspensions. Warm water baths (37°C) have been recommended to speed thawing ^123^. However, this may introduce risk of cross contamination by *Pseudomonas* species from the water bath and may reduce bacterial viability of the fecal suspension. Thaw the fecal suspension overnight in a 4°C refrigerator or during 5 hours at room temperature (for 200 ml suspensions). Thaw times vary related to fecal suspension volume.
- After thawing, saline could be added if necessary to obtain a desired suspension volume.
- The fecal suspension should be at room temperature while infusing into the recipient in order to avoid ‘cold shock’. Depending on the volume of fecal suspension administration will take 15 to 60 minutes (recommended transfusion rate is 10 ml per minute).
- Once thawed, fecal suspensions should not be refrozen. Freeze-thaw cycles adversely affect the viability of the microbial communities in the fecal suspension ^185^.

**Recommendation. Thawing of FMT suspensions at ambient temperature or overnight in the refrigerator is preferable over warm water baths.**

**Recommendation. Thawed FMT suspensions should be infused the same day, and should NOT be refrozen.**

Pooling of donor feces

- Pooling (mixing) of multiple donor feces during processing is not recommended. Firstly, it hampers the traceability of the fecal suspension to the individual donor and risk on transmissible disease may be increased. Secondly, the principle of transfusing a well (characterized and) balanced microbiota suspension might be lost. The pooled microbiota of different donors might even be antagonistic to each other. The efficacy of an FMT with pooled fecal suspensions is not known.

**Recommendation. Pooling of donor feces during processing is not recommended.**

#### 14.8 Q 5: How should FMT be administered to patients?

Dr. Josbert Keller, prof. Dr. Cyriel Ponsioen, prof. Dr. Marc Benninga, dr. Els van Nood, dr. Bram Goorhuis, prof. Dr. Chris Mulder

**What is the preferred route of administration?**

There are 5 different methods for instillation of donor feces suspension/microbiota in patients:

1. The nasoduodenal route is effective, well tolerated and generally safe. The cure rate after one single infusion is > 80%. To date, there is no evidence that small intestinal bacterial overgrowth (SIBO) is induced by upper GI FMT. Adverse events appear to be uncommon, mild and self-limiting; although serious adverse events including bacteraemia, perforations and death have been reported. [1,2] Especially regurgitation, vomiting and aspiration have been described after FMT [2,3,4] by the duodenal route. For this reason, care should be taken in patients with impaired gastrointestinal motility, and the suspension needs to be infused slowly.

Alternatively, the suspension can be infused during gastroscopy in the duodenum of patients. Rapid infusion and larger volumes may increase the risk of regurgitation.

In one patient, aspiration pneumonia and subsequent death was described after general anesthesia and infusion during gastroscopy. [2] Preferably, general anesthesia during FMT should be avoided.

2. FMT by colonoscopy appears equally effective as by duodenal infusion. There are no studies directly comparing the two methods. FMT by colonoscopy is safe, but may be demanding in (fragile) patients. [5]
3. Donor feces suspensions can be administered by enemas. This method appears less effective, but repeated infusions may be required. [8].
4. Capsules containing donor feces (suspension) appear effective and promising. [6,7,8] Not all capsules necessarily contain lyophylized microbiota, frozen preparations have also been shown to be effective. However, a recent meta-analysis on the effect of FMT in IBS demonstrated a clinical benefit of FMT using nasojejunal tubes, but no clinical benefit of FMT capsules. [13] Capsules are often large, and swallowing large numbers of capsules (e.g. 30 capsultes) in a single day may be a significant undertaking for certain patients. Newly produced capsules should be tested in a clinical study before implementation in daily practice.
5. FMT using nasogastric tube for delivery of feces suspensions has been described in a few patients after prescription of a proton pump inhibitor. [9] We do not recommend this route of instillation, because of the potential risk of regurgitation of the donor feces suspension.

**Recommendation: FMT appears generally safe and effective if administered by nasoduodenal tube or colonoscopy. In patients with (suspected) impaired GI motility, colonoscopy is the preferred route. In fragile patients, colonoscopy may preferably be avoided. The primary cure rate of enemas seems lower, and this route is generally not advised.**

**What is the preferred volume of donor feces suspensions?**

Initially, large volumes of donor feces suspensions were used [10], these appeared effective and safe. However, later studies showed that regurgitation, vomiting and aspiration after FMT using the duodenal route may occur. [2, 3] The NDFB has therefore reduced the volume of donor feces suspensions to < 200 cc (198 cc). This appears safe, if precautions are taken (slow infusion of donor feces suspensions). A restricted volume of the suspension appears unnecessary if FMT is administered by colonoscopy.

**Recommendation: Larger volumes should be avoided if FMT is administered by the nasoduodenal/nasogastric route. The results of the NDFB suggest that a donor feces suspension of up to 200 cc is safe, if precautions are taken.**

**Is bowel lavage required before FMT:**

Bowel lavage is always prescribed before colonoscopy [11], and is generally also prescribed before FMT administration using the upper GI route. [3,10,12] It is not known if bowel lavage is required before donor feces infusion. Given the excellent results of FMT after bowel lavage using polyethylene glycol preparations, it is generally prescribed. However, FMT can be considered without bowel lavage as well.

**Should prokinetics, PPI, or loperamide be administered before or after FMT?**

There is no evidence that PPI, prokinetics or loperamide can improve efficacy or safety of FMT.

**Recommendation: Prokinetics can be administered if patients experience nausea after infusion of donor feces suspension via a nasoduodenal tube. PPI’s should be given prior to FMT if the donor feces suspension is administered using a nasogastric tube (which is generally not advised as route of administration).**

**Should antibiotics be administered prior to FMT? When should antibiotics be stopped before instillation of donor feces suspensions (washout period)?**

In general, antibiotics with activity against *C. difficile* are prescribed before FMT for patients with rCDI to eradicate *C. difficile* and to increase engraftment. The necessity of pretreatment for other diseases is unknown. Also, patients need to be treated in the “waiting time” before FMT is scheduled. Although there is no evidence pointing to better outcome due to pre-treatment with antibiotics, it seems reasonable to initiate treatment with antibiotics against *C. difficile* immediately after a positive *C. difficile* test.

**Recommendation:** **In general, vancomycin 125-250 mg qid, or fidaxomicin 200 mg tid should be administered during at least 4 days before donor feces infusion. [10]**

To minimise the deleterious effects of antibiotics on the donor (FMT) microbiota, a minimum washout time of 24 hours is required.

**Which infection prevention measures should be undertaken?**

Local infection prevention protocols should be followed to prevent transmission of

*C. difficile* to other patients while administering FMT to patients with CDI.

#### 14.9 Q 6: What is the general approach for follow-up after FMT?

Prof. dr. Ed.J. Kuijper, prof. dr. H. Verspaget, prof. dr. C. Ponsioen, prof. dr. Mark Benninga, and dr. J Keller

**Recommendation: Irrespective of the treatment indication, all FMT recipients and donors should routinely receive follow-up for early onset (<30 days) adverse events. Clinicians preferably follow-up FMT recipients and donors for 10 years or longer to fully establish efficacy, adverse events and disease development. A National Registry should be developed to register and evaluate patients by an independent committee. Follow-up in children can be extended to a period of 30 years, depending on the FMT indication or study design.**

The gut microbiota is a complex consortium with many components that have never been characterized. Currently, knowledge is not available regarding the impact of transferring these complex communities from one individual to another, although many studies in mice indicate that the composition of the gut microbiota can affect host susceptibility to various diseases.

Follow-up after FMT varies between studies and is strongly dependent upon study design and outcomes. Post-FMT surveillance can be performed by outpatient visits, telephone interviews, electronic diary and by standardized questionnaires. The duration of follow up also varies but the maximum period was never longer than 8 years (1). Post-FMT follow up should take into account:

1) Clinical outcome in recipients of FMT

2) Early and late adverse events of FMT recipients (annex I and II)

3) Development of new diseases in donors that can influence recipients health (annex III).

**Early adverse events** after FMT for CDI are usually **mild**: self-limiting GI symptoms have been the most frequently reported adverse events, and are typically short-lived, resolving in hours - days. Early serious adverse events are often procedure-related, for instance: perforation, aspiration (pneumonia), gastrointestinal haemorrhage (anticoagulans), sedation complications etc. In addition, non-procedure related serious adverse events include infections/sepsis.

**Post-FMT serious adverse events** can be defined as “significant morbidity necessitating hospital admission or resulting in death during the follow up period.” Other reported post-FMT SAE include flares of IBD, recurrent UTI, new onset autoimmune diseases/metabolomic diseases, microscopic colitis, malignancies, peripheral neuropathy and psychiatric syndrome. It is often difficult to assess the association with FMT, but all post FMT SAE should be registered and evaluated by an independent expert panel. This expert panel will be composed by independent scientists and physicians who are not involved in FMT studies or associated with the NDFB.

Of greater concern and uncertainty is the possibility **of long-term AEs**. The possibility that gut microbiota associated with a disease phenotype (e.g., metabolic syndrome, cardiovascular disease, cancer, psychological disorders) will be transplanted and result in chronic disease in recipients must be assessed.

**A long-term safety follow-up is currently lacking for both recipients and donors**. Self-screening questionnaires which focus on high risk behaviors for blood-borne infections, questionnaires that focus on previous potential transferable medical conditions and adaptations from the Blood Banks Donor are necessary for an appropriate follow-up of donors.The working group thinks that a screening process could be made mandatory for at least a period of 10 years after the last donation, though it may be prolonged in donating children.

We therefore propose a “**FMT national registry”.** This will include follow-up information from the patient’s healthcare provider at 1 month, 6 months, 1 year, and 2 years after FMT as well as direct communication with patients at least annually up to 10 years after FMT. Follow-up information to be collected will be designed to assess potential short-term and long-term safety, and effectiveness.

Both recipients and donors should provide informed consent for follow-up and collection of stool samples for microbiota composition. For pediatric patients under the age of 12 years, consent is given by a guardian.

**Possible adverse events should be registered as**:

**1. Not Related:**

• Temporal relationship is lacking (e.g., the event occurred before FMT); or

• Other causative factors explain the event (e.g. pre-existing condition, other concomitant treatment);

**2. Possibly Related:**

• Positive temporal relationship (e.g., the event occurred within a reasonable time frame following FMT); and

• The SAE is possibly explained by FMT, and there is a lack of other causal factors.

**3. Related:**

• Positive temporal relationship (e.g., the event occurred within a reasonable time frame following FMT); and

• The SAE is more likely explained by FMT than by other causes.

**The FMT AE Committee** will oversee the adverse events. This Committee will report to the daily board of the NDFB and subsequent to the supervisory board of the NDFB, the LUMC board and IGJ. The Committee will be comprised of members who are not involved in FMT studies or are affiliated to the NDFB

**Annex I: FMT Short-term Adverse Outcomes (within 30 days) in recipients**

• Procedure-related

o	Sedation complication

o	Bleeding

o	Perforation

o	Regurgitation of donor feces

- Aspiration of donor feces
- Aspiration pneumonia
- Bowel perforation
- Sedation complication
- Other

• Symptoms post-FMT (within 30 days, specify which day/weeks)

o Diarrhea

o Constipation

o Nausea and/or vomiting

o Bloating

o Abdominal pain

o Fever

o Headache

o Weight gain or loss (in relation to weight before CDI episode)

- Other

• Surgeries or other Procedures

o Describe

• Documented Infection (any)

o Specify site/organism

o FMT-related (related, possibly related, unrelated)

• Hospitalization

o Reason for hospitalization

o FMT-related (related, possibly related, unrelated)

• Life-threatening experience

o Describe/diagnosis

o FMT-related (related, possibly related, unrelated)

• Death

o Cause of death

o FMT-related (related, possibly related, unrelated)

o Site of death

▪ Hospital

▪ Home

▪ Convalescent or skilled nursing facility

**Annex II. FMT Long-term Adverse Outcomes (up to 2 years by physician report and 10 years by patient report) in recipients**

• Characteristics of the patient

o Height

o Weight

• Serious Infection (HIV, viral hepatitis, prion, etc)

• Use of new drugs

o Describe

• Surgeries or other Procedures

o Describe

• Diagnosis of any new condition

o Autoimmune (hypothyroid, ITP, RA, SLE, MS, celiac, Type I diabetes, Sjogrens)

o Asthma

o Allergy/atopy

o Metabolic disease

▪ Diabetes II

▪ Obesity

o Psychiatric disorder

o Neurologic disease

▪ Parkinson’s disease

▪ Amyotrophic lateral sclerosis (ALS)

▪ Autism spectrum diagnosis

o Cardiovascular disease

▪ Myocardial infarction

▪ Coronary artery revascularization

▪ Cerebrovascular accident

▪ Hypertension

o Colon cancer

o Other malignancy

o Inflammatory bowel disease

▪ Crohn’s

▪ Ulcerative colitis

▪ IBD-U

o IBS

▪ IBS-C

▪ IBS-D

▪ IBS-M

o Other

• Hospitalization

o Reason for hospitalization

o FMT-related (related, possibly related, unrelated)

• Life-threatening illness

o Describe/diagnosis

o FMT-related (related, possibly related, unrelated)

• Death

o Cause of death

o FMT-related (related, possibly related, unrelated

**Annex III. FMT Follow-up of donors (up to 10 years after last donation)**

• Characteristics of the donor

o Height

o Weight

• Serious Infection (HIV, viral hepatitis, prion, etc)

• Use of new drugs

o Describe

• Surgeries or other Procedures

o Describe

• Diagnosis of any new condition

o Autoimmune

o Asthma

o Allergy/atopy

o Metabolic disease

▪ Diabetes II

▪ Obesity

o Psychiatric disorder

o Neurologic disease

▪ Parkinson’s disease

▪ Amyotrophic lateral sclerosis (ALS)

▪ Autism spectrum diagnosis

o Cardiovascular disease

▪ Myocardial infarction

▪ Coronary artery revascularization

▪ Cerebrovascular accident

▪ Hypertension

o Colon cancer

o Other malignancy

o Inflammatory bowel disease

▪ Crohn’s

▪ Ulcerative colitis

▪ IBD-U

o IBS

▪ IBS-C

▪ IBS-D

▪ IBS-M

o Other

• Hospitalization

o Reason for hospitalization

• Life-threatening illness

o Describe/diagnosis

• Death

o Cause of death
