## Supplementa File S3 for "Safety and feasibility of fecal microbiota transplantation for Parkinson’s disease patients: a protocol for a self-controlled interventional donor-FMT pilot study"

Andrea E. van der Meulen-de Jong (Department of Gastroenterology and hepatology, Leiden University Medical Center, Leiden, the Netherlands), Ed J. Kuijper (Department of Medical Microbiology, Leiden University Medical Center, Leiden, the Netherlands; Center for Infectious Disease Control, National Institute for Public Health and the Environment (Rijksinstituut voor Volksgezondheid en Milieu, RIVM), Bilthoven, the Netherlands), Elisabeth M. Terveer (Department of Medical Microbiology, Leiden University Medical Center, Leiden, the Netherlands), Eric Berssenbrugge (Department of Medical Microbiology, Leiden University Medical Center, Leiden, the Netherlands), Hein Verspaget (Department of Biobanking, Leiden University Medical Center, Leiden, the Netherlands), Jacobus J. van Hilten (Department of Neurology, Leiden University Medical Center, Leiden, the Netherlands), Jannie G.E. Henderickx (Department of Medical Microbiology, Leiden University Medical Center, Leiden, the Netherlands), Jelle J. Goeman (Department of Biostatistics, Leiden University Medical Center, Leiden, the Netherlands), Josbert J. Keller (Department of Gastroenterology, Haaglanden Medical Center, Den Haag, the Netherlands), Karuna E.W. Vendrik (Department of Medical Microbiology, Leiden University Medical Center, Leiden, the Netherlands; Center for Infectious Disease Control, National Institute for Public Health and the Environment (Rijksinstituut voor Volksgezondheid en Milieu, RIVM), Bilthoven, the Netherlands), Maria F. Contarino (Department of Neurology, Leiden University Medical Center, Leiden, the Netherlands; Department of Neurology, Haga Teaching hospital, The Hague, the Netherlands), Martijn Bauer (Department of Internal Medicine, Leiden University Medical Center, Leiden, the Netherlands), Romy Zwittink (Department of Medical Microbiology, Leiden University Medical Center, Leiden, the Netherlands),Vlada O. Bekker-Chernova (Department of Medical Microbiology, Leiden University Medical Center, Leiden, the Netherlands; Department of Neurology, Leiden University Medical Center, Leiden, the Netherlands).
